# Cross-ancestral GWAS identifies 29 novel variants across Head and Neck Cancer subsites

**DOI:** 10.1101/2024.11.18.24317473

**Authors:** E Ebrahimi, A Sangphukieo, HA Park, V Gaborieau, A Ferreiro-Iglesias, B Diergaarde, W Ahrens, L Alemany, LMRB Arantes, J Betka, SV Bratman, C Canova, MSC Conlon, DI Conway, M Cuello, M Curado, A de Carvalho, J de Oliviera, M Gormley, M Hadji, S Hargreaves, CM Healy, I Holcatova, RJ Hung, LP Kowalski, P Lagiou, A Lagiou, G Liu, GJ Macfarlane, AF Olshan, S Perdomo, LF Pinto, JV Podesta, J Polesel, M Pring, H Rashidian, RR Gama, L Richiardi, M Robinson, PA Rodriguez-Urrego, SA Santi, DP Saunders, SC Soares-Lima, N Timpson, M Vilensky, SV von Zeidler, T Waterboer, K Zendehdel, A Znaor, P Brennan, HEADSpAcE Consortium, J McKay, S Virani, T Dudding

**Author notes:** A list of authors and their affiliations appears at the end of the paper. Co-last, co-corresponding authors: Shama Virani, *Tom Dudding. **Conflict of Interest Disclosure:** Tim Waterboer serves on advisory boards for MSD (Merck) Sharp & Dohme. Scott V Bratman is inventor on patents related to cell-free DNA mutation and methylation analysis technologies that are unrelated to this work and have been licensed to Roche and Adela, respectively. Scott V Bratman is a co-founder of and has ownership in Adela.

## Abstract

In this multi-ancestry genome-wide association study (GWAS) and fine mapping study of head and neck squamous cell carcinoma (HNSCC) subsites, we analysed 19,073 cases and 38,857 controls and identified 29 independent novel loci. We provide robust evidence that a 3’ UTR variant in *TP53* (rs78378222, T>G) confers a 40% reduction in odds of developing overall HNSCC. We further examine the gene-environment relationship of *BRCA2* and *ADH1B* variants demonstrating their effects act through both smoking and alcohol use. Through analyses focused on the human leukocyte antigen (HLA) region, we highlight that although human papilloma virus (HPV)(+) oropharyngeal cancer (OPC), HPV(-) OPC and oral cavity cancer (OC) all show GWAS signal at 6p21, each subsite has distinct associations at the variant, amino acid, and 4-digit allele level. We also defined the specific amino acid changes underlying the well-known DRB1*13:01-DQA1*01:03-DQB1*06:03 protective haplotype for HPV(+) OPC. We show greater heritability of HPV(+) OPC compared to other subsites, likely to be explained by HLA effects. These findings advance our understanding of the genetic architecture of head and neck squamous cell carcinoma, providing important insights into the role of genetic variation across ancestries, tumor subsites, and gene-environment interactions.

## Main

Head and neck squamous cell carcinomas (HNSCC) are a heterogeneous group of cancers originating primarily in the oral cavity (OC), oropharynx (OPC), larynx (LA) and hypopharynx (HPC). Currently, HNSCC is ranked the 6th most common cancer globally, although incidence is predicted to increase 30% by 2030^1,2^. Tobacco smoking and alcohol consumption are major risk factors, particularly in high income countries, contributing to 72% of cases when used together, while betel quid/areca nut products significantly increase risk in some Asia-Pacific populations^2^. HNSCC subsites can be differentially affected, with smoking more strongly linked to laryngeal cancer and drinking more strongly linked to OC/OPC^3^. There has been a decline in smoking in high income countries, as such, the increasing incidence could be due to changes in etiology^4^. Infection with human papillomavirus (HPV), particularly HPV type 16, is a recently identified causal risk factor for OPC^5–7^ and the proportion of HPV-associated OPCs is highest in high-income countries (63%-85%)^8^. Disparities in epidemiology, risk, and prognosis highlight the recognition of HPV-associated OPC as a distinct biological entity^9^.

Although a limited number of genome-wide association studies (GWAS) have been conducted on HNSCC, a germline contribution to HNSCC risk has been established, with multiple susceptibility loci associated with risk. These include the 4q23 locus (*ADH1B, ADH7*) linked to genes involved in alcohol metabolism, the 5p15 locus (*TERT-CLPTM1L*) associated with genes responsible for DNA stability maintenance, and the 6p21 and 6p22 loci, mostly within the human leukocyte antigen (HLA) region, corresponding with genes regulating the innate immune response ^10–14^. The 6p locus within the HLA region has been a specific area of focus for HPV-driven cancers, with the hypothesis being that variants influencing immune response to viral antigens would be most relevant for risk ^10,13,14^. However, there is an emerging role for the immune microenvironment for other HNSCC subsites^15^, suggesting that the HLA may confer risk separately in other HNSCC subsites, potentially via non-HPV mechanisms. Previous GWAS were limited in sample size for HNSCC subsites making inference between subsites, particularly for HLA, difficult. They were also conducted predominantly in subjects of European ancestry, limiting generalizability of findings.

Despite knowledge of the major risk factors and several risk loci for HNSCC, identifying those who will develop cancer is still difficult. Not all smokers develop cancer and risk loci only offer a fractional change in risk at the population level. The interaction between environmental factors and risk loci may help explain additional risk and have been reported for lung cancer (smoking)^16^, colorectal cancer (alcohol)^17^ and bladder cancer (arsenic exposure)^18^ among others. Studies investigating these interactions need large sample sizes and individual level exposure data harmonised across studies which, is often not possible in large GWAS meta-analyses.

Here, we perform a cross-ancestry GWAS of HNSCC using individual level data, bringing together studies from Europe, North America, South America, South Asia and the Middle East. We identify multiple novel genetic risk susceptibility loci, determine shared and unique risk loci across subsites, explore interactions between genetic and environmental factors in HNSCC risk and conduct fine mapping of the HLA region. This work lays the foundation for identifying HNSCC susceptibility loci with increased representation from non-European populations.

## Results

### Cross-ancestral meta-analysis identifies 18 novel genetic loci across HNSCC subsites

In this cross-ancestral meta-analysis of two pooled individual level datasets (Table S1), we evaluated 13,092,551 genetic variants in 19,073 HNSCC cases and 38,857 controls. Of the HNSCC cases, there were 5,596 (29%) oral cavity (OC), 5,411 (28%) oropharyngeal (OPC), 4,409 (23%) laryngeal (LA), 898 (5%) hypopharyngeal (HPC), 2,759 (14%) unknown (either unknown primary site or not available) or overlapping sites. HPV status was available for 68% of OPC cases, of which 3,685 (60%) were HPV(+) (Table S2).

We identified 18 novel genome-wide associated variants, including two specific to non-European ancestry (Table 1, Figure 1, Figure S1) and validated 6 previously identified loci (Table S3, S4). A specific focus on the HLA region identified 11 further novel variants. The proportion of variance attributable to genome-wide SNPs for HNSCC overall was 14% (95% Confidence Interval (CI): 12.7, 15.3). Across subsites, heritability ranged from 7.6% (95% CI: 5.0, 10.2) for HPC to 29.1% (95% CI: 25.5, 32.7) for HPV(+) OPC (Table S5).

**Figure 1.**
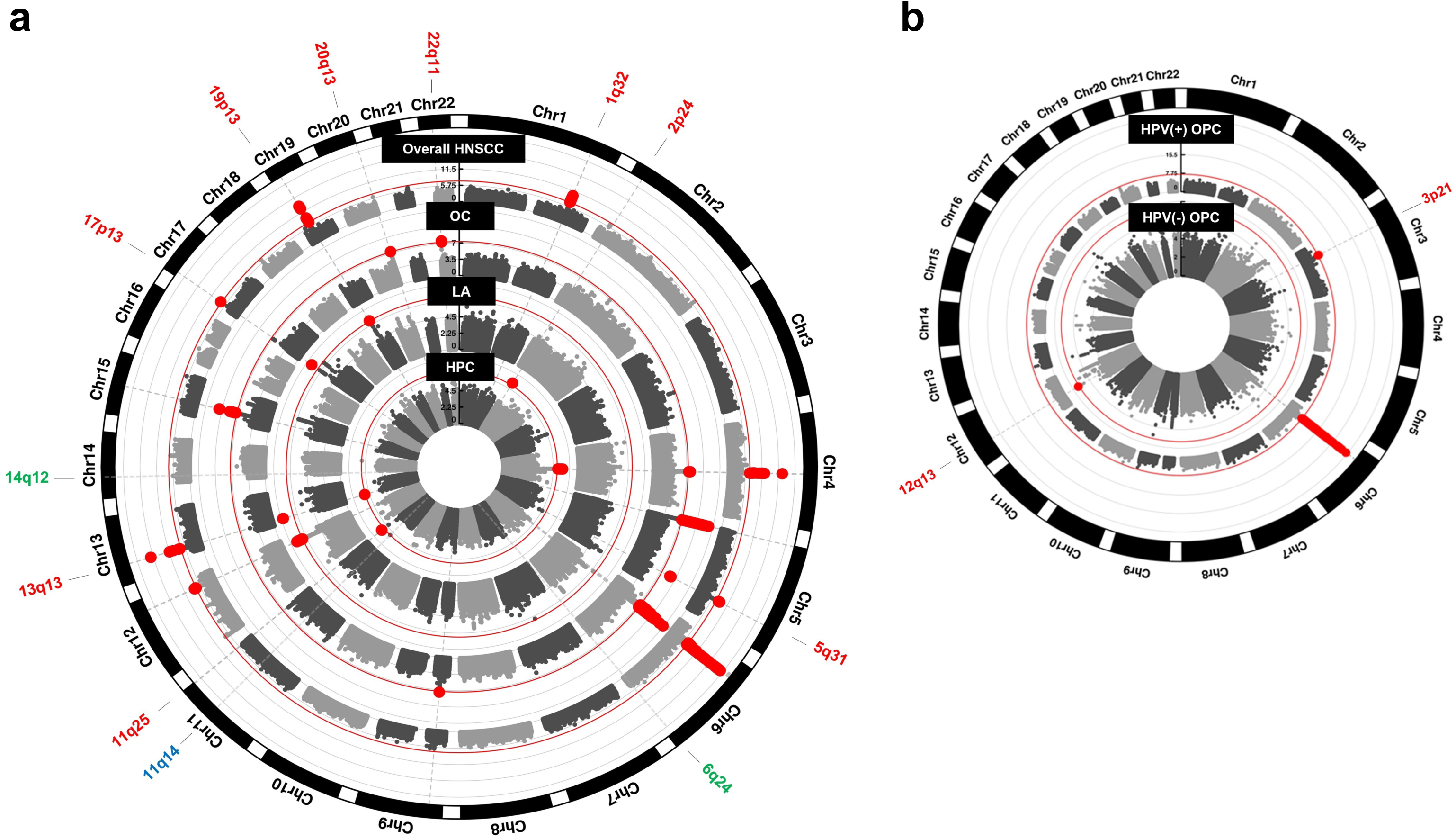
Novel loci identified for HNSCC. a) Circular Manhattan plots showing novel risk loci identified in this study. Red labels indicate the cytogenetic locations of novel signals identified in meta-analyses for all sites combined or subsite-specific. Blue labels represent novel loci identified in the European group only, and green labels indicate novel loci identified in the Mixed group only. Red lines mark the threshold for genome-wide significance (p = 5×10^-8^). b) Circular Manhattan plots from the GWAS analyses of HPV(+) and HPV(-) oropharyngeal cancer. Separate Manhattan plots for each group can be found in Supplementary Figure 1.

**Table 1.**
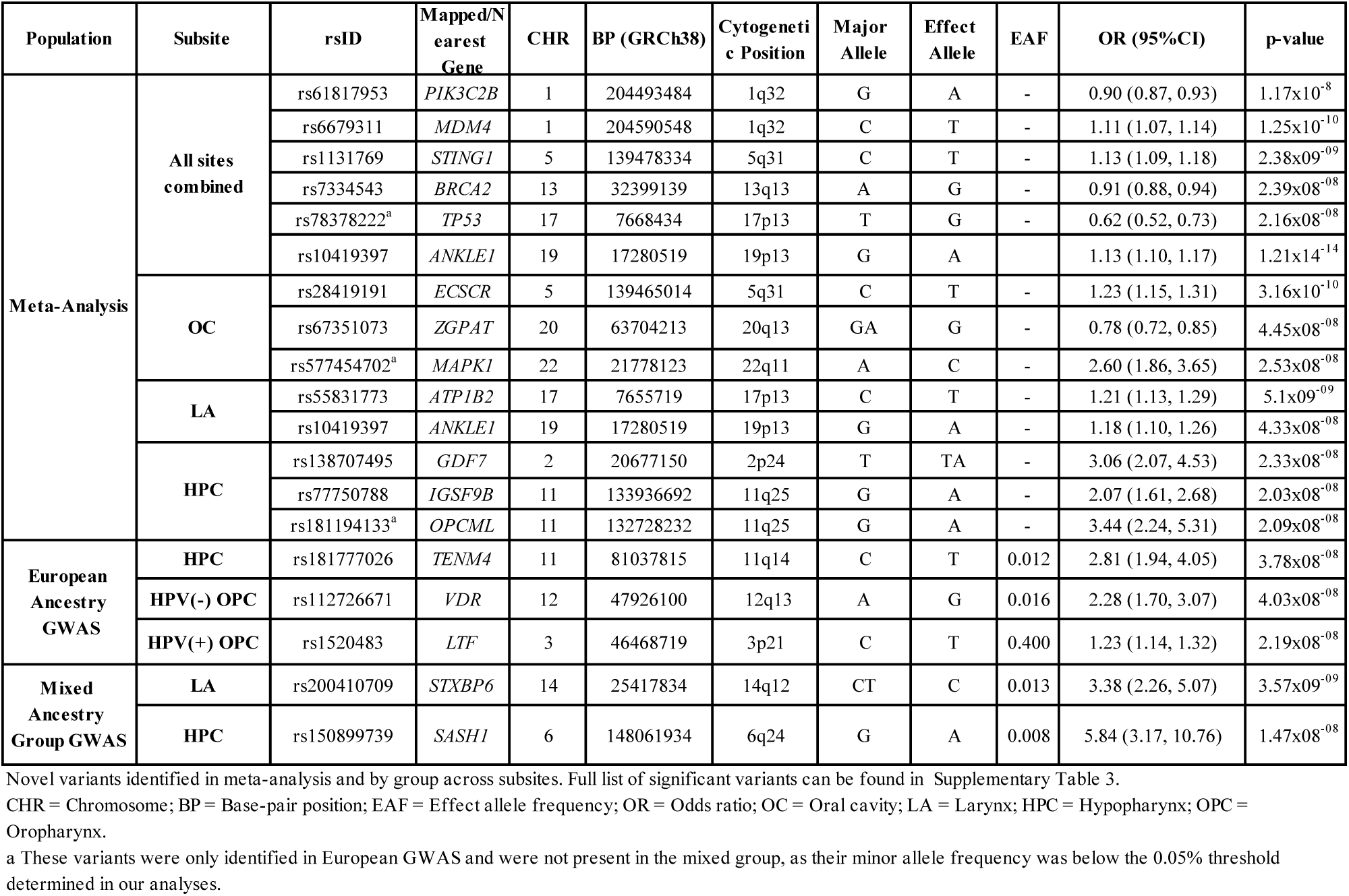
Summary of Novel Genetic Variants Identified in European and Mixed Groups Through GWAS and Meta-Analysis.

For overall HNSCC, two novel variants in the 1q32 region were identified. rs61817953, near *PIK3C2B,* was associated with decreased risk (OR (95%CI)=0.90 (0.87, 0.93), p_meta_= 2.17×10^-8^) and rs6679311 near *MDM4,* a strong negative regulator of p53, was associated with increased risk (OR (95% CI)=1.11 (1.07, 1.14), p_meta_=1.25×10^-10^) (Figure S2). The latter is in moderately high LD (r^2^=0.75) with rs4245739, an *MDM4* 3’ UTR variant known to increase breast^19^ and prostate^20^ cancer risk. At the 13q13 locus, rs7334543, a novel 3’ UTR variant in *BRCA2* was associated with decreased risk of overall HNSCC (OR (95%CI)= 0.91 (0.88, 0.94), p_meta_=2.39×10^-8^) and was independent from rs11571833, stop gain variant previously identified in this region for UADTs^12^. Within those of European ancestry, rs78378222 a 3’ UTR variant in *TP53*, was associated with a reduced risk of HNSCC overall, (OR (95% CI)=0.62 (0.52, 0.73), p=2.16×10^-8^) (Figure 2a). The effect was consistent across all non-HPV related HNSCC subtypes but had no effect in HPV(+) OPC. The T>G allele frequency of rs78378222 is 0.01 in EUR, 0.002 in AFR and AMR populations, and nearly absent in all other 1000 Genomes super-populations; as such, there was no effect of this variant in the Mixed ancestry. Given its low frequency, technical validation was performed in 2,370 samples and concordance with imputed data was 99.9% (Table S6). There was strong evidence for this variant being correlated with decreased gene expression of *TP53* (Figure 2b) (Table S7). This variant is in the poly-adenylation signal of the *TP53* gene and potentially leads to impaired 3’ end processing of *TP53* mRNA^21^. rs78378222 is located within a highly conserved sequence (TTTTATTGTAAAATA -> TTGTATTGTAAAATA) that appears to be crucial for miRNA binding. This region is predicted to interact with 5 different microRNAs (miRNAs) as suggested by TarBase (https://dianalab.e-ce.uth.gr/tarbasev9) (Figure 2c).

**Figure 2.**
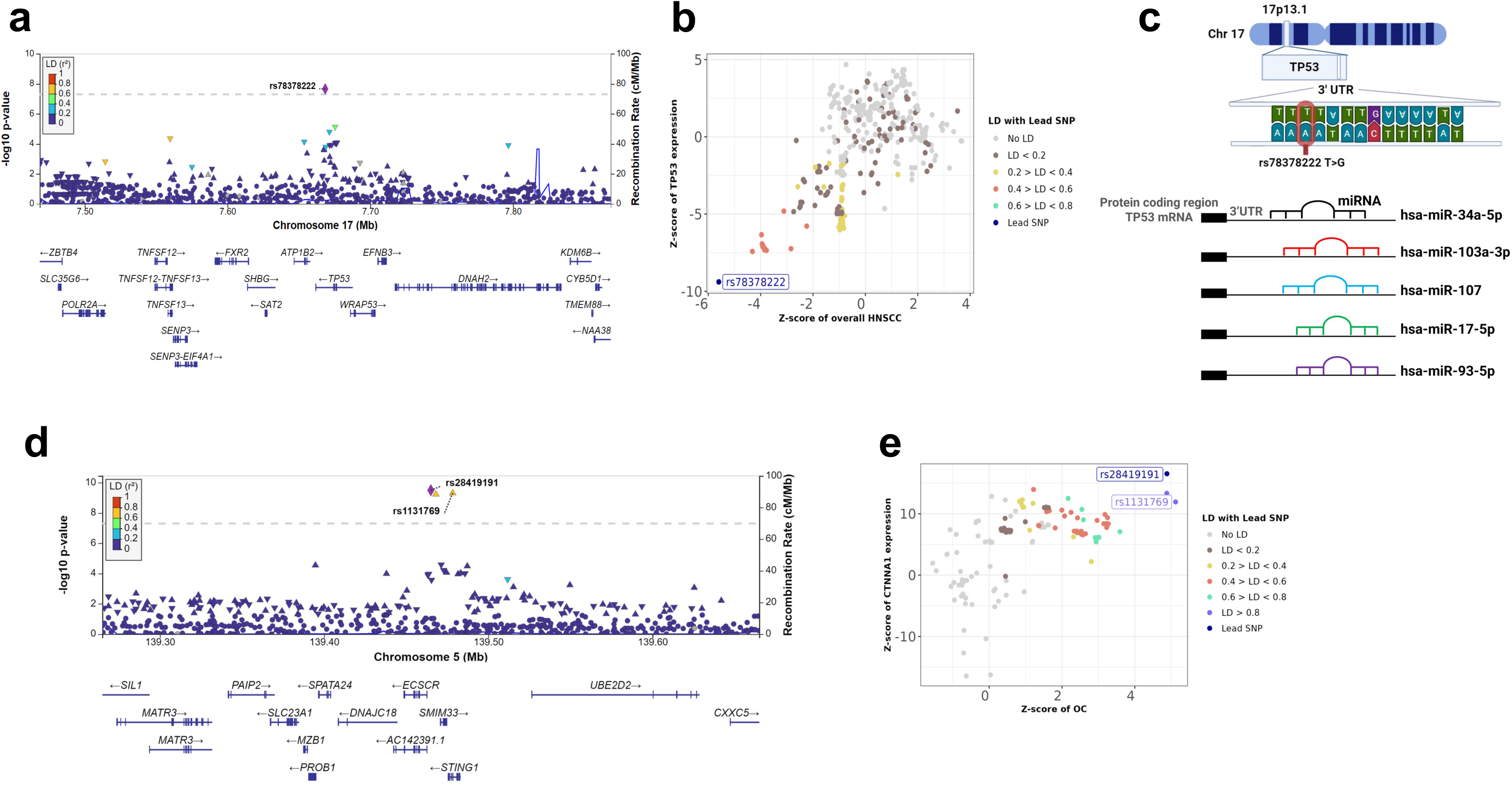
Regional and Colocalization Analysis of *TP53* and *STING1* Variants. a) Regional plot of the 3’ UTR variant rs78378222 in *TP53* at chromosome 17p13. The x-axis represents chromosomal location, while the y-axis displays the –log10 p-value. The dotted line marks the genome-wide significance threshold of 5×10^−8^. Single nucleotide polymorphisms (SNPs) are color-coded according to their linkage disequilibrium (r²) with rs78378222. b) Z-Z locus plot showing rs78378222, the lead variant, is associated with reduced TP53 expression in whole blood, with a high PP4 score of 99%. c) The cytogenetic location of rs78378222, along with its sequence and allele change, is shown at the chromosomal level. The variant overlaps with multiple predicted microRNA binding sites. d) Regional plot of rs28419191 (intergenic) and rs1131769 (*STING1*) at 5q31 from the oral cavity (OC) meta-analysis (r² = 0.93). e) Z-Z locus plot showing colocalization of rs28419191 and rs1131769 with *CTNNA1* expression in whole blood, both with a PP4 score of 99%.

For OC, three novel loci were identified (Table 1). First, rs28419191, an intergenic variant at 5q31 associated with an increased risk of OC (OR (95% CI)=1.23 (1.15, 1.31), p_meta_=3.16×10^-10^). This variant was in high LD with rs1131769 (r^2^=0.93), a missense variant in *STING1* which was a novel loci for overall HNSCC risk (OR=1.13 (1.09, 1.18), p_meta_=2.38×10^-10^) (Table 1, Figure 2d). Both rs28419191 and rs1131769 correlated with expression of catenin alpha 1 (*CTNNA1),* a gene related to RNA and actin filament binding, but not *STING1* expression in whole blood; as such, the function of this variant is unclear (Figure 2e). The second novel variant rs67351073, located at 20q13 in Zinc Finger CCCH-Type And G-Patch Domain Containing (*ZGPAT*), was associated with reduced risk of OC (OR (95%CI)=0.78 (0.72, 0.85), p_meta_=4.45×10^-8^). A highly correlated variant seen within the European ancestry only (rs4809325, r^2^=0.97), which also decreased OC risk, was correlated with decreased *ZGPAT* gene expression in whole blood (PP4 score=0.97) and increased *LIME1* gene expression in oesophageal and lung mucosa (Figure S3, Table S7). Finally, a novel, rare, European ancestry-specific intronic variant, rs577454702, located in the mitogen-activated protein kinase 1 (*MAPK1)* gene at 22q11, was associated with a large increased risk of OC (OR (95%CI)=2.60 (1.86, 3.65), p=2.53×10^-8^).

For laryngeal cancer, rs55831773, a novel splice variant, mapping to *ATP1B2* was associated with increased risk (OR (95% CI)=1.21 (1.13, 1.29), p_meta_=5.1×10^-9^). *ATP1B2* is in close proximity to *TP53* but conditional analyses suggest this variant is independent of the rare *TP53* 3’ UTR variant described for overall HNSCC. There was also no evidence that rs55831773 alters *TP53* expression, further suggesting independent effects of these two variants (Figure S4). A novel intronic variant, rs10419397, located in a gene dense region of 19p13 was also strongly associated with LA (OR (95%CI)=1.13 (1.10, 1.17), p_meta_=1.21×10^-14^). This variant has been found to associate with mitochondrial dysfunction^22,23^ and is in very high LD with several variants associated with risk of other cancers, including rs4808616 (r^2^>0.99), a 3’ UTR for *ABHD8* linked to breast and lung cancers^24^. rs200410709 is a novel variant which showed increased risks in the Mixed ancestry but with no evidence of effect in Europeans. It is a deletion variant in an intergenic region, adjacent to the Syntaxin Binding Protein 6 (*STXBP6*) gene (14q12), and was associated with increased risk of LA (OR (95% CI)=3.38 (2.26, 5.07), p=3.57×10^-9^) (Figure S5).

Five HPC specific variants were identified for the first time. rs138707495, a rare (MAF: European = 0.009, Mixed = 0.005) variant located in the 3’ UTR of *GDF7* (OR (95%CI)=3.06 (2.07, 4.53), p_meta_=2.33×10^-8^), rs77750788 at 11q25 near *IGSF9B* (OR (95%CI)=2.07 (1.61, 2.68), p_meta_=2.03×10^-8^) and rs181194133 an intronic variant in *OPCML* (OR (95%CI)= 3.44 (2.24, 5.31), p_meta_= 2.09×08^-08^) were all associated with increased risk of HPC in the cross-ancestral meta-analysis. Within the European group, rs181777026 (11q14) located near *TENM4* was associated with increased risk of HPC. Conversely, rs150899739 (6q24), which showed an increased risk in the Mixed ancestry but no effect in Europeans, is within *SASH1* and greatly increased the risk for HPC (OR (95% CI)=5.84 (3.17, 10.76), p=1.47×10^-8^) (Figure S6).

At 3p21, rs1520483, a novel intronic variant in the lactotransferrin (*LTF*) gene, associated with an increased risk of HPV(+) OPC (OR (95% CI)=1.23 (1.14, 1.32), p=2.19×10^-8^) in Europeans. *LTF* acts as a transcription factor, inducing expression of innate immune related genes for antiviral host defence^25,26^.

rs112726671, a variant near the vitamin D receptor (VDR) gene, was associated with risk of HPV(-) OPC (OR (95%CI)=1.23 (1.14, 1.32), p_meta_=2.19×10^-8^). This variant is independent from rs35189640, a nearby variant previously identified to increase risk of HPV(-) OPC (r^2^=0.0005)^10^.

### Refining previously identified HNSCC risk variants

Loci identified in previous GWAS of HNSCCs at 4q23 *(ADH1B*, *ADH1C*, *ADH7*), 5p15 *(CLPTM1L*), 6p21 (HLA), 6p22 *(ZNRD1-AS1),* 9p21 (*CDKN2B-AS1*), 12q24 *(ALDH2*) and 15q21 (*FGF7*) were validated here. Notably, rs11571833 (13q13), the rare (MAF: Europeans=0.009, Mixed=0.007) stop gained variant, resulting in a stop codon 93 amino acids early in the BRCA2 protein, was strongly associated with an increased risk of LA (OR (95% CI)=2.09 (1.65, 2.66), p_meta_=1.57×10^-9^) and HPC (lead variant for HPC-rs11571815: OR (95% CI)=2.73 (1.61, 3.90), p_meta_=3.99×10^-8^) separately. Previous GWAS combining lung and aerodigestive tract cancers as well as studies using targeted genotyping have found this variant to substantially increase risk for smoking related cancers^27^, however this is the first time this variant was identified in within specific subsites.

### Distinct interactions of smoking and alcohol use with risk variants

rs11571833 the *BRCA2* stop-gained variant validated here showed clear evidence of a dose-response effect across smoking and drinking strata, but the variant did not correlate with variants related to smoking-related behaviours such as smoking initiation or cigarettes per day in colocalization analysis (Table S7). However, the variant effect was present in both non-drinking smokers and non-smoking drinkers, suggesting the risk increasing effect of rs11571833 requires either carcinogenic influence. This *BRCA2* variant shows a similar gene-environment interaction separately within the European and Mixed ancestries, despite differences in sample size (Figure 3a).

**Figure 3.**
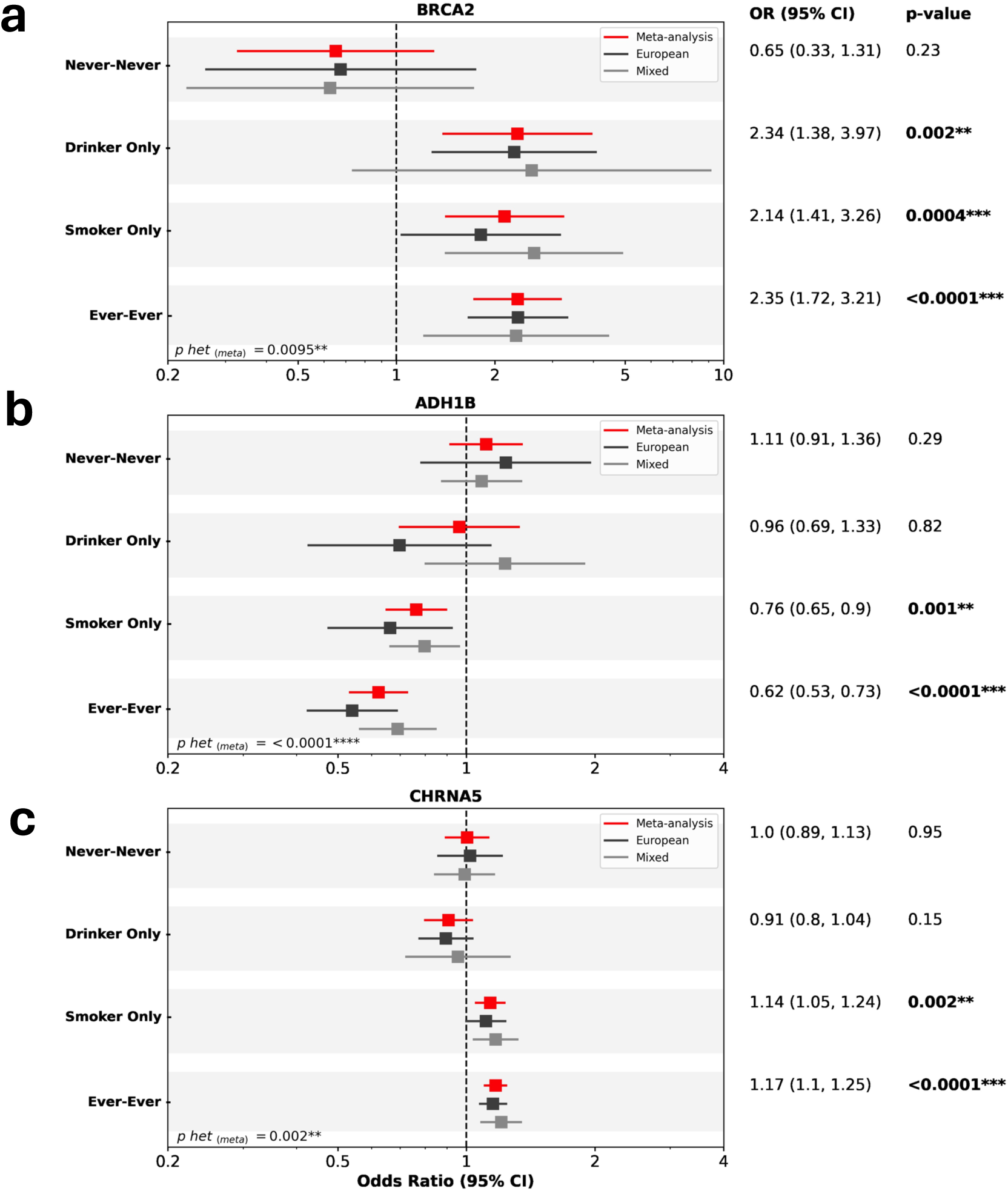
Gene-environment interactions with alcohol and smoking. Effect estimates for a) rs11571833 (*BRCA2*) b) rs1229984 (*ADH1B*) and c) rs58365910 (*CHRNA5*) stratified by smoking and drinking (Never smoker-Never drinker, Smoker Only, Drinker Only, and Ever smoker-Ever drinker) from the meta-analysis, and within European, and Mixed groups. Only the odds ratios, p-value and p-heterogeneity (p-het) for the meta-analysis are shown here.

We confirm that rs1229984, the well-described missense variant in the *ADH1B* gene, has a strong protective effect on OC which is only seen in smokers or in drinkers when stratifying by use (Figure S6b). However, we measure a strong correlation between rs1229984 and variants associated with alcoholic drinks per week but not cigarettes per day or smoking initiation (Table S7). To separate out the linked behaviors of smoking and drinking we investigated the association in combinations of drinking and smoking status. These analyses confirm rs1229984 has an effect in those who smoke and drink and in non-drinking smokers but not non-smoking drinkers, suggesting the mechanism through smoking as may be more important (Figure 3b). Interactions with smoking and drinking for *ADH1C* and *ADH7* were less clear.

rs58365910 near *CHRNA5*, known to alter smoking intensity^36^ showed a suggestive association with LA consistent effects across the European and Mixed ancestries (Figure S8). The increasing risk effect of this variant was correlated with increased smoking intensity and when evaluated by exposure group, this variant shows a clear interaction with smoking but not alcohol use (Figure 3c).

### Novel Loci in the HLA Region specific to oral cavity and oropharynx cancer

Our genome-wide results highlight heterogeneity in the Human Leukocyte Antigen (HLA) region, which encodes genes involved in immune response, across HNSCC subsites. For HPV(+) OPC, signals were identified at both 6p21 and 6p22 but for OC only the 6p21 signal was seen. The HLA region is particularly susceptible to genetic diversity across populations and is highly polymorphic with a dense LD structure. To account for this, genotyped variants in this region were re-imputed to define variants, amino acid changes and 4-digit alleles, which were then analysed separately using fine mapping strategies to identify independent signals. Independence of signals was carefully evaluated using linkage and conditional analysis (Tables S8, S9).

Overall, 19 independent signals reached significance (Tables S10, S11). Eleven novel risk variants were identified specific to OC, HPV(+) OPC, HPV(-) OPC, and for HNSCC overall (Table 2, Figure S9).

**Table 2.** Summary of Novel Genetic Variants Identified Across Ancestry-Specific and Meta-Analysis of HLA-fine mapping in all sites combined and subsite specific.

| Population | Subsite | Variant | Gene | Position (hg19) | Locus; Cytoband | Impact | Ref<sup>a</sup> | Effect Allele<sup>a</sup> | OR (95% CI) | p-value<sup>b</sup> |
| --- | --- | --- | --- | --- | --- | --- | --- | --- | --- | --- |
| Meta-Analysis | All sites combined | Chr6:33046667 | HLA-DPB1 | Chr6:33046667 | class II | intron | C | T | 1.11 (1.07,1.14) | 1.32x10<sup>-8</sup> |
|  |  | rs28360051 | PSORS1C3 | Chr6:31142261 |  | intron | G | A | 1.23 (1.14,1.34) | 1.91x10<sup>-7</sup> |
|  | HPV(-) OPC | rs1131212 | HLA-B | Chr6:31324526 | class I | Gln94His | C | G | 1.33 (1.19,1.49) | 5.33x10<sup>-7</sup> |
|  | HPV(+) OPC | DRB1 37Asn/Ser | HLA-DRB1 | Chr6:32552051 | class II | Amino acid change | Ab | Pr | 0.68 (0.63,0.73) | 3.22x10<sup>-23</sup> |
|  |  | rs4143334 | ZDHHC20P2 | Chr6:31348200 | class I | Non coding transcript exon | A | G | 1.89 (1.51,2.35) | 1.91x10<sup>-8</sup> |
|  |  | DRB1 233Thr | HLA-DRB1 | Chr6:32548048 | class II | Amino acid change | Ab | Pr | 1.27 (1.17,1.38) | 7.15x10<sup>-9</sup> |
|  |  | B 67Cys/Ser/Tyr | HLA-B | Chr6:31324536 | class I | Amino acid change | Ab | Pr | 0.81 (0.74,0.88) | 1.33x10<sup>-06</sup> |
| Admixed | All sites combined | rs1536036 | ITPR3 | Chr6:33632014 |  | Intron | A | G | 0.85 (0.80,0.91) | 8.42x10<sup>-7</sup> |
| European | OC | DRB1 74Ala/Leu/Del | HLA-DRB1 | Chr6:32552625 | class II | Amino acid change | Ab | Pr | 0.82 (0.77,0.87) | 4.94x10<sup>-10</sup> |
|  |  | rs9267280 | MICB-DT | Chr6:31457633 | class I | Intron | G | A | 1.32 (1.19,1.47) | 3.48x10<sup>-7</sup> |
|  | HPV(+) OPC | HLA-B*51:01 | HLA-B | Chr6:31321767 | class I | Amino acid change | Ab | Pr | 1.9 (1.55,2.31) | 3.6x10<sup>-10</sup> |

a Ref/A1 allele is in binary marker format (Ab = Absent, Pr = Present) of classical HLA alleles, amino acid residue, HLA intragenic, insertion/deletions. (see: https://imputationserver.readthedocs.io/en/latest/pipeline)
b meta-analysis Pvalue of the final model including all significant independent variants adjusted by sex, imputation batch and PCs
HLA significance level = $2.4 \times 10^{-6}$ considering all variants in chr6

Three novel intronic variants were associated with risk of HNSCC overall. The Chr6:33046667 variant, near *HLA-DPB1* (OR (95% CI)= 1.11 (1.07, 1.14), p_meta_=1.32×10^-8^) and rs28360051 near *PSORS1C3* (OR (95% CI)= 1.23 (1.14, 1.34), p_meta_=1.91×10^-7^) both increased HNSCC risk. The rs28360051 variant was strongly driven by its effect in HPV(+) OPC. A novel intronic variant, rs1536036, mapping to *ITPR3*, a receptor that mediates the release of intracellular calcium, was protective for HNSCC overall (OR (95% CI)= 0.85 (0.80, 0.91), p= 8.42×10^-7^) only in the admixed ancestry.

For HPV(+) OPC, five novel variants were identified. rs4143334, in the noncoding transcript exon of *ZDHHC20P2* increased cancer risk (OR (95% CI)= 1.89 (1.51, 2.35), p_meta_=1.91×10^-8^). The remaining three had important functional significance. The first (DRB1 37Asn/Ser) causes an amino acid change in the antigen binding pocket (P9 pocket) of the beta chain of the HLA-DR protein and reduces HPV(+) OPC risk (OR (95% CI)= 0.68 (0.63, 0.73), p_meta_= 3.22×10^-23^).The second (HLA-B 67Cys/Ser/Tyr) is in an antigen binding pocket (B-pocket) of HLA-B and also results in decreased HPV(+) OPC risk (OR (95% CI)=0.81 (0.74, 0.88), p_meta_=1.33×10^-6^) (Figure 4a). The third (DRB1 233Thr), is in exon 5 of *DRB1* and increases risk of HPV(+) OPC (OR (95% CI)=1.27 (1.17, 1.38), p_meta_= 7.15×10^-9^). This amino acid change is in high LD with several others that are in the HLA-DR binding pocket, of which 5 have similar risk (Table S12). Accuracy of best-fit models, which included each amino acid in place of DRB1 233Thr, were found to be similar to the original model containing DRB1 233Thr (**△**BIC ±2), indicating that presence of any of these five amino acid changes—including DRB1 10Glu/Gln and 12Lys located in the HLA binding pocket—confers similar levels of risk (Figure 4b, Table S12). Within those of European ancestry, the novel HLA-B*51:01 allele increased risk of HPV(+) OPC (OR (95% CI)=1.9 (1.55, 2.31), p_meta_= 3.6×10^-10^).

**Figure 4.**
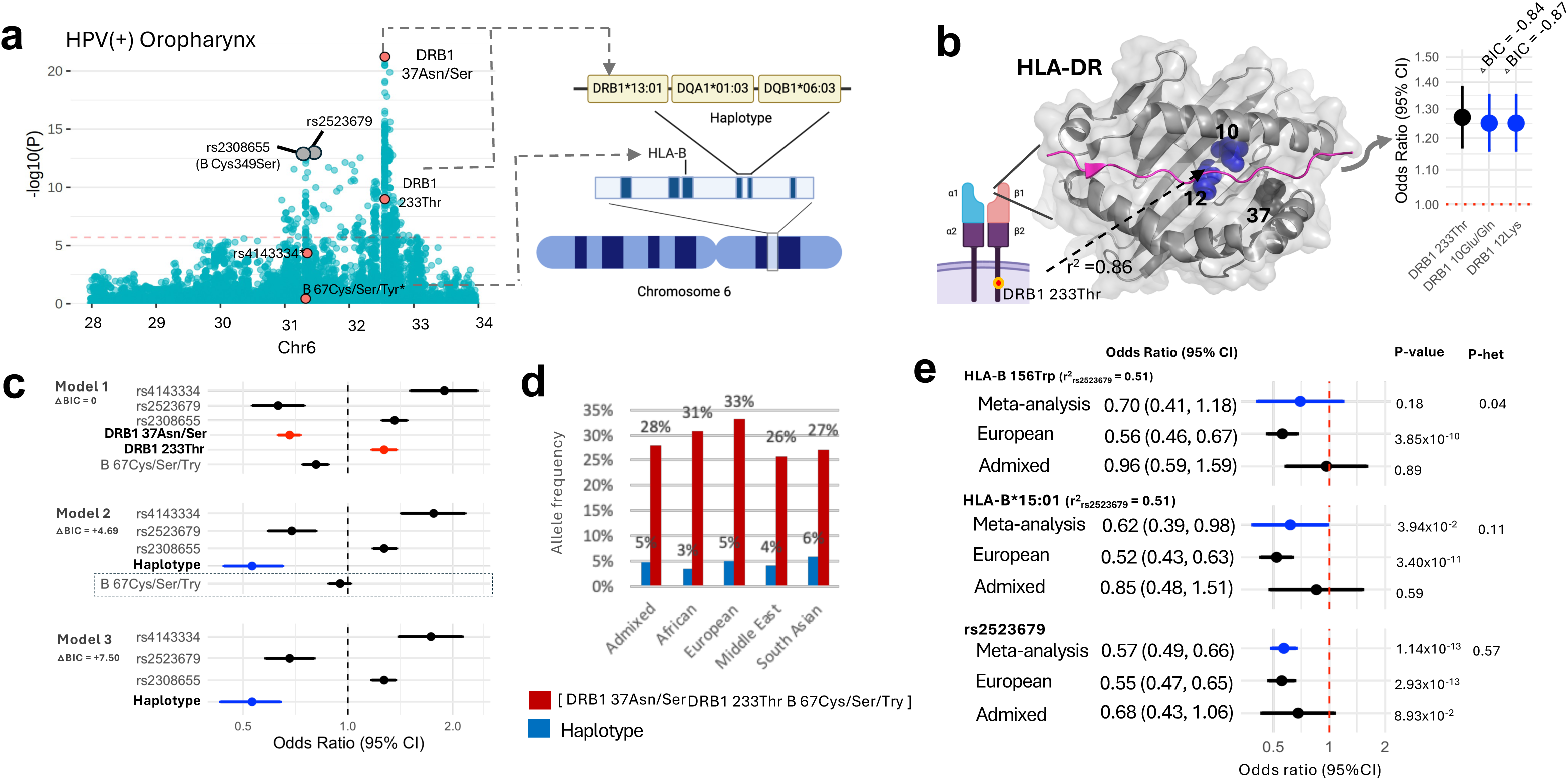
Cross-ancestry HLA risk loci of HPV(+) OPC. a) Manhattan plots showing all independent lead variants for risk of HPV(+) PC. Variants highlighted under significance threshold reached significance in later rounds; only the plot from the first round of epwise analysis is shown here. Novel variants are orange; known variants are grey. The horizontal red line reflects the HLA nificance threshold (p < 2.4 × 10^-6^). DRB1 37Asn/Ser, DRB1 233Thr, are within DRB1*13:01-DQA1*01:03-DQB1*06:03 while HLA-7Cys/Ser/Try was associated with the haplotype. b) Out of the five interchangeable amino acid residues in LD with DRB1 233Thr th △BIC ±2, DRB1 12Lys and DRB1 10Glu/Gln are in the HLA-DR binding pocket. c) Model accuracy and risk estimates of amino id residues and haplotypes. Model 1: identified from fine-mapping, used as the baseline reference model; Model 2: replaces DRB1 Asn/Ser, DRB1 233Thr with the haplotype, effect of HLA-B 67Cys/Ser/Try disappears; Model 3: All 3 amino acids replaced with plotype. d) Allele frequencies of DRB1*13:01-DQA1*01:03-DQB1*06:03 and of having all three amino acid residues by ancestry. e) e HLA-B 156Trp amino acid change and the HLA-B 15:01 allele are specific to European ancestry, but rs2523679 variant which is in with both, has a cross-ancestral effect.

For HPV(-) OPC, rs1131212 was found to be associated with an increased risk (OR (95%CI)=1.33 (1.19, 1.49), p_meta_=5.33×10^-7^) (Figure 5a). This novel, functional variant maps to exon 2 of the *HLA-B* gene causing an amino acid change Gln94His in an *HLA-B* binding pocket. rs1131212 tags another functional HLA-B amino acid change, HLA-B 70Asn/Ser (Table S12) in strong LD (r^2^=1) which has a similar effect and with similar model accuracy (OR (95%CI)=1.32 (1.18, 1.47), p_meta_=8.81×10^-7^) (Figure 5b). These results suggest that presence of either rs1131212 or HLA-B 70Asn/Ser is equivocal to increase cancer risk.

**Figure 5.**
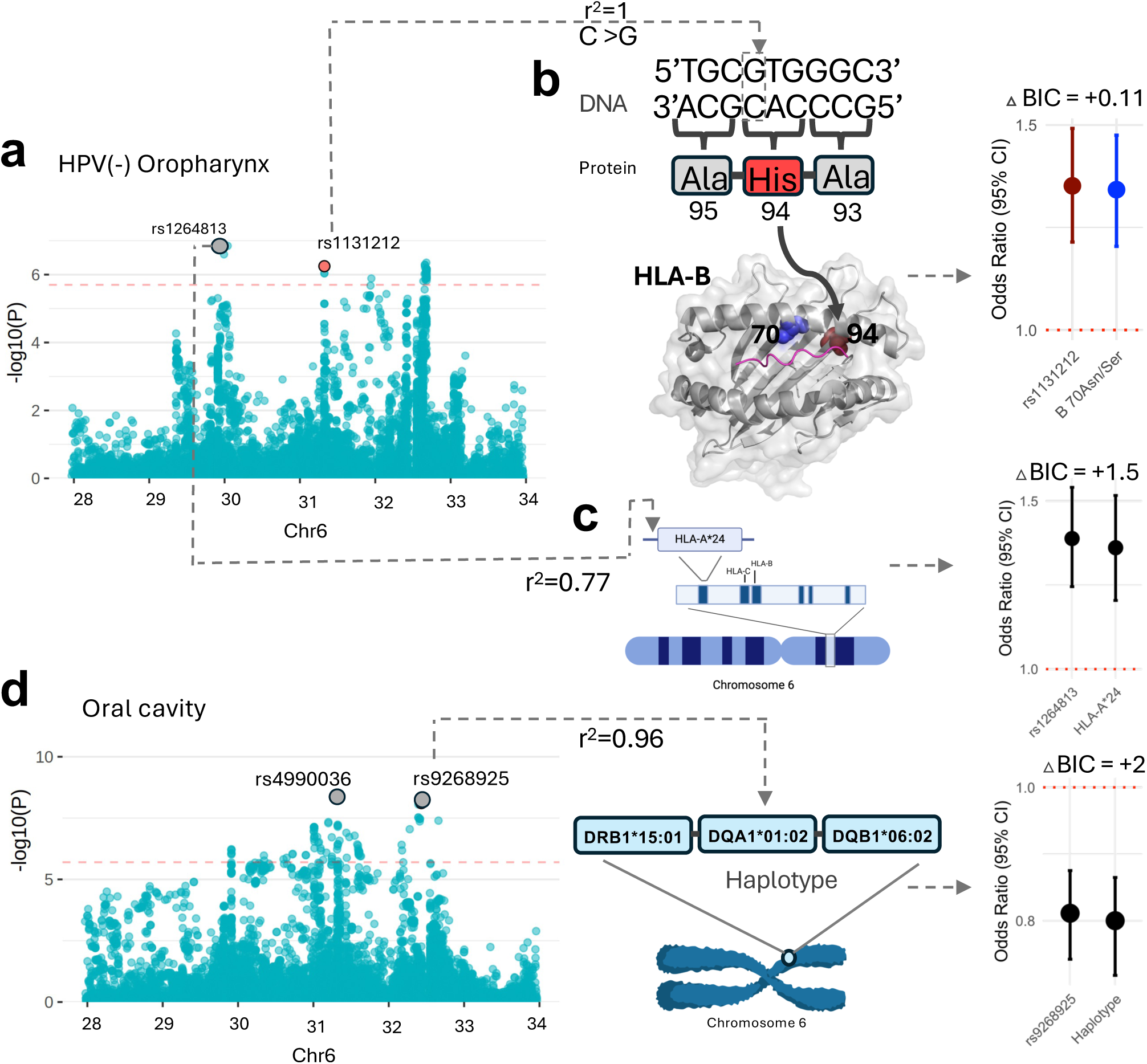
Novel HLA risk loci for HPV(-) oropharynx and oral cavity cancer. Manhattan plots display all independent lead variants of risk for each NSCC subsite. Novel variants are highlighted in red; known variants are in grey. The horizontal red line reflects the HLA significance threshold (p < 4 × 10^-6^) a) HPV(-) oropharynx: The lead SNP, b) rs1131212, causes an amino acid change from Gln to His at residue 94 located in the HLA-B protein binding pocket (PDB ID: 2BVP). This variant is in LD (r^2^=1) with 70Asn/Ser. The right panel shows the comparable risk effects of the two related signals. The known SNP, c) rs1264813, is in high LD (r^2^=0.77) with HLA-A*24 allele and shows comparable risk effects shown in right panel. d) Oral vity: The lead SNP, rs9268925, is highly correlated with a novel risk haplotype, DRB1*15:01-DQA1*01:02-DQB1*06:02, and has a similar risk effect, shown in the right panel. Model accuracy difference (△BIC) between original model in presence of all independent lead variants and the model placing the lead variant with related amino acid residue, allele or haplotype, lower than 2 confer equivalent risk.

The novel HLA-A*24 allele tagged the known intronic variant rs1264813 in *MICD*, and was similarly associated with increased risk of HPV(-) OPC (OR (95% CI)= 1.34 (1.18, 1.52), p_meta_=7.24×10^-6^). Accuracy of the model including this allele was similar to the model including rs1264813, suggesting these signals convey similar risk (Figure 5c, Tables S12).

A novel haplotype was identified that tagged the known intronic variant, rs9268925 in *DRB9*, and was associated with decreased risk of OC (OR (95% CI)= 0.8 (0.73, 0.86), p_meta_= 2.15×10^-8^). The haplotype, DRB1*15:01-DQA1*01:02-DQB1*06:02, had a similar risk and similar model accuracy compared to the known variant, suggesting that this variant and the novel haplotype can be used interchangeable to measure this risk (Figure 5d). Two novel variants specific to the European ancestry were associated with risk of OC: DRB1 74Ala/Leu/Del (OR (95% CI)= 0.82 (0.77, 0.87), p=4.94×10^-10^) and rs9267280 (OR (95% CI)= 1.32 (1.19, 1.47), p=3.48×10^-7^).

### Cross ancestry equivalent of established risk variants, including the well-known haplotype DRB1*13:01-DQA1*01:03-DQB1*06:03

The DRB1*13:01-DQA1*01:03-DQB1*06:03 haplotype is well known to reduce risk of cervical cancer and HPV(+) OPC^10,11,28^. Notably, the two novel *DRB1* amino acid changes, DRB1 37Asn/Ser and DRB1 233Thr, described here for risk of HPV(+) OPCs are within this haplotype (Figure 4a). To determine if the haplotype is completely represented by these amino acid changes, we replaced the amino acids with the full haplotype in the risk model for HPV(+) OPC (Figure 4c). Unexpectedly, the effect of HLA-B 67Cys/Ser/Tyr disappeared when including the haplotype, suggesting these are shared risk loci. When all three variants were replaced by the haplotype, the haplotype was independently associated with HPV(+) OPC risk (OR (95% CI)= 0.53 (0.43, 0.63), p_meta_=1.76×10^-10^), as described previously^11^. Importantly, model accuracy was highest for the model consisting of the original three amino acid changes compared to the haplotype, suggesting that the specific independent effects of the newly identified DRB1 37Asn/Ser, DRB1 233Thr, and possibly HLA-B 67Cys/Ser/Tyr underlie the effect of the DRB1*13:01-DQA1*01:03-DQB1*06:03 haplotype. The importance of these amino acid changes is highlighted by their allele frequencies across populations, compared to the haplotype (Figure 4d). The allele frequency of the haplotype across genetic ancestries is low and ranges from 3% to 6%, while the frequency of the three amino acids across ancestries is much higher, ranging from 26% to 33%.

The novel rs2523679 variant, which decreases risk of HPV(+) OPC (OR (95% CI) = 0.63 (0.53, 0.75), p_meta_=2.26×10^-7^), tags the established HLA-B*15:01 (r^2^=0.51) and HLA-B 156Trp (r^2^=0.51) signals that were previously found in those of European ancestry. Here we show that while the effects of HLA-B*15:01 and HLA-B 156Trp remain specific to European ancestry, rs2523679 confers a similar level of risk for both European and admixed populations, providing a cross-ancestral equivalent of this loci (Figure 4e). Other cross ancestral validated loci are described in Tables S10 and S11.

## Discussion

Across the GWAS and HLA focused analyses, we identify 29 novel variants associated with risk of HNSCC. Due to increased power compared to previous GWA studies, we identified novel genetic variants including in *TP53 and STING1* and validated known variants in *BRCA2* separately in LA and HPC, two under-studied cancer sites, as well as multiple novel signals in HPC, such as *GDF7*. Novel variants from fine mapping highlight key differences in HLA associations between HPV(+) OPCs, HPV(-) OPCs and OCs. Post-GWAS analyses, including colocalisation and the use of harmonized individual level risk factor data enabled the investigation of variant function and variant-environment interactions.

A key finding was the identification of the low frequency rs78378222 variant located in the 3′ UTR of *TP53.* This variant is an eQTL for *TP53* with the variant causing decreased expression in blood and a protective effect against overall HNSCC. This finding supports a previous candidate SNP study in a non-Hispanic white population assessing its effect on HNSCCs (OR=0.44, 95% CI: 0.24,0.79, p=0.008)^29^. Interestingly, while this variant is protective for HNSCCs and breast cancers^30^, it increases risk of skin basal cell carcinoma^21^, brain tumours,^21^ colorectal adenocarcinoma^21^, esophageal SCC,^31^ prostate cancers,^21^ and neuroblastoma^32^. Mouse models developed by Deng and colleagues have demonstrated these contrasting effects^30^. Mice with the variant exhibited reduced tumor formation and increased survival rates for breast cancers but showed the opposite trend for gliomas. The authors demonstrated the rs78378222 variant compromises the microRNA (miR)-325 target site but creates a miR-382 target site in the p53 mRNA. Both these microRNA molecules downregulate p53 but have differential tissue expressions. miR-325 expression is high in breast tissue meaning the variant p53 can escape the miR-325 downregulation, but brain tissue has high miR-382 expression, meaning the variant p53 has increased downregulation. Future work should investigate the effect of miR-325 regulation of p53 in head and neck tissues of carriers of rs78378222.

Two closely linked genetic variants were identified in 5q31, including the missense variant rs1131769 found in the cyclic dinucleotide (CDN) binding domain of *TMEM173* gene, of which the resultant STING1 protein detects viral DNA and bacterial CDNs to activate the host immune response in humans. Notably, this variant shows no association with HPV(+) OPC, but a consistent increased risk for all non-HPV cancer types. Both variants also showed evidence of eQTLs for *CTNNA1,* a gene in which germline genetic variants are known to cause Hereditary Diffuse Gastric Cancer^33^.

We were able to validate several known HNSCC risk variants and further investigate their interaction with major risk factors. rs11571833 has been linked with lung and upper aerodigestive tract cancers^34^; here we demonstrate this effect is largest in LA and HPC cancers. This variant, found in *BRCA2,* causes a 93 amino-acid deletion including the RAD51 binding domain, important in the Fanconi Anaemia Pathway for double strand DNA repair, and is distinct to the highly penetrant familial *BRCA* mutations^35^. Previous literature suggests smoking is mainly implicated in the mechanism of action of rs11571833.^27^ However, here we provide evidence cross ancestry and separately in the European and Mixed ancestry groups that this variant increases HPV-negative cancer risk with either the exposure of smoking or drinking, and that there is no effect in never-smoking non-drinkers. This supports the theory that DNA repair to environmental factors is disrupted^35^ and suggests the crucial DNA damage in HNSCC can be contributed to by alcohol use or smoking. In similar analyses, we show the well-known *ADH1B* variant rs1229984, confers a protective effect for OC which is strongest in non-drinking smokers, suggesting a mechanism through smoking as well as alcohol use. The *CHRNA5* variant, rs58365910, was identified as a suggestive association for risk of LA cancer. As expected, this variant only shows an effect in smokers, suggesting that it acts through its known effect on smoking heaviness.^36^

Through HLA fine mapping efforts, we identified 11 novel loci specific to the HLA region, of which eight were separately associated with risk of OC, HPV(+) OPC or HPV(-) OPC. Most of the class I loci were found in *HLA-B*, while most the class II loci were in *DRB1*. Given the dense, overlapping structure of the HLA region, we also identified functionally equivalent signals at the amino acid, allele, or haplotype level, enabling these data to support a variety of downstream applications requiring functional information.

A novel class II haplotype was identified for risk of OC, DRB1*15:01-DQA1*01:02-DQB1*06:02. This haplotype has been found to reduce autoantibody development and abnormalities of metabolic traits, such as dysglycemia. As such, this haplotype was found to be protective against progression of type I diabetes (DM)^37^. The relevance of this finding is evidenced by a meta-analysis that found that individuals with DM have a higher risk of developing oral cancer^38^, potentially related to DM-related metabolic traits such as hypertension and dyslipidemia^39^. Nevertheless, a link between DM and OC remains inconsistent^40–42^. The OC-specific validated variant, rs4990036, is also associated with a non-HPV infection, varicella zoster^43^, highlighting that other infections may be important in cancer risk. This is especially important considering the oral microbiome as a potential emerging risk factor for oral cavity cancer.

The well-known haplotype, DRB1*13:01-DQA1*01:03-DQB1*06:03, has been found to be protective against cervical cancer and HPV(+) OPC, highlighting its role in detecting HPV infection ^10,11,28^. This haplotype is present at about 5% in the European ancestry and less common in other ancestries. We show here that the DRB1*13:01-DQA1*01:03-DQB1*06:03 haplotype is represented by the novel three novel amino acid changes identified in this work, DRB1 37Asn/Ser, DRB1 233Thr, and HLA-B 67Cys/Ser/Tyr. Notably however, the amino acid changes themselves more precisely estimate risk of HPV(+) OPC across ancestries and likely drive the effect of the haplotype across ancestries. The higher allele frequencies of the amino acids, ranging from 26% to 33%, allows for better detection of risk for HPV(+) OPC across populations and might be easier to incorporate into screening strategies at the population level.

The novel intronic rs2523679 variant is a cross-ancestral equivalent of HLA-B*15:01 and HLA-B 156Trp, two previously identified European-specific variants. This novel variant can now be used to evaluate risk of HPV(+) OPC across multiple ancestries, and highlights the importance of including non-European populations, even with limited sample size.

In this work, we were limited by the power from non-European populations, forcing us to combine multiple populations. Although this did provide additional power for discovery, it will have reduced the ability to identify variants specific to certain populations. Where variants were specific to non-European ancestries, we were able to assess these in the different populations but increased sample sizes from more diverse populations should still be seen as a priority in this field.

Although analysing all-site HNSCC can be beneficial, it must be remembered that these cancers are heterogeneous, and the subsite analyses provide a clearer picture of the genetic architecture of the conditions. Where we identify genetic variants in one site, we assess the effect of this variant across all subsites to assess the heterogeneity but despite the increased sample size in this study, there may still be limited power for discovery, especially in the less common subsites such as HPC.

In summary, in this HNSCC GWAS which includes diverse populations, we identify 29 novel genetic variants associated with HNSCC and its subsites, including rs78378222 in the *TP53* 3′ UTR which confers a 40% reduction in odds of developing overall HNSCC. We expand knowledge of the gene-environment relationship of *BRCA2* and *ADH1B* variants demonstrating their effects act through both smoking and alcohol use. Finally, through analyses focused on the HLA region, we highlight that although HPV(+) OPC, HPV(-) OPC and OC all show GWAS signal at 6p21, each subsite has distinct associations at the variant, amino acid and 4-digit allele level.

## Data Availability Statement

The full GWAS summary statistics generated from our meta-analyses will be made publicly accessible on (https://gwas.mrcieu.ac.uk/).

Additional datasets analyzed in this study are accessible through dbGaP (https://www.ncbi.nlm.nih.gov/gap/) as follows:

OncoArray Consortium - Lung Cancer Studies (dbGaP Study Accession: phs001273.v4.p2), OncoArray: Oral and Pharynx Cancer (dbGaP Study Accession: phs001202.v2.p1), National Cancer Institute (NCI) Head and Neck Cancer Study conducted on the HumanOmniExpress-12v1.0 array (dbGaP Study Accession: phs001173.v1.p1), Genome-Wide Association Study of Oral Cavity, Pharynx, and Larynx Cancers in European, North, and South American populations (dbGaP Study Accession: phs002503.v1.p1).

Data from the UK Biobank and ALSPAC consortium, are available through their respective access protocols.

## Code availability statement

This study did not employ any custom code. Instead, it utilized publicly available software tools for genetic analyses, which are cited throughout the manuscript and reporting summary.

## Acknowledgements and funding

This study was funded in part by the European Union’s Horizon 2020 research and innovation program under grant agreement No 825771 (HEADSpAcE project) and by the US National Institute of Dental and Craniofacial Research (NIDCR) grants R03DE030257 and R01DE025712. Genotyping using the Oncoarray and the All of Us array was performed at Center for Inherited Disease (CIDR) and funded by NIDCR 1X01HG007780-0 and jointly by NIDCR/NCI X01HG010743.

This publication presents data from the Head and Neck 5000 study. The study was a component of independent research funded by the National Institute for Health and Care Research (NIHR) under its Programme Grants for Applied Research scheme (RP-PG-0707-10034). The views expressed in this publication are those of the author(s) and not necessarily those of the NHS, the NIHR or the Department of Health. Core funding was also provided through awards from Above and Beyond, University Hospitals Bristol and Weston Research Capability Funding and the NIHR Senior Investigator award to Professor Andy Ness. Round 1 genotyping was funded by US National Institute of Dental and Craniofacial Research (NIDCR) grant 1X01HG007780-0. Round 2 genotyping was funded by World Cancer Research Fund Pilot Grant (grant number: 2018/1792), Above and Beyond Charity (GA2500), Wellcome Trust Research Training Fellowship (201237/Z/16/Z) and Cancer Research UK Programme Grant, the Integrative Cancer Epidemiology Programme (grant number: C18281/A19169). This latter grant also supported Human papillomavirus (HPV) serology. This research has been conducted using the UK Biobank Resource under Application Number 40644. The work of Dr. Polesel is partially supported by Italian Ministry of Health ‘Ricerca Corrente’.

The University of Pittsburgh head and neck cancer case-control study is supported by US National Institutes of Health grants P50CA097190, P30CA047904 and R01DE025712.

Geoffrey Liu is the M. Qasim Choksi Research Chair in Translational Research at University Health Network and University of Toronto and is supported by the Princess Margaret Head and Neck Translational Program, which is supported by philanthropic funds from the Wharton Family, Joe’s Team, Gordon Tozer, Reed Fund, and the Riley Family.

The University of North Carolina studies were supported in part by grants CA61188 and CA90731 from the National Institutes of Health.

Northern Cancer Foundation (Principal Investigator Grants to MSC Conlon, DP Saunders).

Rayjean J. Hung is the CIHR Canada Research Chair and the study is supported by the Canadian Cancer Society and Canadian Institute of Health Research.

The authors would like to thank all the patients, and their families involved in these studies.

Where members are identified as personnel of the International Agency for Research on Cancer/ World Health Organization, the authors alone are responsible for the views expressed in this article and they do not necessarily represent the decisions, policy or views of the International Agency for Research on Cancer / World Health Organization.

## Online Methods

### Study Design and Populations

Individual level data came from 18 studies across 23 countries in Europe, Middle East, North America, South America, and South Asia, and 9 genotyping arrays (Table S1). Informed consent and ethical approval for genotyping was obtained under each individual study. An overall ethical approval for this analysis was obtained from the IARC Ethics Committee (IEC, 19-38). Data on demographics (sex, age, country), diagnosis (TNM status, year of diagnosis, ICD code -7th edition), HPV status (HPV16E6 serology, P16 immunohistochemistry (IHC), and HPV DNA in situ hybridisation (ISH)) and self-reported behaviors (smoking status, packyears, and drinking status) were collated and harmonized across all study participants. Eligible sites for inclusion consisted of the oral cavity (C00.3, C00.4, C00.5, C00.6, C00.8, C00.9, C02.0– C02.9 (except C02.4 and C02.8), C03.0–C03.9, C04.0–C04.9, C05.0–C06 (except C05.1,

C05.2)); oropharynx (C01-C01.9, C02.4, C05.1, C05.2, and C09.0–C10.9); hypopharynx (C12.0-C13.0); larynx (C32); and unknown primary site/overlapping/not otherwise specified (NOS) sites (C14, C05.8, C02.8, C76.0). Base of tongue (C01) and tonsils (C09) were grouped with oropharynx as these sites are frequently driven by HPV16. For studies with available information on HPV infection for OPC tumors, the HPV status provided by the centre was used (P16 status, HPV DNA ISH, or HPV serology). When information from various methods was available, a positive HPV status was determined by the presence of the HPV16 E6 antibody in serology. If serology data were absent, dual positivity of p16 and HPV DNA ISH was classified as HPV positive (HPV(+)), while dual negativity of p16 and HPV DNA ISH was classified as HPV negative (HPV(-)). Any other combinations of test results were considered as “not available”^44^.

Nineteen studies were included here with either multi-center case-control, cohort, or clinical trial study designs. Previously generated data was either downloaded from dbGap, requested through controlled access from relevant consortia, or contributed by the study PI and contributed 10,404 cases and 34,596 controls. New genotyping data was generated for 8,669 cases and 4261 controls and were not included in any previous GWAS. All study details, including data sources, dbGap accession numbers and case control distributions across subsites can be found in Table S1.

### Genotype quality control and imputation

A flow diagram detailing the preparation of the genetic data can be found in the supplementary material (Figure S10). Genotypes were generated from nine different genotyping arrays (Table S1). All newly generated genotype data was called using GenomeStudio (Illumina, 2014). Quality control steps were conducted within each array. All genotype data were converted to genome build 38, using the LiftOver program (https://genome.ucsc.edu/cgi-bin/hgLiftOver) to convert from previous builds. Genotype data was checked and corrected for consistency of strand, positions and reference alleles. Quality control was conducted using the PlinkQC package^45^ in R, utilising PLINK 1.9^46^. Samples were filtered for sex mismatch (males with SNP sex <0.8; females with SNP sex >0.2), missingness (>3%), heterozygosity (>3 standard deviations from mean) and cryptic relatedness (identity-by-descent > 0.185). Variants were filtered for genotype missingness (>1%), deviation from Hardy Weinberg equilibrium (p <1×10^-5^) and minor allele count (<20). The number of samples and variants removed at each QC step is provided in Table S2 and Table S13. All arrays were imputed to the TOPMED imputation panel^47^ separately using the TOPMED Imputation server^48^.

To increase the number of controls comparable to the participants in the HN5000 study, 17,815 additional participants (including known related individuals) were included from the Avon Longitudinal Study of Parents and Children (ALSPAC), which had been previously genotyped (Table S1)^49,50^. To account for potential batch effects between the HN5000 study (Infinium Global Screening Array [GSA]) and additional ALSPAC controls (Illumina 550 Quad, Illumina 660W Quad), a double imputation approach was applied (Supplementary Note 1). Briefly, GSA HN5000 cases and the additional controls were imputed to the TOPMED reference panel separately as detailed above. Following this step, variants which were (i) genotyped on both arrays, (ii) genotyped on the GSA with high quality imputation (R^2^ score >0.9) on the ALSPAC array and, (iii) genotyped on the ALSPAC array with high quality imputation (R^2^ score >0.9) on the GSA were selected. These variants were merged across the two arrays, converted to ‘best-guess’ genotypes and then included in a second joint imputation to the TOPMED reference panel. This method allowed high quality imputation of both datasets. To address concerns about batch effects between cases and controls genotyped separately, 405 ALSPAC controls were also genotyped on the GSA alongside the HN5000 cases. This enabled sensitivity analyses to account for potential batch effects.

### Genetic Ancestry stratification

Following the imputation process, markers from each imputation batch were filtered based on an imputation score of R^2^ >0.8 and merged across imputation batch and chromosome. Markers were filtered for a call rate ≥ 0.98 and minor allele frequency (MAF) ≥ 1%. The major histocompatibility complex (MHC) region was removed, and the remaining markers were pruned for independent variants using linkage disequilibrium (LD) with a squared correlation (r²) threshold of < 0.2. This set of markers (N=697,099) was utilized to compute kinship estimates between Individuals using the KING-robust kinship estimator^51^ in PLINK 2.0^46^. The KING-robust method is specifically designed to be robust to population structure and admixture. It calculates kinship coefficients without being biased by the fact that certain populations may have different allele frequencies. In addition to the removal of 6,679 known related individuals from the ALSPAC study, a kinship cutoff of > 0.0884 was applied to exclude unexpected duplicates and individuals related at the second degree or closer. This cutoff is based on the geometric mean of the theoretical values for second- and third-degree kinship, as outlined in the manual. Selection of related individuals or duplicates were prioritized based on either disease status (favouring cases over control) or array type (favouring newer arrays over older ones). After this process, 3,441 individuals were excluded from the analysis. The remaining 58,625 individuals were classified into genetic ancestries using supervised ADMIXTURE analysis (ADMIXTURE 1.3^52^) with 75,164 common markers retained after quality control steps (Figure S11). This assigns a percentage probability for belonging to each of the reference super-populations in the 1000 Genomes Project (N=2,504)^53^. We assigned individuals to a dominant genetic ancestry if their probability was ≥70% to any reference super-population. Of all individuals, 48,029 (83%) had a dominant genetic ancestry while the remainder were classified as admixed. The distribution of individuals with a dominant genetic ancestry was as follows: 80.2% European (EUR), 0.1% Admixed Americans (AMR), 1.2% Africans (AFR), 1.3% South Asians (SAS), and 0.2% East Asians (EAS). The remaining 17% were not able to be classified with a dominant genetic ancestry and were grouped as “admixed”. To improve statistical power to detect risk loci across the relatively small sample sizes of non-European genetic ancestries, all five (AMR, AFR, SAS, EAS and admixed) were merged to create a “Mixed” group (n = 11,462) (Figure S12a, S12b). Genome-Wide Association Studies (GWAS) were conducted separately in the European and Mixed ancestry samples and meta-analysed (see later). Principal Component Analysis (PCA) was carried out within each ancestral sample (European and Mixed) to assess population substructure and for covariate adjustment in GWAS (Figure S13). For HLA fine-mapping analyses, a slightly different approach was required due to the region’s high LD and highly correlated variants. Additionally, the HLA region is more susceptible to population substructure, making it challenging to identify causal variants that are consistent across ancestries. Therefore, for fine mapping, samples were grouped according to their dominant genetic ancestry (>70%) (EUR, AFR, and SAS) or admixed. Based on the homogeneous clustering identified through PCA (Figure S14), the samples from Iran were separated to Middle Eastern (ME) ancestry. Small size numbers (Case/Control < 50) of genetic ancestries (AMR and EAS) were merged into admixed. For each sample, PCA identified informative principal components (PCs) that showed significant associations (p <0.05) with case-control status after adjusting for sex and imputation batch. These informative PCs, along with sex and imputation batch, were included as model covariates in the GWAS analysis.

### Association, Meta- and Conditional Analysis

Across the 9 arrays and 19 studies, there were several considerations in how to adjust for batch effects. Some studies, such as ARCAGE, were split across different arrays, such as the Oncoarray and AllofUs array. For other studies, such as UKBiobank, several arrays were used (UKBiLEVE and AffymetricUKB) (Table S1). Finally, HN5000 and ALSPAC differed in their imputation as the ‘double imputation’ method was used. Therefore, a ‘Batch’ variable was created to represent the combination of studies, arrays and imputation approaches that could contribute to batch effects. To evaluate the potential impact of these different batches in the regression models, we conducted a sensitivity analysis by running GWAS within each batch and assessed heterogeneity using METAL^54^. We excluded markers with a heterogeneity p-value <5×10^-8^, resulting in the removal of 137 markers in the European sample GWAS.

Association analysis was conducted separately for all sites combined and for each HNSCC subsite in the European and Mixed samples using PLINK. The results were then meta-analysed with METAL^54^ using a fixed effects model to identify cross-ancestral loci. There was minimal inflation after adjustment for informative PCs in most analyses (lambda (λ) ranging from 1.00 to 1.03). However, the HPV(+) and HPV(-) OPC analyses for the Mixed group did show evidence of inflation (HPV(+) OPC: λ=1.12; HPV(-) OPC: λ=1.20) (Figure S15). Consequently, rather than a meta-analysis, the GWAS analysis for OPC was conducted only in the European sample with consistency of top SNPs assessed separately in the Mixed sample. For all other subsites, loci that achieved p<5×10^-8^ in the meta-analysis were referred to as cross-ancestral. This threshold was selected as it is equivalent to a standard Bonferonni correction for one million independent tests. Loci satisfying p<5×10^-8^ within each ancestral sample which 1) were not significant in the meta-analysis and 2) showed no attenuation upon conditional analysis of nearby lead cross-ancestral SNPs and therefore considered to be independent from the cross-ancestral SNP, were hereby referred to as ancestral-specific (Table S14). Where these existed in the Mixed ancestry sample, further stratification into the five dominant genetic ancestries was performed. Regional association plots were generated using Locus Zoom (https://my.locuszoom.org/).

### HLA fine mapping

Variants that were directly genotyped in chromosome 6 were extracted from genotyping data of all arrays and standardized to hg19 using LiftOver. Due to restrictions in data access from ALSPAC, additional data from UK Biobank was used to replace ALSPAC for double imputation with HN5000 as described above. Per variant QC was conducted by deduplication of SNP data, strand alignment, removal of palindromic variants (i.e., SNPs with A/T or G/C alleles), removal of poor-quality variants with missingness threshold of 10% and Hardy-Weinberg equilibrium threshold of 1×10^-10^. Sample QC was conducted after the removal of samples with high missingness rates, outlier heterozygosity, discordant sex information, and genetically identical samples. A flow diagram of QC steps for the HLA fine mapping is provided in Figure S16.

The HLA region (Chromosome 6:28Mb-34Mb) was imputed for SNPs and classical HLA class I and II alleles using the Michigan imputation server with the most recent HLA Multi-ethnic reference panel (Four-digit Multi-ethnic HLA v2)^55^. Only high-quality SNPs, alleles or amino-acid residues were included in the analysis (imputation r^2^>0.95). The final set of imputed variants used in association analyses were of high quality; 91% of the variants and 71% of the less common variants (MAF < 0.05) had imputation R^2^ ≥ 0.95. HLA-wide association analysis was conducted controlling for sex, informative PCs, and imputation batch (described above), and meta-analyzed with a random effect model using PLINK^46^ to identify cross-ancestral variants. Any genetic ancestries with fewer than 50 samples were excluded from meta-analyses due to power. Stepwise conditional analysis was conducted to identify independent variants within each ancestry where variants with the lowest p-value after each round were added to the subsequent model and the analysis was repeated until no further variants met the significance threshold. As HLA fine mapping was conducted independently from GWAS, a probability threshold was set to 2.4×10^-6^. This was based on the total number of imputed HLA variants (0.05/20,762), which included SNPs, amino acid variants, and classical HLA alleles after quality control as described previously ^56^.

To identify haplotypes associated with risk within each subsite that were linked to the top novel variants identified from fine mapping, the haplo.stats package v.1.9.5.1 in R was applied to identify combinations of HLA 4-digit alleles within each population. The haplo.em and haplo.glm algorithms identified haplotype candidates in each population with a minimum haplotype frequency threshold set at 0.01 in comparison to the most common haplotype within the ancestry. Haplotype candidates that were in high LD (r^2^ >0.8) with variants from fine mapping were then tested for association with risk using the meta-analysis approach to determine if they conferred similar risk compared to their variant counterparts.

### Testing for Independence and functional equivalents of lead variants

Variants identified in each HNSCC subsite analysis from the GWAS, and fine mapping were compared across subsites to evaluate whether they were linked or independent. This was also performed to define variants that were novel compared to previously reported signals and to determine overlapping signals between cross-ancestral and population-specific variants. LD was measured by r^2^ using PLINK 1.9^46^ within the overall dataset. If LD>0.3, then conditional analysis was performed to evaluate if the significance of the variant of interest attenuated to lower than 2.4×10^-6^. If both criteria were met, variants were considered to be dependent.

To determine functional equivalents of the variants identified through fine mapping, amino acid changes, alleles and haplotypes that were in moderate to high LD (r^2^>0.5) with lead novel variants were further evaluated. Effect sizes and significance levels were compared when replacing the lead variant with the related variant in the fully adjusted cross-ancestral model. Bayesian Information Criterion (BIC) were then evaluated to compare the model fit of the original model with the lead variants identified from fine mapping to the model with the related variant replacing the original lead variant. Every permutation of variants was considered to determine if one variant could replace by another and still provide the same information as the original lead variant.

### Stratified analyses

For each independent top hit identified in GWAS and HLA fine mapping, the analysis was repeated, stratified by sex, smoking status, drinking status, geographic region, and within all cancer subsites separately. The effects across strata were assessed for heterogeneity using χ^2^-based Q test (Cochran’s Q test) using R (v4.1.2). Further stratification for specific variants related to smoking and alcohol was conducted in non-HPV related cancers. This assessed effects in never-smoking non-drinkers, smoking non-drinkers, never-smoking drinkers and ever-smoking drinkers to assess the independence of these risk factors where data was available. Results were presented in forest plots (Figures S7, S9).

### Heritability and genetic correlation

SNP based heritability was estimated in the European and Mixed ancestry samples using the Genome-based restricted maximum likelihood (GREML) method in GCTA^57^. Imputed genetic data was used, variants with MAF<0.01 and Hardy Weinberg equilibrium p <0.05 were removed, as suggested for case-control data in Lee *et al*. 2011^58^. Univariate GREML was used to estimate heritability and was transformed onto the liability scale using global prevalence estimates from GLOBOCAN^1^. Heritability estimates in the Mixed ancestry sample are not presented in the main manuscript due to the heterogeneous nature of these samples which make estimates of heritability unreliable. These are provided in Table S5 for completeness.

### Colocalisation of GWAS and eQTL mapping

Colocalisation of genetic associations between all identified top hit variants (from GWAS analyses and fine mapping) and their gene expression and related traits was calculated using default LDs and a window size of ±75 kb using the COLOC package^59^. All colocalisation analyses were conducted using HNSCC data of European ancestry. Expression quantitative trait loci (eQTLs) in whole blood were obtained from the eQTLGen Consortium^60^ due to its role in immune response and systemic inflammation. eQTLs in esophagus and lung tissue were sourced from the Genotype-Tissue Expression (GTEx) project (v8)^61^, given their anatomical proximity and shared risk factors, such as tobacco and alcohol exposure. In the analyses, we considered eQTLs coinciding with genomic loci identified in this study at p< 3.9×10^-10^ in whole blood data; p< 2.5×10^-7^ in tissue data to be considered as significant). Summary statistics from GWAS for smoking and alcohol consumption behaviors were sourced from the GWAS & Sequencing Consortium on Alcohol and Nicotine Use (GSCAN) ^62^. The analysis considers the posterior probability of colocalisation for a single shared variant responsible for the associations in both traits (posterior probability for hypothesis 4 (PP4)), values over 0.7 were considered strong evidence of colocalisation. Where the lead variant was not available in the LD reference panel required for COLOC, the variant with the highest LD was used instead.

### Technical validation

For the technical validation of the imputed *TP53* variant, we utilized a Taqman assay to genotype this specific variant in a subset of samples from the Central and Eastern European Study (CEE) and ARCAGE studies. Individuals removed from the GWAS in QC steps or those with technical issues during the Taqman assays, e.g. failure to amplify, were removed resulting in 2,370 samples where consistency could be assessed. Overall concordance and non-reference discordance were calculated.

### HEADSpAcE Consortium Member Acknowledgements

Adam R^1^, Agudo A^2^, Alibhai S^3^, AlWaheidi SF^4^, Angel Pavon M^2^, Anwar N^5^, Arantes P^6^, Arguello L^7^, Avello Y^8^, Avondet L^1^, Baldión-Elorza AM^8^, Batista Daniel C^9^, Beraldi B^10^, Berenstein B^11^, Bernal P^12^, Bernardino Rodrigues N^13^, Bilic Zimmermann J^2^, Botta MG^6^, Bouvard L^4^, Brenes J^2^, Brenner N^14^, Brentisci C^15^, Burtica C^8^, Cabañas ML^16^, Cantor E^17^, Carvalho RS^18^, Carvalho A L^19^, Chiusa L^20^, Chopard P^4^, Chundriger Q^21^, Clavero O^2^, Costa I^22^, Creaney G^23^, Cuffini C^24^, Dias TC^18^, Duccini de Souza E^10^, durant l C^25^, Escallón A^26^, Fernandes G^6^, Fervers B^27^, Fiano V^28^, Firme Figueira F^13^, Furbino Villefort R^13^, Gangemi M^15^, Garzino-Demo P^29^, Gholipour M^30^, Giglio R^11^, Goulart MA^31^, Graça Sant’Anna J^9^, Grega M^32^, Gregório Có A^9^, Guasch A^2^, Hakim JA^26^, Hayes DN^33^, Homero de Sá Santos M^10^, Hurley K^34^, Insfran M^35^, Iorio GC^36^, Iqbaluddin Siddiqui M^37^, Johannsen J^38^, Kaňa M^39^, Klussmann J^38^, Legal E^40^, Lenzi J^10^, Luiz Dias F^22^, Lyra González I^41^, Machado Zorzaneli W^9^, Mai Rocha R^10^, Maños M^2^, Marinho de Abreu P^9^, Marzban M^42,43^, McCaul J^44^, McMahon AD^31^, Mena C^40^, Mendonça EF^45^, Mendoza L^35^, Meza L^35^, Michels B^14^, Mineiro MS^46^, Moccia C^28^, Mongelos P^35^, Montealegre-Páez AL^47^, Morey Cortes F^2^, Muñoz A^48^, Ness A^34^, Neves AB^6^, Oliva M^2^, Oliveira J^49^, Ortiz H^7^, Ortiz J^40^, Osorio M^40^, Ospina V^17^, Ostellino O^50^, Palau M^8^, Paterson C^51^, Paytubi Casabona S^2^, Pecorari G^29^, Pereira DM^52^, Pérol O^27^, Pervez S^21^, Pomata A^16^, Popovic M^28^, Poveda A^47^, Prado CP^6^, Prager KM^14^, Ramieri G^29^, Rasul S^3^, Rego JN^53^, Reis RM^18^, Renard H^4^, Ricardi U^36^, Riva G^29^, Rodilla F^2^, Rodriguez I^54^, Rodríguez MI^35^, Ross A^31^, Roux P^55^, Saeed Ali T^56^, Saintigny P^57^, Santivañez J J^26^, Scapultampo-Neto C^58^, Segovia J^17^, Sena A^10^, Serrano R^40^, Sharma S^38^, Siefer O^38^, Smart S^59^, Sorroche BP^18^, Sosa C^16^, Souza JD^60^, Stura A^15^, Thomas S^34^, Torres O^61^, Tous S^2^, Ucross G^12^, Valenzuela A^35^, Vasconcelos de Podestá J^10^, Whitmarsh A^34^, Wright S^62^

^1^H&N cancer Department, Universidad de Buenos Aires, Ciudad Autonoma de Buenos Aires, Argentina, ^2^Catalan Institute of Oncology (ICO), Barcelona, Spain, ^3^Department of Surgery, Dental Hygiene Program, Aga Khan University Hospital, Karachi, Pakistan, ^4^Genomic Epidemiology Branch, International Agency for Research on cancer (IARC/WHO), Lyon, France, ^5^Faculty of Science and Technology, University of Central Punjab, Lahore, Pakistan, ^6^Group of Epidemiology and Statistics on Cancer, A.C.Camargo Cancer Center, Sao Paolo, Brazil, ^7^Servicio de Cabeza y Cuello, Instituto Nacional del Cáncer, Ministerio de Salud Pública y Bienestar Social, Capiatá, Paraguay, ^8^Pathology and Laboratory Department, Fundación SantaFe de Bogotá, Bogotá, Colombia, ^9^Postgraduate Program in Biotechnology, Universidade Federal do Espirito Santo, Vitoria, Brazil, ^10^Head and Neck Surgery Division, Associação Feminina de Educação e Combateao Câncer(AFECC), Hospital Santa Rita de Cássia, Vitoria, Brazil, ^11^H&N cancer Department, Institute of Oncology Angel H. Roffo, University of Buenos Aires, Ciudad Autonoma de Buenos Aires, Argentina, ^12^Department of Radiology, Division of Nuclear Medicine, Fundación SantaFe de Bogotá, Bogotá, Colombia, ^13^Department of Pathology, Universidade Federal do Espirito Santo, Vitoria, Brazil, ^14^Division of Infections and Cancer Epidemiology, German Cancer Research Center (DKFZ), Heidelberg, Germany, ^15^Department of Medical Sciences, Cancer Epidemiology Unit, AOU Città della Salute e della Scienza di Torino, Turin, Italy, ^16^Departamento de Anatomía Patológica, Instituto Nacional del Cáncer, Ministerio de Salud Pública y Bienestar Social, Capiatá, Paraguay, ^17^Oncology Deparment, Fundación SantaFe de Bogotá, Bogotá, Colombia, ^18^Molecular Oncology Research Center, Barretos Cancer Hospital, Barretos, Brazil, ^19^Department of Head and Neck Surgery, Barretos Cancer Hospital, Barretos, Brazil, ^20^Pathology Unit, AOU Città della Salute e della Scienza di Torino, Turin, Italy, ^21^Department of Pathology and Laboratory Medicine, Section of Histopathology, Aga Khan University Hospital, Karachi, Pakistan, ^22^INCA, Rio de Janeiro, Brazil, ^23^School of Medicine, Dentistry and Nursing, University of Glasgow, Glasgow, United Kingdom, ^24^Universidad Nacional de Cordoba, Cordoba, Argentina, ^25^A.C Camargo Cancer Center, São Paulo, Brazil, ^26^Department of Surgery, Head and Neck Division, Fundación SantaFe de Bogotá, Bogotá, Colombia, ^27^Department Cancer Environnement, Centre Léon Bérard, Lyon, France, ^28^Department of Medical Sciences, Cancer Epidemiology Unit, University of Turin, Turin, Italy, ^29^Department of Surgical Sciences, University of Turin, Turin, Italy, ^30^Metabolic Disorders Research Center, Golestan University of Medical Sciences, Gorgan, Iran, ^31^School of Medicine, Dentistry and Nursing, University of Glasgow, Glasgow, United Kingdom, ^32^Department of Pathology and Molecular Medicine, 2nd Faculty of Medicine, Charles University and Motol University Hospital, University Hospital in Motol, Prague, Czech Republic, ^33^Department of Genetics, Genomics and Informatics, University of Tennessee Health Science Center, Memphis, USA, ^34^Bristol Dental School, University of Bristol, Bristol, United Kingdom, ^35^Salud Pública, Instituto de Investigaiones en Ciencias de la Salud (IICS), Universidad Nacional de Asunción (UNA), San Lorenzo, Paraguay, ^36^Department of Oncology, University of Turin, Turin, Italy, ^37^Department of Surgery, Section of E.N.T, Aga Khan University Hospital, Karachi, Pakistan, ^38^Department of Otorhinolaryngology, Head and Neck Surgery, University of Cologne, Cologne, Germany, ^39^Departrment of Otorhinolaryngology and Head and Neck Surgery, University Hospital in Motol, Prague, Czech Republic, ^40^Cátedra Otorrinonaringología, Hospital de Clínicas, Facultad de Ciencias Médicas, Universidad Nacional de Asunción, San Lorenzo, Paraguay, ^41^Servicio de Oncología Clínica Hospital de Clínicas, Universidad de la República, Montevideo, Uruguay, ^42^The Persian Gulf Tropical Medicine Research Center, The Persian Gulf Biomedical Sciences Research Institute, Bushehr University of Medical Sciences, Bushehr, Iran, ^43^Statistics Genetic Lab, QIMR, Berghofer Medical Research Institute, Brisbane, Australia, ^44^Department of Oral and Maxillofacial/Head and Neck Surgery, NHS Greater Glasgow and Clyde, Glasgow, United Kingdom, ^45^Hospital Câncer Araújo Jorge, Goiânia, Brazil, ^46^Hospital Câncer Araújo jorge, Goiânia, Brazil, ^47^Faculty of Medicine, El Bosque University, Bogotá, Colombia, ^48^Oncology Department, Division of Radiotherapy, Fundación SantaFe de Bogotá, Bogotá, Colombia, ^49^Goiânia Cancer Registry (BR), Goiânia, Brazil, ^50^Department of Oncology, Division of Medical Oncology, AOU Città della Salute e della Scienza di Torino, Turin, Italy, ^51^Beatson West of Scotland Cancer Centre, NHS Greater Glasgow and Clyde, Glasgow, United Kingdom, ^52^Radiation Oncology Department, Institute of Oncology Angel H. Roffo, University of Buenos Aires, Ciudad Autonoma de Buenos Aires, Argentina, ^53^Clinical Research Center, Associação Feminina de Educação e Combateao Câncer(AFECC), Hospital Santa Rita de Cássia, Vitoria, Brazil, ^54^Laboratorio de Anatomía Patológica, Hospital de Clínicas, Facultad de Ciencias Médicas, Universidad Nacional de Asunción, San Lorenzo, Paraguay, ^55^Department of Surgery, Centre Léon Bérard, Lyon, France, ^56^School of Nursing and Midwifery, Aga Khan University Hospital, Karachi, Pakistan, ^57^Centre Léon Bérard, Lyon, France, ^58^Pathology and Molecular Diagnostics Service, Barretos Cancer Hospital, Barretos, Brazil, ^59^University of Glasgow, Glasgow, United Kingdom, ^60^Epidemiology and Statistics Group, Research Center, A.C Camargo Cancer Center, São Paulo, Brazil, ^61^Radiology Department, Fundación SantaFe de Bogotá, Bogotá, Colombia, ^62^Institute of Cancer Sciences, University of Glasgow, Glasgow, United Kingdom.

