## Supplementary Figures for "Cross-ancestral GWAS identifies 29 novel variants across Head and Neck Cancer subsites"

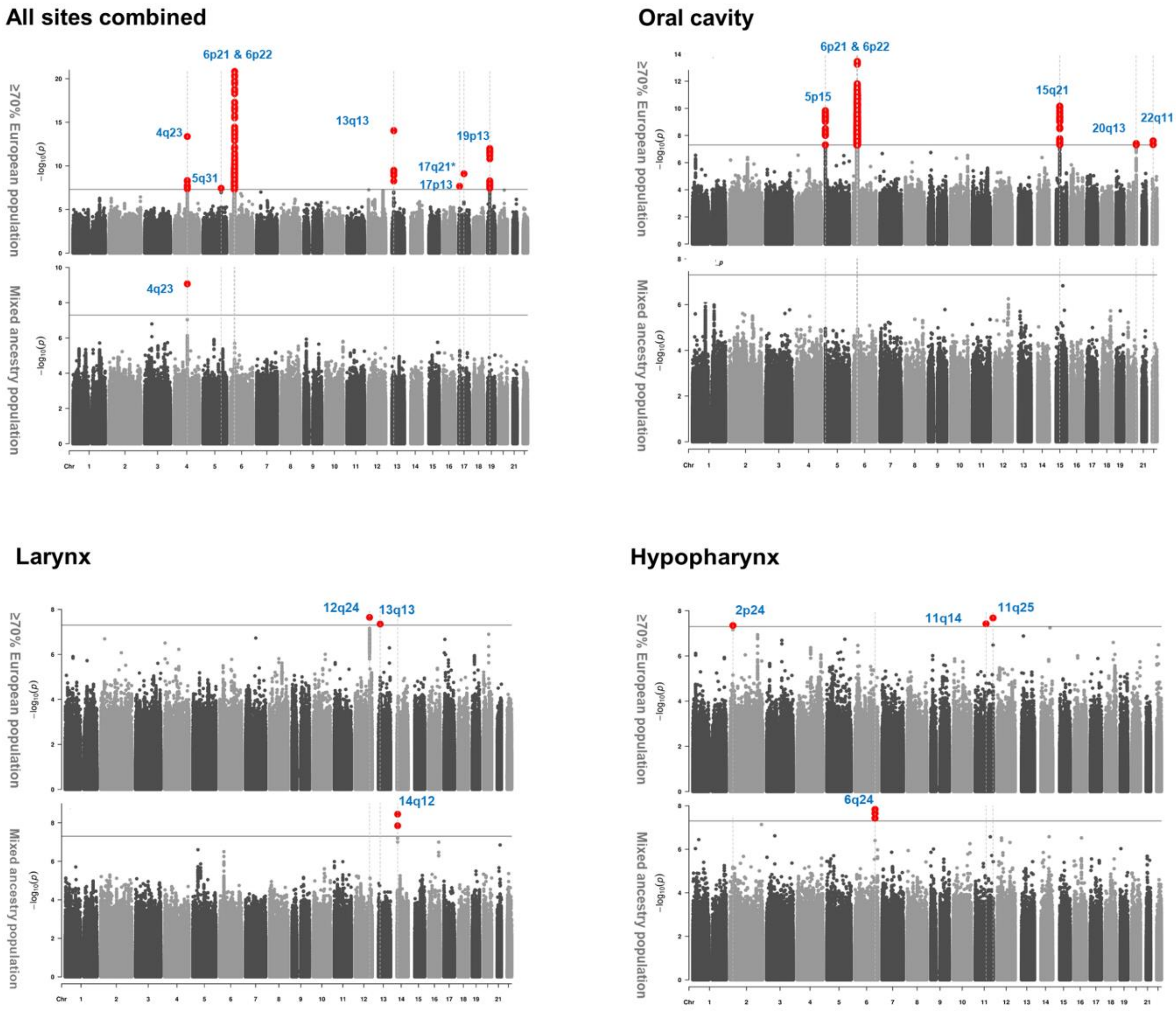


**Supplementary Figure 1. Genome-wide association results for overall HNSCC, oral cavity, larynx and hypopharynx in European and mixed ancestry groups.** The grey line indicates the genome-wide significance threshold of $5x10^{-8}$. Variants that met or exceeded this threshold are highlighted in red.

*In All sites combined the 17q21 variant was excluded due statistically significant heterogeneity across imputation batches/studies.

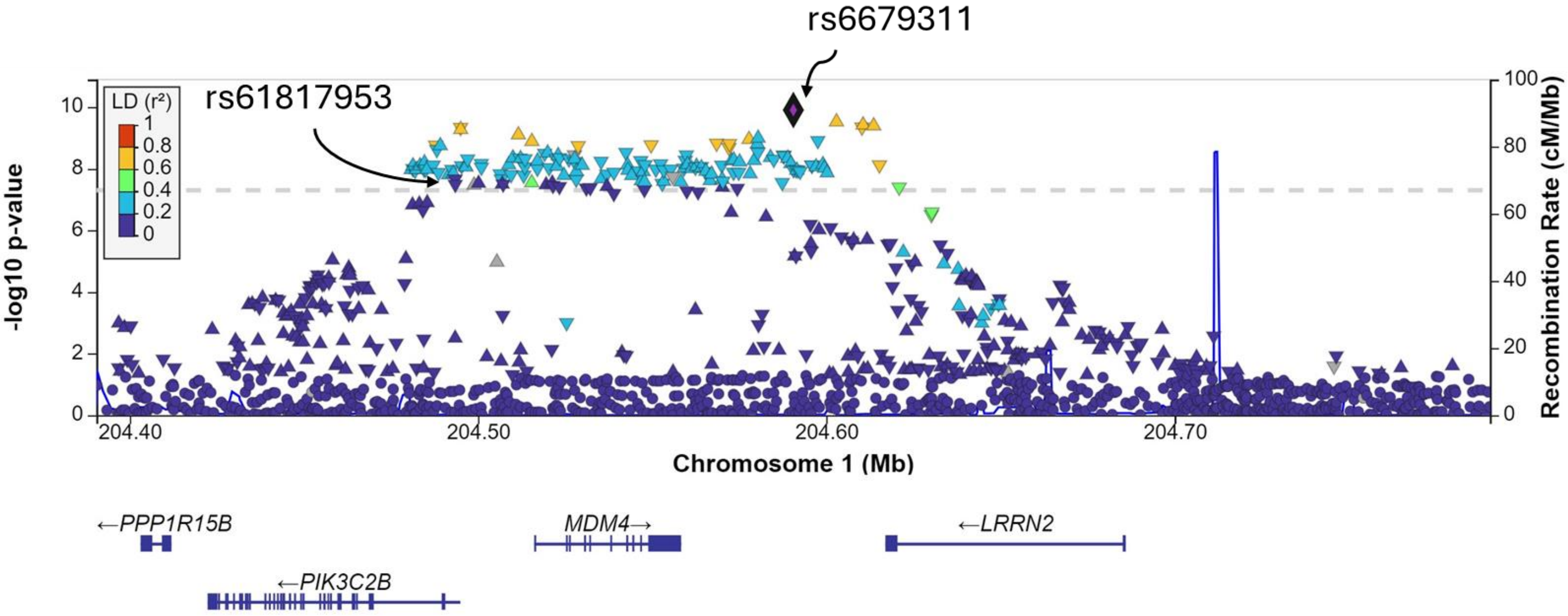


**Supplementary Figure 2. Regional plot for two independent variants, rs61817953 and rs6679311, identified at 1q32.** The x-axis represents the chromosome position, while the y-axis shows the –log10 P value. rs61817953, near *PIK3C2B* (OR (95%CI) = 0.90 (0.87, 0.93), $p_{meta}$= 2.17x$10^{-8}$) and rs6679311 near *MDM4,* a strong negative regulator of *TP53* (OR (95% CI) = 1.11 (1.07,1.14),$p_{meta}$=1.25x$10^{-10}$).

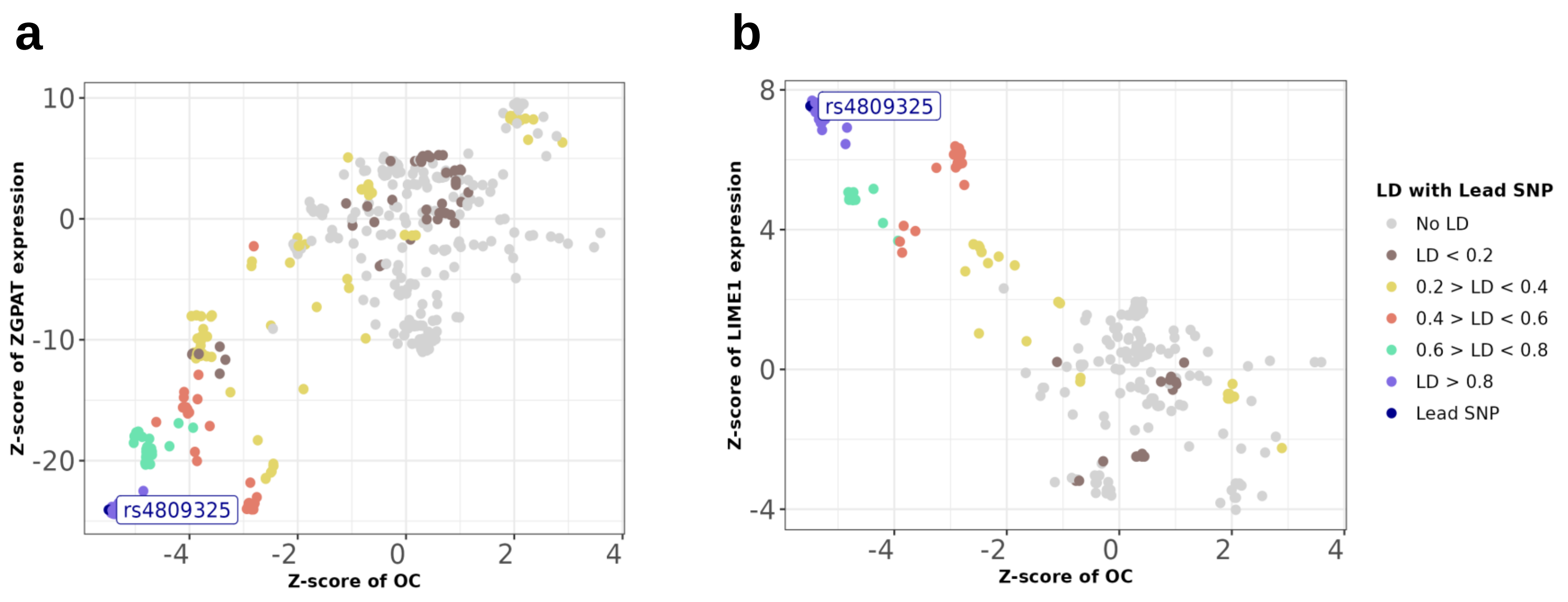


**Supplementary Figure 3. Z-Z locus plot of rs4809325.** a) rs4809325, colocalised with Zinc Finger CCCH-Type And G-Patch Domain Containing (*ZGPAT*) in whole blood (PP4 score= 0.97) and b) *LIME1* in esophagus mucosa (PP4 score=0.97); OC, Oral cavity.

**a**

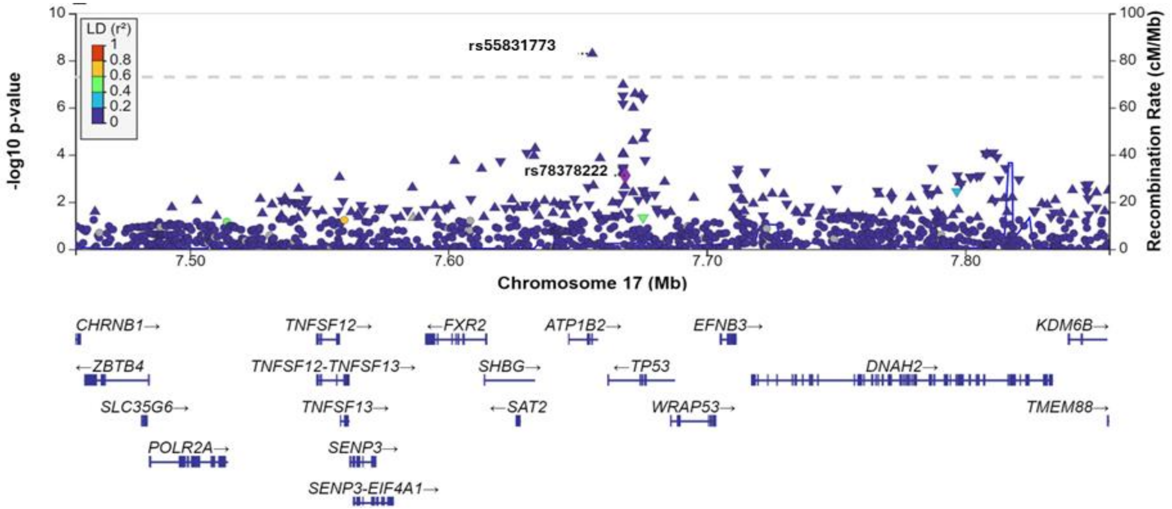


**b**

| Subsite | p-value before conditioning | p-value after conditioning |
|---|---|---|
| **All sites combined (European ancestry group)** | | |
| rs55831773 | 0.0004 | 0.001 |
| rs78378222 | 2.16E-08 | 2.16E-08 |
| **Larynx (European ancestry group)** | | |
| rs55831773 | 2.16E-07 | 6.23E-07 |
| rs78378222 | 0.0007 | 0.0007 |

**c**

| Variable | NO.Case | NO.Control | OR.(95% Conf. Int.) | P-Heterogeneity |
|---|---|---|---|---|
| **Cross-ancestry meta-analysis** | | | | |
| Larynx | 4409 | 37722 | 1.21(1.13 to 1.29) | |
| **Genetic ancestry** | | | | |
| European | 2787 | 31833 | 1.23(1.14 to 1.32) | 0.53 |
| Mixed | 1622 | 5889 | 1.17(1.05 to 1.31) | |
| **Sex** | | | | |
| Men | 3826 | 19246 | 1.22(1.14 to 1.31) | 0.33 |
| Women | 583 | 18476 | 1.12(0.96 to 1.31) | |
| **Smoking Status** | | | | |
| Never | 552 | 15197 | 1.05(0.86 to 1.29) | 0.21 |
| Former | 1726 | 9741 | 1.29(1.15 to 1.44) | |
| Current | 2114 | 5675 | 1.26(1.12 to 1.41) | |
| **Drinking Status** | | | | |
| Never | 774 | 3398 | 1.13(0.96 to 1.33) | 0.26 |
| Ever | 3366 | 26338 | 1.26(1.16 to 1.36) | |
| **Geographic Region** | | | | |
| Europe | 2003 | 30967 | 1.17(1.08 to 1.27) | 0.7 |
| North America | 1354 | 3694 | 1.15(1.03 to 1.30) | |
| South America | 739 | 2239 | 1.17(1.00 to 1.37) | |
| Middle East | 282 | 504 | 1.00(0.79 to 1.28) | |
| **Subsite** | | | | |
| Oral cavity | 5596 | 37722 | 1.02(0.96 to 1.08) | 0.97 |
| Hypopharynx | 898 | 37722 | 1.02(0.90 to 1.15) | |
| Oropharynx (HPV-) | 1473 | 23132 | 1.00(0.91 to 1.11) | |
| Oropharynx (HPV+) | 2212 | 22803 | 0.99(0.91 to 1.08) | |

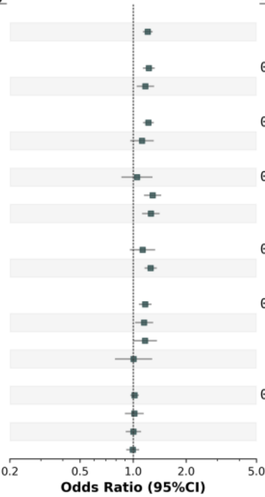


**Supplementary Figure 4. Regional plot of rs55831773 identified at 17p13.** a) rs55831773, a splice polypyrimidine tract variant, mapped to *ATP1B2* was associated with increased risk of laryngeal cancer in the cross-ancestral analysis (OR (95% CI)=1.21 (1.13,1.29), $p_{meta}$=5.1x10$^{-9}$). b) *ATP1B2* is in close proximity to *TP53* but conditional analyses (performed in European population) confirm this variant is independent of rs78378222 (the rare *TP53* 3'-UTR variant) identified in overall HNSCC in European population. c) forest plot of Odds ratio for rs55831773 variant stratified by sex, smoking- and drinking status, and geographic region within cross-ancestry laryngeal cancer meta-analysis. The risk-increasing effect of this variant was observed only in laryngeal cancer, with no effect on other subsites. In contrast, the *TP53* variant rs78378222 showed a protective effect in non-HPV-related HNSCC cancer sites.

| Variable | NO.Case | NO.Control | OR.(95% Conf. Int.) | P-Heterogeneity |
|---|---|---|---|---|
| **Cross-ancestry meta-analysis** | | | | |
| Larynx | 4409 | 37722 | 1.25(1.00 to 1.55) | |
| **Genetic ancestry** | | | | |
| European | 2787 | 31833 | 0.82(0.63 to 1.07) | < 0.0001*** |
| Mixed | 1622 | 5889 | 3.38(2.26 to 5.07) | |
| **Sex** | | | | |
| Men | 3826 | 19246 | 1.22(0.96 to 1.54) | 0.57 |
| Women | 583 | 18476 | 1.45(0.82 to 2.59) | |
| **Smoking Status** | | | | |
| Never | 552 | 15197 | 2.29(1.15 to 4.55) | 0.08 |
| Former | 1726 | 9741 | 1.27(0.86 to 1.87) | |
| Current | 2114 | 5675 | 0.93(0.62 to 1.38) | |
| **Drinking Status** | | | | |
| Never | 774 | 3398 | 2.71(1.53 to 4.79) | 0.003** |
| Ever | 3366 | 26338 | 1.05(0.79 to 1.38) | |
| **Geographic Region** | | | | |
| Europe | 2003 | 30967 | 1.17(0.89 to 1.52) | 0.0009*** |
| North America | 1354 | 3694 | 1.00(0.68 to 1.48) | |
| South America | 739 | 2239 | 3.05(1.91 to 4.87) | |
| Middle East | 282 | 504 | 3.12(0.82 to 11.88) | |
| **Subsite** | | | | |
| Oral cavity | 5596 | 37723 | 1.04(0.86 to 1.26) | 0.08 |
| Hypopharynx | 898 | 37689 | 0.61(0.36 to 1.03) | |
| Oropharynx (HPV-) | 1473 | 23170 | 0.68(0.46 to 1.02) | |
| Oropharynx (HPV+) | 2212 | 22803 | 0.83(0.62 to 1.10) | |

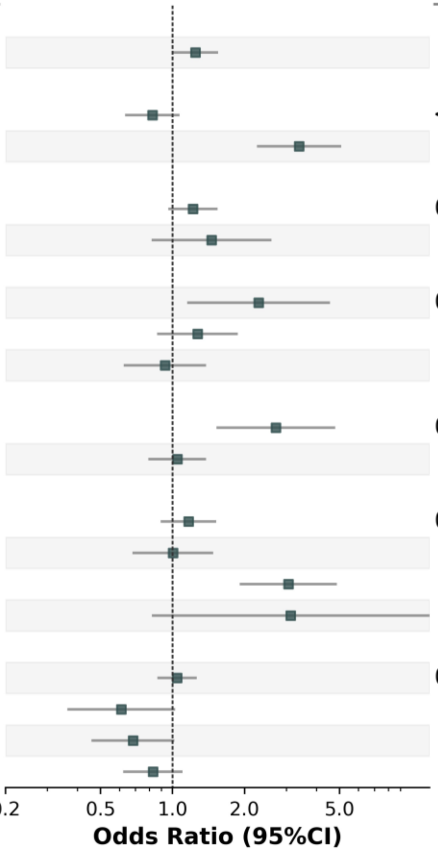


**Supplementary Figure 5. Forest plot of Odds ratios for rs200410709 showing significant heterogeneity in European-vs Mixed ancestry group.** rs200410709, a deletion variant intronic within *STXBP6* (14q12), was linked to a large increased risk of LA (3.38 (2.26, 5.07), p=$3.57x10^{-9}$).

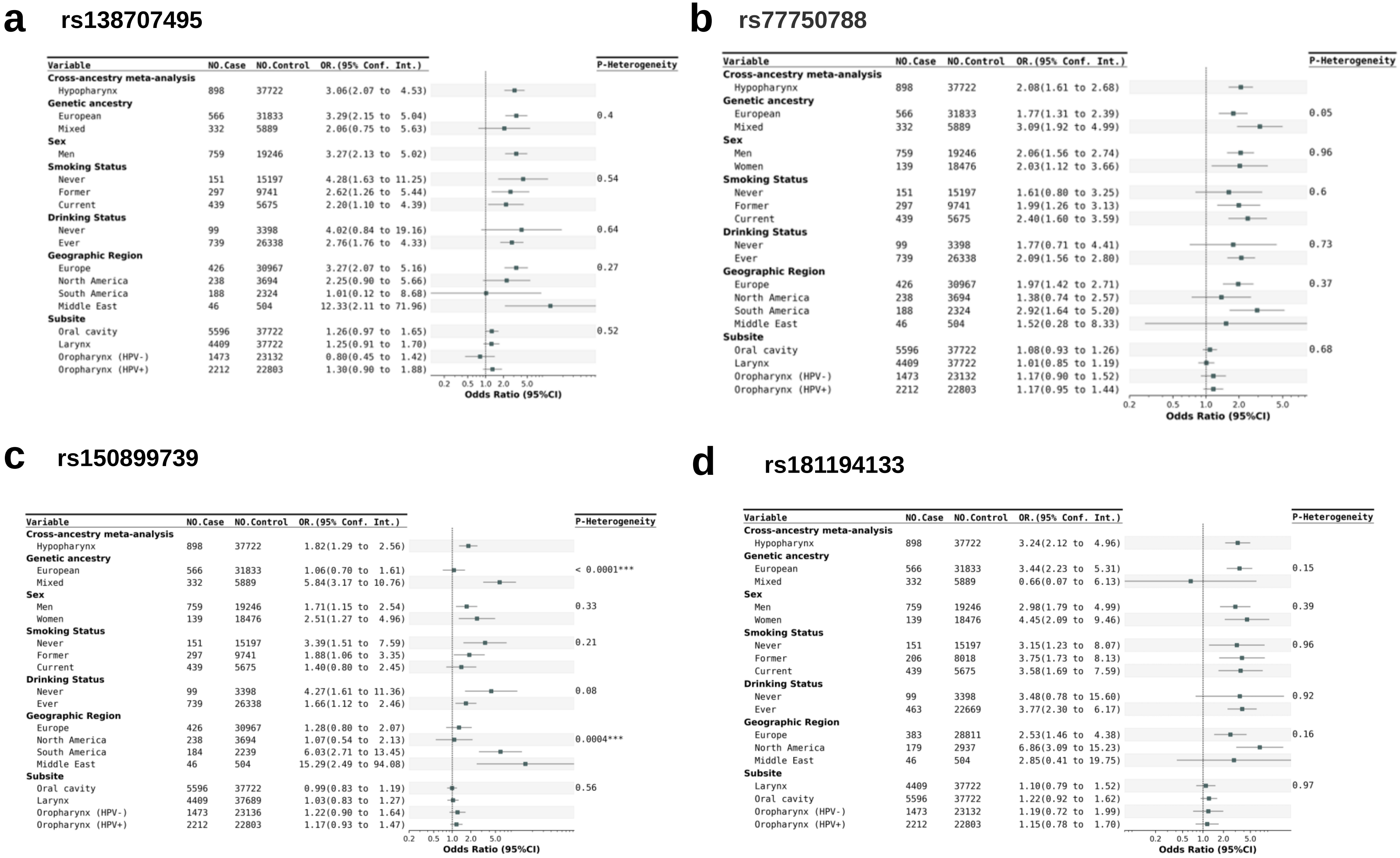


**Supplementary Figure 6. Forest plots of odds ratios for five variants identified in hypopharyngeal cancer (HPC).** a) rs138707495 located in the 3' UTR of *GDF7* at the locus 2p24 identified in meta-analysis. b) rs77750788 near *IGSF9B* at the locus of 11q25. c) rs150899739 in *SASH1* at locus 6q24. d) rs181194133 an intronic variant in *OPCML* at the locus 11q25. e) rs181777026 intergenic variant near *TENM4* at the locus 11q14.

**e** **rs181777026**

| Variable | NO.Case | NO.Control | OR.(95% Conf. Int.) | P-Heterogeneity |
|---|---|---|---|---|
| **Cross-ancestry meta-analysis** | | | | |
| Hypopharynx | 898 | 37722 | 2.51(1.80 to 3.50) | |
| **Genetic ancestry** | | | | |
| European | 566 | 31833 | 2.81(1.94 to 4.05) | 0.17 |
| Mixed | 332 | 5889 | 1.56(0.72 to 3.34) | |
| **Sex** | | | | |
| Men | 759 | 19246 | 2.40(1.65 to 3.48) | 0.37 |
| Women | 100 | 16715 | 3.48(1.67 to 7.22) | |
| **Smoking Status** | | | | |
| Never | 102 | 12587 | 1.87(0.61 to 5.77) | 0.25 |
| Former | 297 | 9741 | 3.26(1.91 to 5.56) | |
| Current | 439 | 5675 | 1.72(0.97 to 3.04) | |
| **Drinking Status** | | | | |
| Never | 99 | 3398 | 1.06(0.19 to 5.72) | 0.31 |
| Ever | 739 | 26338 | 2.58(1.79 to 3.74) | |
| **Geographic Region** | | | | |
| Europe | 426 | 30967 | 2.31(1.50 to 3.55) | 0.43 |
| North America | 238 | 3694 | 3.35(1.79 to 6.25) | |
| South America | 184 | 2239 | 1.66(0.63 to 4.33) | |
| **Subsite** | | | | |
| Oral cavity | 5596 | 37722 | 1.35(1.10 to 1.65) | 0.55 |
| Larynx | 4409 | 37722 | 1.07(0.84 to 1.38) | |
| Oropharynx (HPV-) | 1473 | 23132 | 1.21(0.82 to 1.78) | |
| Oropharynx (HPV+) | 2212 | 22803 | 1.13(0.82 to 1.55) | |

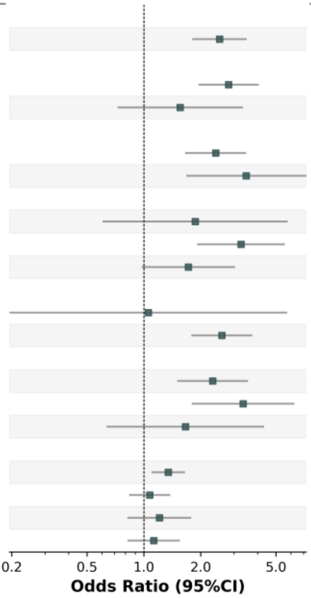

### a Cross-ancestry (all sites combined)

### rs61817953

| Variable | NO.Case | NO.Control | OR.(95% Conf. Int.) | P-Heterogeneity |
|---|---|---|---|---|
| **Cross-ancestry meta-analysis** | | | | |
| All sites combined | 19073 | 38857 | 0.90(0.87 to 0.93) | |
| **Genetic ancestry** | | | | |
| European | 13554 | 32914 | 0.91(0.87 to 0.94) | 0.51 |
| Mixed | 5519 | 5943 | 0.88(0.81 to 0.95) | |
| **Sex** | | | | |
| Men | 14848 | 20074 | 0.91(0.87 to 0.95) | 0.67 |
| Women | 4225 | 18783 | 0.89(0.83 to 0.96) | |
| **Smoking Status** | | | | |
| Never | 3827 | 15197 | 0.90(0.84 to 0.98) | 0.9 |
| Former | 5811 | 9741 | 0.90(0.83 to 0.96) | |
| Current | 6944 | 5675 | 0.92(0.85 to 1.00) | |
| **Drinking Status** | | | | |
| Never | 2983 | 3398 | 0.93(0.83 to 1.03) | 0.71 |
| Ever | 12866 | 26338 | 0.91(0.86 to 0.95) | |
| **Geographic Region** | | | | |
| Europe | 7808 | 30967 | 0.91(0.87 to 0.95) | 0.93 |
| North America | 7555 | 4829 | 0.89(0.84 to 0.96) | |
| South America | 3030 | 2324 | 0.90(0.80 to 1.02) | |
| Middle East | 505 | 504 | 0.91(0.73 to 1.14) | |
| South Asia | 175 | 233 | 1.05(0.74 to 1.48) | |
| **Subsite** | | | | |
| Oral cavity | 5596 | 37722 | 0.92(0.87 to 0.97) | 0.9 |
| Larynx | 4409 | 37722 | 0.88(0.83 to 0.95) | |
| Hypopharynx | 566 | 31833 | 0.88(0.75 to 1.02) | |
| Oropharynx (HPV-) | 1473 | 23132 | 0.91(0.82 to 1.01) | |
| Oropharynx (HPV+) | 2212 | 22803 | 0.89(0.81 to 0.97) | |

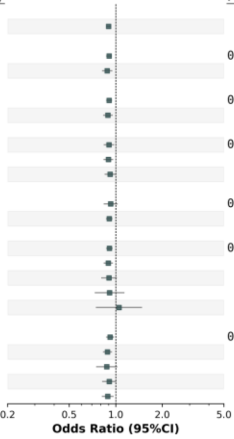


### rs6679311

| Variable | NO.Case | NO.Control | OR.(95% Conf. Int.) | P-Heterogeneity |
|---|---|---|---|---|
| **Cross-ancestry meta-analysis** | | | | |
| All sites combined | 19073 | 38857 | 1.11(1.07 to 1.14) | |
| **Genetic ancestry** | | | | |
| European | 13554 | 32914 | 1.10(1.06 to 1.14) | 0.3 |
| Mixed | 5519 | 5943 | 1.14(1.07 to 1.21) | |
| **Sex** | | | | |
| Men | 14848 | 20074 | 1.10(1.07 to 1.15) | 0.83 |
| Women | 4225 | 18783 | 1.11(1.05 to 1.18) | |
| **Smoking Status** | | | | |
| Never | 3827 | 15197 | 1.11(1.04 to 1.18) | 0.46 |
| Former | 5811 | 9741 | 1.12(1.06 to 1.19) | |
| Current | 6944 | 5675 | 1.06(0.99 to 1.14) | |
| **Drinking Status** | | | | |
| Never | 2983 | 3398 | 1.09(1.00 to 1.19) | 0.97 |
| Ever | 12866 | 26338 | 1.09(1.05 to 1.13) | |
| **Geographic Region** | | | | |
| Europe | 7808 | 30967 | 1.06(1.02 to 1.11) | 0.02* |
| North America | 7555 | 4829 | 1.18(1.11 to 1.25) | |
| South America | 3030 | 2324 | 1.14(1.05 to 1.24) | |
| Middle East | 505 | 504 | 1.07(0.88 to 1.31) | |
| South Asia | 175 | 233 | 0.83(0.60 to 1.16) | |
| **Subsite** | | | | |
| Oral cavity | 5596 | 37722 | 1.09(1.04 to 1.15) | 0.42 |
| Larynx | 4409 | 37722 | 1.11(1.05 to 1.17) | |
| Hypopharynx | 898 | 37722 | 1.22(1.10 to 1.35) | |
| Oropharynx (HPV-) | 1473 | 23132 | 1.13(1.04 to 1.23) | |
| Oropharynx (HPV+) | 2212 | 22803 | 1.13(1.05 to 1.21) | |

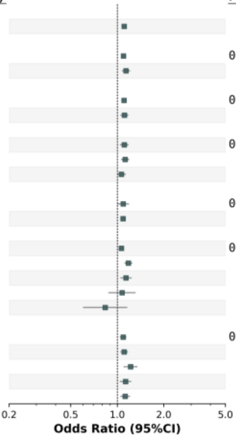


### rs17529509

| Variable | NO.Case | NO.Control | OR.(95% Conf. Int.) | P-Heterogeneity |
|---|---|---|---|---|
| **Cross-ancestry meta-analysis** | | | | |
| All sites combined | 19073 | 38857 | 0.83(0.79 to 0.87) | |
| **Genetic ancestry** | | | | |
| European | 13554 | 32914 | 0.85(0.80 to 0.90) | 0.13 |
| Mixed | 5519 | 5943 | 0.78(0.70 to 0.86) | |
| **Sex** | | | | |
| Men | 14848 | 20074 | 0.81(0.76 to 0.86) | 0.04* |
| Women | 4225 | 18783 | 0.90(0.82 to 0.99) | |
| **Smoking Status** | | | | |
| Never | 3827 | 15197 | 0.95(0.86 to 1.05) | 0.04* |
| Former | 5811 | 9741 | 0.82(0.74 to 0.90) | |
| Current | 6944 | 5675 | 0.81(0.73 to 0.90) | |
| **Drinking Status** | | | | |
| Never | 2983 | 3398 | 0.89(0.78 to 1.02) | 0.36 |
| Ever | 12866 | 26338 | 0.83(0.78 to 0.89) | |
| **Geographic Region** | | | | |
| Europe | 7808 | 30967 | 0.84(0.79 to 0.90) | 0.21 |
| North America | 7555 | 4829 | 0.87(0.80 to 0.95) | |
| South America | 3030 | 2324 | 0.72(0.62 to 0.84) | |
| Middle East | 505 | 504 | 0.81(0.61 to 1.07) | |
| South Asia | 175 | 233 | 1.33(0.59 to 3.01) | |
| **Subsite** | | | | |
| Oral cavity | 5596 | 37722 | 0.84(0.78 to 0.91) | 0.01* |
| Larynx | 4409 | 37722 | 0.86(0.79 to 0.94) | |
| Hypopharynx | 2898 | 37722 | 0.63(0.51 to 0.77) | |
| Oropharynx (HPV-) | 1473 | 23132 | 0.80(0.69 to 0.93) | |
| Oropharynx (HPV+) | 2212 | 22803 | 0.93(0.83 to 1.04) | |

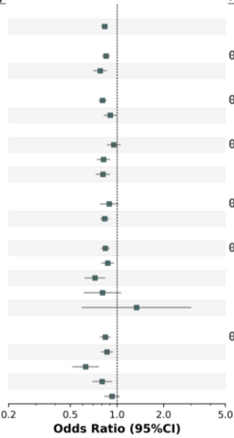


### rs58223772

| Variable | NO.Case | NO.Control | OR.(95% Conf. Int.) | P-Heterogeneity |
|---|---|---|---|---|
| **Cross-ancestry meta-analysis** | | | | |
| All sites combined | 19073 | 38857 | 0.90(0.87 to 0.93) | |
| **Genetic ancestry** | | | | |
| European | 13554 | 32914 | 0.92(0.88 to 0.95) | 0.17 |
| Mixed | 5519 | 5943 | 0.87(0.82 to 0.93) | |
| **Sex** | | | | |
| Men | 14848 | 20074 | 0.89(0.86 to 0.93) | 0.25 |
| Women | 4225 | 18783 | 0.93(0.87 to 0.99) | |
| **Smoking Status** | | | | |
| Never | 3827 | 15197 | 0.93(0.87 to 1.00) | 0.03* |
| Former | 5811 | 9741 | 0.95(0.89 to 1.01) | |
| Current | 6944 | 5675 | 0.84(0.79 to 0.91) | |
| **Drinking Status** | | | | |
| Never | 2983 | 3398 | 0.94(0.86 to 1.03) | 0.59 |
| Ever | 12866 | 26338 | 0.91(0.87 to 0.95) | |
| **Geographic Region** | | | | |
| Europe | 7808 | 30967 | 0.93(0.89 to 0.98) | 0.08 |
| North America | 7555 | 4829 | 0.90(0.85 to 0.96) | |
| South America | 3030 | 2324 | 0.81(0.74 to 0.89) | |
| Middle East | 505 | 504 | 0.94(0.77 to 1.14) | |
| South Asia | 175 | 233 | 0.98(0.73 to 1.32) | |
| **Subsite** | | | | |
| Oral cavity | 3511 | 31833 | 0.86(0.80 to 0.92) | 0.0008*** |
| Larynx | 4409 | 37722 | 0.94(0.88 to 1.00) | |
| Hypopharynx | 898 | 37722 | 0.87(0.78 to 0.97) | |
| Oropharynx (HPV-) | 1473 | 23132 | 0.77(0.70 to 0.85) | |
| Oropharynx (HPV+) | 2212 | 22803 | 0.98(0.91 to 1.06) | |

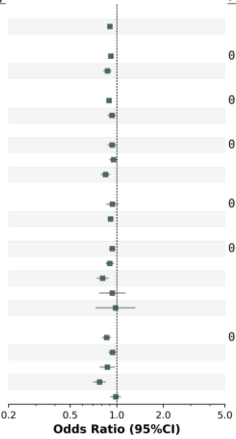


**Supplementary Figure 7.** For each independent top hit identified in the cross-ancestry GWAS, analyses were stratified by sex, smoking status, drinking status, geographic region, and cancer subsites. Forest plots of variants identified by cancer subsite: a) top hit variants identified in all sites combined, b) variants identified in the oral cavity, c) variants identified in the larynx, and d) variants identified in the hypopharynx.

**a** **Cross-ancestry (all sites combined)**

## rs1131769

| Variable | NO.Case | NO.Control | OR.(95% Conf. Int.) | P-Heterogeneity |
|---|---|---|---|---|
| **Cross-ancestry meta-analysis** | | | | |
| All sites combined | 19073 | 38857 | 1.13(1.09 to 1.18) | |
| **Genetic ancestry** | | | | |
| European | 13554 | 32914 | 1.15(1.09 to 1.20) | 0.45 |
| Mixed | 5519 | 5943 | 1.10(1.02 to 1.20) | |
| **Sex** | | | | |
| Men | 14848 | 20074 | 1.13(1.08 to 1.19) | 0.68 |
| Women | 4225 | 18783 | 1.11(1.03 to 1.20) | |
| **Smoking Status** | | | | |
| Never | 3827 | 15197 | 1.10(1.01 to 1.20) | 0.67 |
| Former | 5811 | 9741 | 1.16(1.07 to 1.26) | |
| Current | 6944 | 5675 | 1.14(1.05 to 1.25) | |
| **Drinking Status** | | | | |
| Never | 2983 | 3398 | 1.18(1.05 to 1.33) | 0.83 |
| Ever | 12866 | 26338 | 1.16(1.10 to 1.22) | |
| **Geographic Region** | | | | |
| Europe | 7808 | 30967 | 1.16(1.10 to 1.22) | 0.83 |
| North America | 7555 | 4829 | 1.12(1.04 to 1.21) | |
| South America | 3030 | 2324 | 1.08(0.97 to 1.21) | |
| Middle East | 505 | 504 | 1.21(0.93 to 1.57) | |
| South Asia | 175 | 233 | 1.15(0.70 to 1.89) | |
| **Subsite** | | | | |
| Oral cavity | 5596 | 37690 | 1.22(1.15 to 1.30) | 0.001** |
| Larynx | 4409 | 37722 | 1.15(1.07 to 1.23) | |
| Hypopharynx | 898 | 37689 | 1.15(1.00 to 1.32) | |
| Oropharynx (HPV-) | 1473 | 23132 | 1.13(1.01 to 1.26) | |
| Oropharynx (HPV+) | 2212 | 22803 | 0.94(0.85 to 1.04) | |

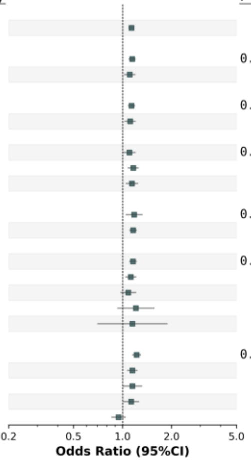


## rs541752611

| Variable | NO.Case | NO.Control | OR.(95% Conf. Int.) | P-Heterogeneity |
|---|---|---|---|---|
| **Cross-ancestry meta-analysis** | | | | |
| All sites combined | 19073 | 38857 | 1.22(1.14 to 1.31) | |
| **Genetic ancestry** | | | | |
| European | 13554 | 32914 | 1.26(1.16 to 1.36) | 0.15 |
| Mixed | 5519 | 5943 | 1.12(0.98 to 1.28) | |
| **Sex** | | | | |
| Men | 14848 | 20074 | 1.25(1.15 to 1.36) | 0.32 |
| Women | 4225 | 18783 | 1.16(1.01 to 1.32) | |
| **Smoking Status** | | | | |
| Never | 3827 | 15197 | 1.21(1.06 to 1.39) | 0.23 |
| Former | 5811 | 9741 | 1.26(1.11 to 1.43) | |
| Current | 6944 | 5675 | 1.06(0.91 to 1.24) | |
| **Drinking Status** | | | | |
| Never | 2983 | 3398 | 1.06(0.86 to 1.32) | 0.5 |
| Ever | 12866 | 26338 | 1.15(1.05 to 1.25) | |
| **Geographic Region** | | | | |
| Europe | 7808 | 30967 | 1.23(1.13 to 1.34) | 0.59 |
| North America | 7555 | 4829 | 1.21(1.07 to 1.38) | |
| South America | 3030 | 2324 | 1.12(0.95 to 1.33) | |
| Middle East | 505 | 504 | 0.70(0.32 to 1.52) | |
| South Asia | 175 | 233 | 1.09(0.16 to 7.36) | |
| **Subsite** | | | | |
| Oral cavity | 5596 | 37690 | 1.13(1.01 to 1.25) | 0.0001*** |
| Larynx | 4409 | 37722 | 1.07(0.94 to 1.21) | |
| Hypopharynx | 898 | 37689 | 1.24(0.99 to 1.55) | |
| Oropharynx (HPV-) | 1473 | 23132 | 1.19(0.99 to 1.43) | |
| Oropharynx (HPV+) | 2212 | 22803 | 1.64(1.42 to 1.89) | |

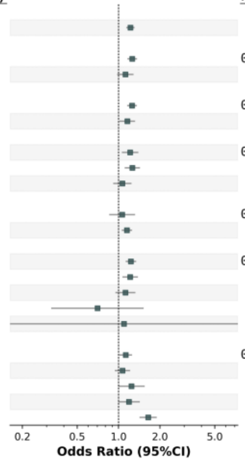


## rs9266806

| Variable | NO.Case | NO.Control | OR.(95% Conf. Int.) | P-Heterogeneity |
|---|---|---|---|---|
| **Cross-ancestry meta-analysis** | | | | |
| All sites combined | 19073 | 38857 | 0.89(0.86 to 0.92) | |
| **Genetic ancestry** | | | | |
| European | 13554 | 32914 | 0.87(0.84 to 0.91) | 0.1 |
| Mixed | 5519 | 5943 | 0.93(0.87 to 0.99) | |
| **Sex** | | | | |
| Men | 14848 | 20074 | 0.88(0.85 to 0.92) | 0.49 |
| Women | 4225 | 18783 | 0.91(0.85 to 0.97) | |
| **Smoking Status** | | | | |
| Never | 3827 | 15197 | 0.87(0.81 to 0.94) | 0.37 |
| Former | 5811 | 9741 | 0.90(0.84 to 0.96) | |
| Current | 6944 | 5675 | 0.94(0.87 to 1.01) | |
| **Drinking Status** | | | | |
| Never | 2983 | 3398 | 0.99(0.90 to 1.09) | 0.12 |
| Ever | 12866 | 26338 | 0.91(0.87 to 0.95) | |
| **Geographic Region** | | | | |
| Europe | 7808 | 30967 | 0.89(0.85 to 0.93) | 0.43 |
| North America | 7555 | 4829 | 0.88(0.82 to 0.94) | |
| South America | 3030 | 2324 | 0.95(0.87 to 1.05) | |
| Middle East | 505 | 504 | 0.97(0.80 to 1.19) | |
| South Asia | 175 | 233 | 1.02(0.75 to 1.40) | |
| **Subsite** | | | | |
| Oral cavity | 5595 | 37690 | 0.93(0.88 to 0.98) | < 0.0001*** |
| Larynx | 4409 | 37722 | 0.97(0.92 to 1.04) | |
| Hypopharynx | 898 | 37689 | 0.88(0.78 to 0.99) | |
| Oropharynx (HPV-) | 1473 | 23132 | 0.93(0.85 to 1.02) | |
| Orophraynx (HPV+) | 2212 | 22803 | 0.71(0.65 to 0.77) | |

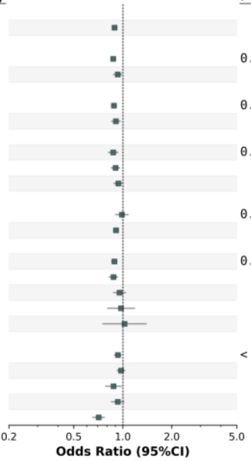


## rs9282195

| Variable | NO.Case | NO.Control | OR.(95% Conf. Int.) | P-Heterogeneity |
|---|---|---|---|---|
| **Cross-ancestry meta-analysis** | | | | |
| All sites combined | 19073 | 38857 | 0.84(0.81 to 0.87) | |
| **Genetic ancestry** | | | | |
| European | 13554 | 32914 | 0.83(0.80 to 0.86) | 0.42 |
| Mixed | 5519 | 5943 | 0.86(0.80 to 0.93) | |
| **Sex** | | | | |
| Men | 14848 | 20074 | 0.83(0.79 to 0.86) | 0.15 |
| Women | 4225 | 18783 | 0.87(0.82 to 0.93) | |
| **Smoking Status** | | | | |
| Never | 3827 | 15197 | 0.85(0.79 to 0.91) | 0.46 |
| Former | 5811 | 9741 | 0.84(0.79 to 0.90) | |
| Current | 6944 | 5675 | 0.89(0.83 to 0.96) | |
| **Drinking Status** | | | | |
| Never | 2983 | 3398 | 0.87(0.79 to 0.97) | 0.58 |
| Ever | 12866 | 26338 | 0.84(0.81 to 0.88) | |
| **Geographic Region** | | | | |
| Europe | 7808 | 30967 | 0.85(0.81 to 0.89) | 0.53 |
| North America | 7555 | 4829 | 0.82(0.77 to 0.88) | |
| South America | 3030 | 2324 | 0.84(0.76 to 0.93) | |
| Middle East | 505 | 504 | 0.76(0.56 to 1.02) | |
| South Asia | 175 | 233 | 1.13(0.76 to 1.69) | |
| **Subsite** | | | | |
| Oral cavity | 5596 | 37722 | 0.84(0.79 to 0.88) | < .0001*** |
| Larynx | 4409 | 37722 | 0.94(0.88 to 1.00) | |
| Hypopharynx | 898 | 37722 | 0.83(0.73 to 0.94) | |
| Oropharynx (HPV-) | 1473 | 23132 | 0.85(0.77 to 0.94) | |
| Oropharynx (HPV+) | 2212 | 22803 | 0.69(0.63 to 0.75) | |

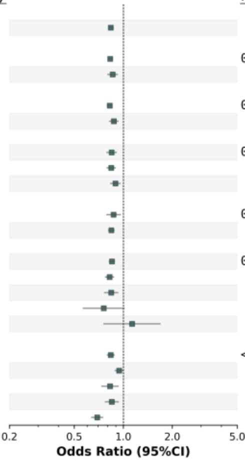

### a Cross-ancestry (all sites combined)

### rs13215307

| Variable | NO.Case | NO.Control | OR.(95% Conf. Int.) | P-Heterogeneity |
|---|---|---|---|---|
| **Cross-ancestry meta-analysis** | | | | |
| All sites combined | 19073 | 38857 | 0.83(0.77 to 0.88) | |
| **Genetic ancestry** | | | | |
| European | 13554 | 32914 | 0.81(0.75 to 0.87) | 0.23 |
| Mixed | 5519 | 5943 | 0.88(0.78 to 1.00) | |
| **Sex** | | | | |
| Men | 14848 | 20074 | 0.82(0.76 to 0.88) | 0.48 |
| Women | 4225 | 18783 | 0.87(0.76 to 0.99) | |
| **Smoking Status** | | | | |
| Never | 3827 | 15197 | 0.84(0.73 to 0.97) | 0.99 |
| Former | 5811 | 9741 | 0.83(0.74 to 0.95) | |
| Current | 6944 | 5675 | 0.84(0.73 to 0.95) | |
| **Drinking Status** | | | | |
| Never | 2983 | 3398 | 0.93(0.77 to 1.12) | 0.24 |
| Ever | 12866 | 26338 | 0.82(0.75 to 0.89) | |
| **Geographic Region** | | | | |
| Europe | 7808 | 30967 | 0.87(0.80 to 0.94) | 0.98 |
| North America | 7555 | 4829 | 0.83(0.73 to 0.94) | |
| South America | 3030 | 2324 | 0.88(0.73 to 1.05) | |
| Middle East | 505 | 504 | 0.88(0.61 to 1.26) | |
| South Asia | 175 | 233 | 0.92(0.15 to 5.67) | |
| **Subsite** | | | | |
| Oral cavity | 5596 | 37723 | 0.82(0.74 to 0.91) | 0.4 |
| Larynx | 4409 | 37722 | 0.91(0.81 to 1.02) | |
| Hypopharynx | 898 | 37722 | 0.81(0.64 to 1.02) | |
| Oropharynx (HPV-) | 1473 | 23132 | 0.73(0.60 to 0.89) | |
| Oropharynx (HPV+) | 2212 | 22803 | 0.81(0.69 to 0.95) | |

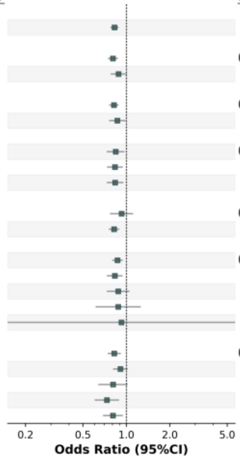


### rs12314527

| Variable | NO.Case | NO.Control | OR.(95% Conf. Int.) | P-Heterogeneity |
|---|---|---|---|---|
| **Cross-ancestry meta-analysis** | | | | |
| All sites combined | 19073 | 38857 | 1.11(1.07 to 1.15) | |
| **Genetic ancestry** | | | | |
| European | 13554 | 32914 | 1.12(1.07 to 1.17) | 0.5 |
| Mixed | 5519 | 5943 | 1.09(1.02 to 1.16) | |
| **Sex** | | | | |
| Men | 14848 | 20074 | 1.12(1.07 to 1.17) | 0.52 |
| Women | 4225 | 18783 | 1.09(1.02 to 1.17) | |
| **Smoking Status** | | | | |
| Never | 3827 | 15197 | 1.02(0.94 to 1.10) | 0.03* |
| Former | 5811 | 9741 | 1.12(1.05 to 1.20) | |
| Current | 6944 | 5675 | 1.17(1.09 to 1.26) | |
| **Drinking Status** | | | | |
| Never | 2983 | 3398 | 1.05(0.95 to 1.16) | 0.16 |
| Ever | 12866 | 26338 | 1.13(1.08 to 1.18) | |
| **Geographic Region** | | | | |
| Europe | 7808 | 30967 | 1.11(1.06 to 1.17) | 0.74 |
| North America | 7555 | 4829 | 1.12(1.05 to 1.20) | |
| South America | 3030 | 2324 | 1.04(0.95 to 1.14) | |
| Middle East | 505 | 504 | 1.13(0.91 to 1.41) | |
| South Asia | 175 | 233 | 1.11(0.79 to 1.58) | |
| **Subsite** | | | | |
| Oral cavity | 5596 | 37723 | 1.05(1.00 to 1.11) | 0.0001*** |
| Larynx | 4409 | 37722 | 1.20(1.13 to 1.28) | |
| Hypopharynx | 898 | 37689 | 1.17(1.04 to 1.32) | |
| Oropharynx (HPV-) | 1473 | 23132 | 1.24(1.13 to 1.36) | |
| Oropharynx (HPV+) | 2212 | 22803 | 0.98(0.90 to 1.07) | |

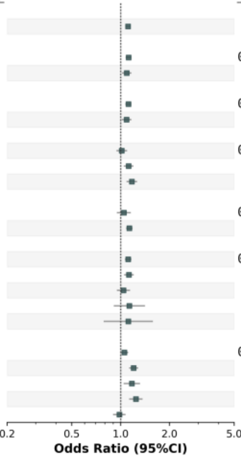


### rs7334543

| Variable | NO.Case | NO.Control | OR.(95% Conf. Int.) | P-Heterogeneity |
|---|---|---|---|---|
| **Cross-ancestry meta-analysis** | | | | |
| All sites combined | 19073 | 38857 | 0.91(0.88 to 0.94) | |
| **Genetic ancestry** | | | | |
| European | 13554 | 32914 | 0.91(0.88 to 0.95) | 0.98 |
| Mixed | 5519 | 5943 | 0.91(0.85 to 0.97) | |
| **Sex** | | | | |
| Men | 14848 | 20074 | 0.91(0.87 to 0.94) | 0.46 |
| Women | 4225 | 18783 | 0.93(0.88 to 0.99) | |
| **Smoking Status** | | | | |
| Never | 3827 | 15197 | 0.90(0.84 to 0.97) | 0.78 |
| Former | 5811 | 9741 | 0.93(0.88 to 0.99) | |
| Current | 6944 | 5675 | 0.92(0.86 to 0.99) | |
| **Drinking Status** | | | | |
| Never | 2983 | 3398 | 0.94(0.85 to 1.03) | 0.47 |
| Ever | 12866 | 26338 | 0.90(0.86 to 0.94) | |
| **Geographic Region** | | | | |
| Europe | 7808 | 30967 | 0.91(0.87 to 0.95) | 0.6 |
| North America | 7555 | 4829 | 0.92(0.87 to 0.98) | |
| South America | 3030 | 2324 | 0.93(0.85 to 1.02) | |
| Middle East | 505 | 504 | 1.00(0.78 to 1.27) | |
| South Asia | 175 | 233 | 0.69(0.47 to 1.03) | |
| **Subsite** | | | | |
| Oral cavity | 5596 | 37722 | 0.96(0.92 to 1.01) | 0.22 |
| Larynx | 4409 | 37722 | 0.90(0.85 to 0.95) | |
| Hypopharynx | 898 | 37722 | 0.88(0.78 to 0.98) | |
| Oropharynx (HPV-) | 1473 | 23132 | 0.88(0.80 to 0.96) | |
| Oropharynx (HPV+) | 2212 | 22803 | 0.91(0.85 to 0.98) | |

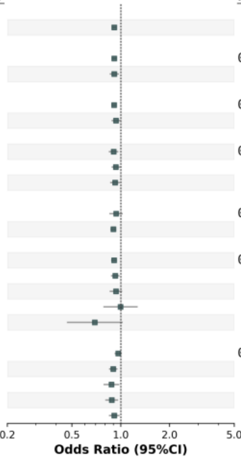


### rs78378222

| Variable | NO.Case | NO.Control | OR.(95% Conf. Int.) | P-Heterogeneity |
|---|---|---|---|---|
| **Cross-ancestry meta-analysis** | | | | |
| All sites combined | 19073 | 38857 | 0.64(0.54 to 0.75) | |
| **Genetic Ancestry** | | | | |
| European | 13554 | 32914 | 0.62(0.52 to 0.73) | 0.3 |
| Mixed* | 5519 | 5943 | 0.82(0.49 to 1.37) | |
| **Sex** | | | | |
| Men | 14848 | 20074 | 0.64(0.53 to 0.77) | 0.87 |
| Women | 4225 | 18783 | 0.66(0.48 to 0.89) | |
| **Smoking Status** | | | | |
| Never | 3827 | 15197 | 0.61(0.43 to 0.85) | 0.15 |
| Former | 5811 | 9741 | 0.82(0.61 to 1.09) | |
| Current | 6944 | 5675 | 0.53(0.36 to 0.77) | |
| **Drinking Status** | | | | |
| Never | 2983 | 3398 | 0.73(0.43 to 1.23) | 0.64 |
| Ever | 12866 | 26338 | 0.64(0.52 to 0.79) | |
| **Geographic Region** | | | | |
| Europe | 7808 | 30967 | 0.56(0.45 to 0.68) | 0.25 |
| North America | 7555 | 4829 | 0.74(0.57 to 0.97) | |
| South America | 3030 | 2324 | 0.68(0.34 to 1.38) | |
| **Subsite** | | | | |
| Oral cavity | 5596 | 37722 | 0.51(0.38 to 0.68) | 0.02* |
| Larynx | 4409 | 37722 | 0.57(0.41 to 0.79) | |
| Hypopharynx | 898 | 37722 | 0.69(0.36 to 1.29) | |
| Oropharynx (HPV-) | 1473 | 23132 | 0.58(0.35 to 0.96) | |
| Oropharynx (HPV+) | 2212 | 22803 | 1.00(0.75 to 1.35) | |

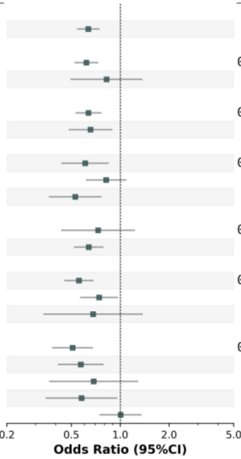

**b** **Cross-ancestry (oral cavity)**

## rs3846449

| Variable | NO.Case | NO.Control | OR.(95% Conf. Int.) | P-Heterogeneity |
|---|---|---|---|---|
| **Cross-ancestry meta-analysis** | | | | |
| Oral cavity | 5596 | 37722 | 0.86(0.82 to 0.91) | |
| **Genetic ancestry** | | | | |
| European | 3511 | 31833 | 0.86(0.81 to 0.92) | 0.98 |
| Mixed | 2085 | 5889 | 0.86(0.79 to 0.94) | |
| **Sex** | | | | |
| Men | 3749 | 19246 | 0.85(0.80 to 0.90) | 0.38 |
| women | 1847 | 18476 | 0.89(0.81 to 0.97) | |
| **Smoking Status** | | | | |
| Never | 1498 | 15197 | 0.94(0.86 to 1.04) | 0.05 |
| Former | 1598 | 9741 | 0.85(0.77 to 0.93) | |
| Current | 2339 | 5675 | 0.80(0.73 to 0.88) | |
| **Drinking Status** | | | | |
| Never | 1218 | 3398 | 0.91(0.81 to 1.02) | 0.44 |
| Ever | 4098 | 26338 | 0.87(0.81 to 0.92) | |
| **Geographic Region** | | | | |
| Europe | 2337 | 30967 | 0.89(0.83 to 0.96) | 0.1 |
| North America | 1770 | 3694 | 0.82(0.74 to 0.90) | |
| South America | 1194 | 2324 | 0.79(0.71 to 0.89) | |
| Middle East | 153 | 504 | 1.09(0.82 to 1.45) | |
| South Asia | 142 | 233 | 1.01(0.74 to 1.38) | |
| **Subsite** | | | | |
| Larynx | 4409 | 37722 | 0.94(0.88 to 1.00) | 0.0007*** |
| Hypopharynx | 898 | 37722 | 0.87(0.78 to 0.98) | |
| Oropharynx (HPV-) | 1473 | 23132 | 0.77(0.70 to 0.85) | |
| Oropharynx (HPV+) | 2212 | 22803 | 0.98(0.91 to 1.06) | |

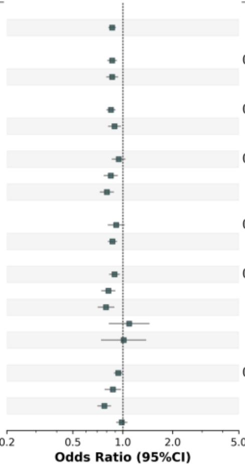


## rs1229984

| Variable | NO.Case | NO.Control | OR.(95% Conf. Int.) | P-Heterogeneity |
|---|---|---|---|---|
| **Cross-ancestry meta-analysis** | | | | |
| Oral cavity | 5596 | 37722 | 0.74(0.66 to 0.82) | |
| **Genetic ancestry** | | | | |
| European | 3511 | 31833 | 0.65(0.54 to 0.78) | 0.1 |
| Mixed | 2085 | 5889 | 0.79(0.69 to 0.90) | |
| **Sex** | | | | |
| Men | 3749 | 19246 | 0.65(0.56 to 0.74) | 0.001** |
| Women | 1847 | 18476 | 0.94(0.78 to 1.12) | |
| **Smoking Status** | | | | |
| Never | 1498 | 15197 | 1.08(0.91 to 1.29) | < 0.0001*** |
| Former | 1598 | 9741 | 0.90(0.74 to 1.10) | |
| Current | 2339 | 5675 | 0.43(0.34 to 0.55) | |
| **Drinking Status** | | | | |
| Never | 1218 | 3398 | 1.22(1.01 to 1.46) | < 0.0001*** |
| Ever | 4098 | 26338 | 0.58(0.50 to 0.68) | |
| **Geographic Region** | | | | |
| Europe | 2337 | 30967 | 0.61(0.50 to 0.73) | 0.04* |
| North America | 1770 | 3694 | 0.70(0.56 to 0.88) | |
| South America | 1194 | 2324 | 0.71(0.56 to 0.90) | |
| Middle East | 153 | 504 | 0.98(0.71 to 1.35) | |
| South Asia | 142 | 233 | 2.39(0.65 to 8.77) | |
| **Subsite** | | | | |
| Larynx | 4409 | 37722 | 0.80(0.71 to 0.90) | 0.001** |
| Hypopharynx | 898 | 37722 | 0.57(0.43 to 0.77) | |
| Oropharynx (HPV-) | 948 | 18166 | 0.35(0.22 to 0.54) | |
| Oropharynx (HPV+) | 1980 | 18166 | 0.82(0.65 to 1.04) | |

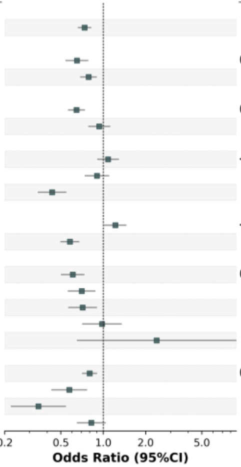


## rs7726159

| Variable | NO.Case | NO.Control | OR.(95% Conf. Int.) | P-Heterogeneity |
|---|---|---|---|---|
| **Cross-ancestry meta-analysis** | | | | |
| Oral cavity | 5596 | 37722 | 1.18(1.12 to 1.23) | |
| **Genetic ancestry** | | | | |
| European | 3511 | 31833 | 1.18(1.12 to 1.25) | 0.86 |
| Mixed | 142 | 233 | 1.10(0.81 to 1.48) | |
| **Sex** | | | | |
| Men | 3749 | 19246 | 1.15(1.09 to 1.22) | 0.17 |
| Women | 1847 | 18476 | 1.23(1.14 to 1.33) | |
| **Smoking Status** | | | | |
| Never | 1498 | 15197 | 1.25(1.15 to 1.36) | 0.28 |
| Former | 1598 | 9741 | 1.19(1.10 to 1.30) | |
| Current | 2339 | 5675 | 1.13(1.04 to 1.23) | |
| **Drinking Status** | | | | |
| Never | 1218 | 3398 | 1.26(1.14 to 1.40) | 0.3 |
| Ever | 4098 | 26338 | 1.19(1.12 to 1.25) | |
| **Geographic Region** | | | | |
| Europe | 2337 | 30967 | 1.19(1.12 to 1.27) | 0.99 |
| North America | 1770 | 3694 | 1.19(1.09 to 1.30) | |
| South America | 1194 | 2324 | 1.19(1.08 to 1.33) | |
| Middle East | 153 | 504 | 1.15(0.88 to 1.51) | |
| South Asia | 142 | 233 | 1.10(0.81 to 1.49) | |
| **Subsite** | | | | |
| Larynx | 4409 | 37722 | 0.98(0.93 to 1.04) | 0.46 |
| Hypopharynx | 898 | 37722 | 1.03(0.93 to 1.14) | |
| Oropharynx (HPV-) | 1473 | 23132 | 1.06(0.98 to 1.15) | |
| Oropharynx (HPV+) | 2212 | 22803 | 1.01(0.94 to 1.09) | |

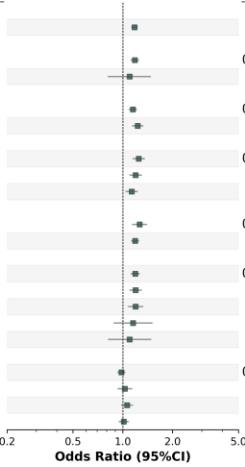


## rs60622800

| Variable | NO.Case | NO.Control | OR.(95% Conf. Int.) | P-Heterogeneity |
|---|---|---|---|---|
| **Cross-ancestry meta-analysis** | | | | |
| Oral cavity | 5596 | 37722 | 1.19(1.13 to 1.24) | |
| **Genetic ancestry** | | | | |
| European | 3511 | 31833 | 1.20(1.13 to 1.26) | 0.65 |
| Mixed | 1470 | 4128 | 1.17(1.08 to 1.26) | |
| **Sex** | | | | |
| Men | 3749 | 19246 | 1.15(1.09 to 1.21) | 0.06 |
| Women | 1847 | 18476 | 1.25(1.16 to 1.36) | |
| **Smoking Status** | | | | |
| Never | 1498 | 15197 | 1.23(1.13 to 1.34) | 0.009** |
| Former | 1598 | 9741 | 1.28(1.17 to 1.38) | |
| Current | 2339 | 5675 | 1.07(0.98 to 1.16) | |
| **Drinking Status** | | | | |
| Never | 1218 | 3398 | 1.28(1.16 to 1.43) | 0.12 |
| Ever | 4098 | 26338 | 1.17(1.11 to 1.24) | |
| **Geographic Region** | | | | |
| Europe | 2337 | 30967 | 1.14(1.07 to 1.22) | 0.05 |
| North America | 1770 | 3694 | 1.28(1.18 to 1.40) | |
| South America | 1194 | 2324 | 1.11(1.00 to 1.23) | |
| Middle East | 153 | 504 | 1.37(1.03 to 1.82) | |
| South Asia | 142 | 233 | 0.87(0.58 to 1.31) | |
| **Subsite** | | | | |
| Larynx | 4409 | 37722 | 0.97(0.92 to 1.02) | 0.5 |
| Hypopharynx | 898 | 37722 | 0.97(0.87 to 1.07) | |
| Oropharynx (HPV-) | 1473 | 23132 | 0.93(0.86 to 1.01) | |
| Oropharynx (HPV+) | 2212 | 22803 | 0.91(0.85 to 0.97) | |

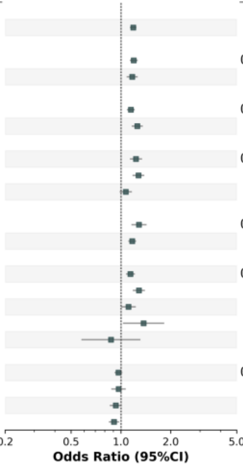

**b** **Cross-ancestry (oral cavity)**

### rs31493

| Variable | NO.Case | NO.Control | OR.(95% Conf. Int.) | P-Heterogeneity |
|---|---|---|---|---|
| **Cross-ancestry meta-analysis** | | | | |
| Oral cavity | 5596 | 37722 | 0.85(0.81 to 0.90) | |
| **Genetic ancestry** | | | | |
| European | 3511 | 31833 | 0.85(0.80 to 0.90) | 0.85 |
| Mixed | 2085 | 5889 | 0.86(0.79 to 0.93) | |
| **Sex** | | | | |
| Men | 3749 | 19246 | 0.85(0.80 to 0.90) | 0.61 |
| Women | 1847 | 18476 | 0.87(0.80 to 0.94) | |
| **Smoking Status** | | | | |
| Never | 1498 | 15197 | 0.88(0.80 to 0.96) | 0.66 |
| Former | 1598 | 9741 | 0.83(0.76 to 0.91) | |
| Current | 2339 | 5675 | 0.85(0.78 to 0.93) | |
| **Drinking Status** | | | | |
| Never | 1218 | 3398 | 0.81(0.73 to 0.90) | 0.29 |
| Ever | 4098 | 26338 | 0.87(0.82 to 0.92) | |
| **Geographic Region** | | | | |
| Europe | 2337 | 30967 | 0.86(0.80 to 0.92) | 0.93 |
| North America | 1770 | 3694 | 0.89(0.81 to 0.97) | |
| South America | 1194 | 2324 | 0.84(0.75 to 0.94) | |
| Middle East | 153 | 504 | 0.93(0.70 to 1.24) | |
| South Asia | 142 | 233 | 0.90(0.66 to 1.21) | |
| **Subsite** | | | | |
| Larynx | 4409 | 37722 | 0.99(0.94 to 1.05) | 0.17 |
| Hypopharynx | 898 | 37722 | 1.10(0.99 to 1.22) | |
| Oropharynx (HPV-) | 1473 | 23132 | 0.95(0.87 to 1.03) | |
| Oropharynx (HPV+) | 2212 | 22803 | 1.01(0.94 to 1.09) | |

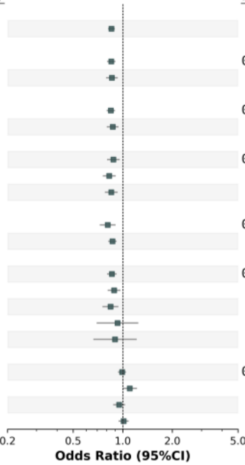


### rs28419191

| Variable | NO.Case | NO.Control | OR.(95% Conf. Int.) | P-Heterogeneity |
|---|---|---|---|---|
| **Cross-ancestry meta-analysis** | | | | |
| Oral cavity | 5596 | 37722 | 1.23(1.15 to 1.31) | |
| **Genetic ancestry** | | | | |
| European | 3511 | 31833 | 1.22(1.12 to 1.32) | 0.77 |
| Mixed | 2085 | 5889 | 1.24(1.12 to 1.38) | |
| **Sex** | | | | |
| Men | 3749 | 19246 | 1.26(1.17 to 1.36) | 0.18 |
| Women | 1847 | 18476 | 1.15(1.03 to 1.29) | |
| **Smoking Status** | | | | |
| Never | 1498 | 15197 | 1.16(1.03 to 1.31) | |
| Former | 1598 | 9741 | 1.24(1.10 to 1.39) | 0.74 |
| Current | 2339 | 5675 | 1.23(1.09 to 1.39) | |
| **Drinking Status** | | | | |
| Never | 1218 | 3398 | 1.23(1.06 to 1.43) | 0.99 |
| Ever | 4098 | 26338 | 1.23(1.14 to 1.33) | |
| **Geographic Region** | | | | |
| Europe | 2337 | 30967 | 1.18(1.08 to 1.29) | 0.79 |
| North America | 1770 | 3694 | 1.26(1.12 to 1.42) | |
| South America | 1194 | 2324 | 1.28(1.11 to 1.48) | |
| Middle East | 153 | 504 | 1.30(0.89 to 1.92) | |
| South Asia | 142 | 233 | 1.04(0.60 to 1.80) | |
| **Subsite** | | | | |
| Larynx | 4409 | 37722 | 1.15(1.07 to 1.24) | 0.01* |
| Hypopharynx | 898 | 37722 | 1.12(0.97 to 1.29) | |
| Oropharynx (HPV-) | 1473 | 23132 | 1.14(1.02 to 1.28) | |
| Oropharynx (HPV+) | 2212 | 22803 | 0.94(0.85 to 1.04) | |

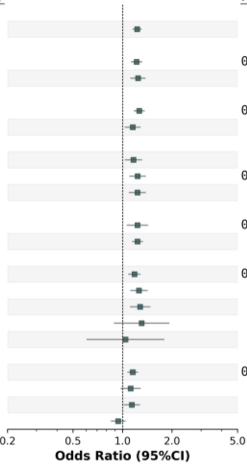


### rs9271300

| Variable | NO.Case | NO.Control | OR.(95% Conf. Int.) | P-Heterogeneity |
|---|---|---|---|---|
| **Cross-ancestry meta-analysis** | | | | |
| Oral cavity | 5596 | 37722 | 0.84(0.80 to 0.88) | |
| **Genetic ancestrt** | | | | |
| European | 3511 | 31833 | 0.83(0.78 to 0.87) | 0.32 |
| Mixed | 2085 | 5889 | 0.87(0.81 to 0.93) | |
| **Sex** | | | | |
| Men | 3749 | 19246 | 0.82(0.78 to 0.86) | 0.09 |
| Women | 1847 | 18476 | 0.89(0.82 to 0.96) | |
| **Smoking Status** | | | | |
| Never | 1498 | 15197 | 0.85(0.78 to 0.92) | 0.2 |
| Former | 1598 | 9741 | 0.80(0.74 to 0.87) | |
| Current | 2339 | 5675 | 0.89(0.82 to 0.96) | |
| **Drinking Status** | | | | |
| Never | 1218 | 3398 | 0.81(0.73 to 0.89) | 0.26 |
| Ever | 4098 | 26338 | 0.86(0.82 to 0.91) | |
| **Geographic Region** | | | | |
| Europe | 2337 | 30967 | 0.85(0.80 to 0.90) | 0.78 |
| North America | 1770 | 3694 | 0.84(0.77 to 0.91) | |
| South America | 1194 | 2324 | 0.89(0.80 to 0.98) | |
| Middle East | 153 | 504 | 0.95(0.72 to 1.24) | |
| South Asia | 142 | 233 | 0.93(0.69 to 1.24) | |
| **Subsite** | | | | |
| Larynx | 4409 | 37722 | 0.95(0.91 to 1.00) | 0.2 |
| Hypopharynx | 898 | 37722 | 0.92(0.83 to 1.01) | |
| Oropharynx (HPV-) | 1473 | 23132 | 0.87(0.80 to 0.94) | |
| Oropharynx (HPV+) | 2212 | 22803 | 0.90(0.84 to 0.96) | |

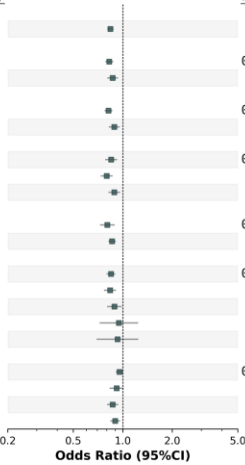


### rs407238

| Variable | NO.Case | NO.Control | OR.(95% Conf. Int.) | P-Heterogeneity |
|---|---|---|---|---|
| **Cross-ancestry meta-analysis** | | | | |
| Oral cavity | 5596 | 37722 | 1.15(1.10 to 1.20) | |
| **Genetic ancestry** | | | | |
| European | 3511 | 31833 | 1.20(1.14 to 1.27) | 0.004** |
| Mixed | 2085 | 5889 | 1.04(0.97 to 1.13) | |
| **Sex** | | | | |
| Men | 3749 | 19246 | 1.14(1.08 to 1.20) | 0.51 |
| Women | 1847 | 18476 | 1.17(1.09 to 1.27) | |
| **Smoking Status** | | | | |
| Never | 1498 | 15197 | 1.14(1.05 to 1.24) | 0.33 |
| Former | 1598 | 9741 | 1.19(1.10 to 1.29) | |
| Current | 2339 | 5675 | 1.09(1.00 to 1.18) | |
| **Drinking Status** | | | | |
| Never | 1218 | 3398 | 1.14(1.03 to 1.27) | 0.79 |
| Ever | 4098 | 26338 | 1.16(1.10 to 1.22) | |
| **Cluster** | | | | |
| AFR | 125 | 273 | 1.15(0.81 to 1.62) | 0.01* |
| SAS | 184 | 488 | 0.96(0.74 to 1.24) | |
| EAS | 28 | 24 | 0.76(0.27 to 2.15) | |
| EUR | 3511 | 31833 | 1.20(1.14 to 1.27) | |
| AMR | 6 | 29 | 3.33(0.60 to 18.43) | |
| Admixed | 1742 | 5075 | 1.02(0.94 to 1.10) | |
| **Geographic Region** | | | | |
| Europe | 2337 | 30967 | 1.18(1.11 to 1.25) | 0.37 |
| North America | 1770 | 3694 | 1.13(1.04 to 1.23) | |
| South America | 1194 | 2324 | 1.07(0.96 to 1.19) | |
| Middle East | 153 | 504 | 0.96(0.73 to 1.26) | |
| South Asia | 142 | 233 | 1.12(0.81 to 1.54) | |
| **Subsite** | | | | |
| Larynx | 4409 | 37722 | 1.08(1.02 to 1.14) | 0.01* |
| Hypopharynx | 332 | 5889 | 0.92(0.78 to 1.10) | |
| Oropharynx (HPV-) | 1473 | 23132 | 1.00(0.92 to 1.08) | |
| Oropharynx (HPV+) | 2212 | 22803 | 0.95(0.89 to 1.02) | |

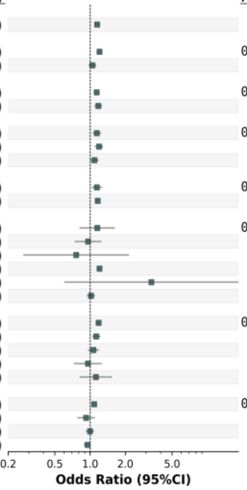

**b** **Cross-ancestry (oral cavity)**

## rs3731239

| Variable | NO.Case | NO.Control | OR.(95% Conf. Int.) | P-Heterogeneity |
|---|---|---|---|---|
| **Cross-ancestry meta-analysis** | | | | |
| Oral cavity | 5596 | 37722 | 1.14(1.09 to 1.19) | |
| **Genetic ancestry** | | | | |
| European | 3511 | 31833 | 1.14(1.08 to 1.20) | 1 |
| Mixed | 2085 | 5889 | 1.14(1.05 to 1.24) | |
| **Sex** | | | | |
| Men | 3749 | 19246 | 1.16(1.09 to 1.22) | 0.39 |
| Women | 1847 | 18476 | 1.11(1.02 to 1.20) | |
| **Smoking Status** | | | | |
| Never | 1498 | 15197 | 1.12(1.03 to 1.22) | 0.65 |
| Former | 1598 | 9741 | 1.16(1.07 to 1.27) | |
| Current | 2339 | 5675 | 1.19(1.08 to 1.30) | |
| **Drinking Status** | | | | |
| Never | 1218 | 3398 | 1.14(1.02 to 1.27) | 0.91 |
| Ever | 4098 | 26338 | 1.15(1.09 to 1.22) | |
| **Geographic Region** | | | | |
| Europe | 2337 | 30967 | 1.14(1.07 to 1.21) | 0.14 |
| North America | 1770 | 3694 | 1.17(1.07 to 1.27) | |
| South America | 1194 | 2324 | 1.21(1.08 to 1.36) | |
| Middle East | 153 | 504 | 0.88(0.65 to 1.19) | |
| South Asia | 142 | 233 | 0.83(0.57 to 1.21) | |
| **Subsite** | | | | |
| Larynx | 4409 | 37722 | 1.09(1.04 to 1.16) | 0.005** |
| Hypopharynx | 898 | 37722 | 0.93(0.83 to 1.03) | |
| Oropharynx (HPV-) | 1473 | 23132 | 1.08(1.00 to 1.18) | |
| Oropharynx (HPV+) | 2212 | 22803 | 0.97(0.91 to 1.04) | |

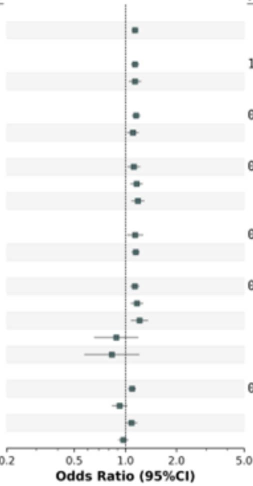


## rs12910284

| Variable | NO.Case | NO.Control | OR.(95% Conf. Int.) | P-Heterogeneity |
|---|---|---|---|---|
| **Cross-ancestry meta-analysis** | | | | |
| Oral cavity | 5596 | 37722 | 1.17(1.12 to 1.23) | |
| **Genetic ancestry** | | | | |
| European | 3511 | 31833 | 1.20(1.14 to 1.27) | 0.13 |
| Mixed | 2085 | 5889 | 1.11(1.03 to 1.21) | |
| **Sex** | | | | |
| Men | 3749 | 19246 | 1.19(1.13 to 1.26) | 0.25 |
| Women | 1847 | 18476 | 1.13(1.04 to 1.22) | |
| **Smoking Status** | | | | |
| Never | 1498 | 15197 | 1.20(1.10 to 1.30) | 0.7 |
| Former | 1598 | 9741 | 1.14(1.05 to 1.24) | |
| Current | 2339 | 5675 | 1.19(1.09 to 1.29) | |
| **Drinking Status** | | | | |
| Never | 1218 | 3398 | 1.12(1.00 to 1.24) | 0.42 |
| Ever | 4098 | 26338 | 1.17(1.11 to 1.24) | |
| **Geographic Region** | | | | |
| Europe | 2337 | 30967 | 1.15(1.08 to 1.22) | 0.92 |
| North America | 1770 | 3694 | 1.18(1.08 to 1.28) | |
| South America | 1194 | 2324 | 1.11(0.99 to 1.24) | |
| Middle East | 153 | 504 | 1.07(0.79 to 1.44) | |
| South Asia | 142 | 233 | 1.13(0.80 to 1.59) | |
| **Subsite** | | | | |
| Larynx | 4409 | 37722 | 1.02(0.97 to 1.07) | 0.11 |
| Hypopharynx | 898 | 37722 | 0.98(0.89 to 1.09) | |
| Oropharynx (HPV-) | 1473 | 23132 | 1.12(1.03 to 1.21) | |
| Oropharynx (HPV+) | 2212 | 22803 | 1.00(0.93 to 1.07) | |

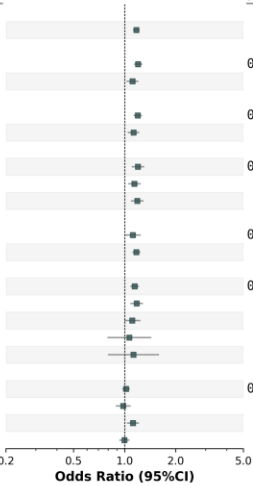


## rs67351073

| Variable | NO.Case | NO.Control | OR.(95% Conf. Int.) | P-Heterogeneity |
|---|---|---|---|---|
| **Cross-ancestry meta-analysis** | | | | |
| Oral cavity | 5596 | 37722 | 0.78(0.72 to 0.85) | |
| **Genetic ancestry** | | | | |
| European | 3511 | 31833 | 0.75(0.67 to 0.83) | 0.13 |
| Mixed | 2085 | 5889 | 0.86(0.74 to 1.01) | |
| **Sex** | | | | |
| Men | 3749 | 19246 | 0.76(0.69 to 0.85) | 0.61 |
| Women | 1847 | 18476 | 0.80(0.69 to 0.93) | |
| **Smoking Status** | | | | |
| Never | 1498 | 15197 | 0.76(0.64 to 0.90) | 0.86 |
| Former | 1598 | 9741 | 0.79(0.68 to 0.93) | |
| Current | 2339 | 5675 | 0.75(0.63 to 0.88) | |
| **Drinking Status** | | | | |
| Never | 1218 | 3398 | 0.91(0.74 to 1.11) | 0.15 |
| Ever | 4098 | 26338 | 0.77(0.69 to 0.85) | |
| **Geographic Region** | | | | |
| Europe | 2337 | 30967 | 0.76(0.67 to 0.86) | |
| North America | 1770 | 3694 | 0.76(0.65 to 0.89) | 0.71 |
| South America | 1194 | 2324 | 0.85(0.69 to 1.05) | |
| Middle East | 153 | 504 | 1.13(0.54 to 2.37) | |
| South Asia | 142 | 233 | 0.63(0.31 to 1.28) | |
| **Subsite** | | | | |
| Larynx | 4409 | 37722 | 0.99(0.90 to 1.10) | 0.42 |
| Hypopharynx | 898 | 37722 | 0.91(0.75 to 1.10) | |
| Oropharynx (HPV-) | 1473 | 23132 | 0.94(0.81 to 1.09) | |
| Oropharynx (HPV+) | 2212 | 22803 | 1.07(0.95 to 1.20) | |

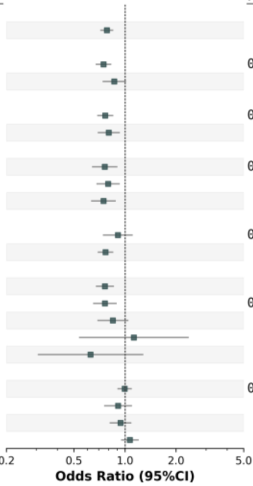


## rs577454702

| Variable | NO.Case | NO.Control | OR.(95% Conf. Int.) | P-Heterogeneity |
|---|---|---|---|---|
| **Cross-ancestry meta-analysis** | | | | |
| Oral cavity | 5596 | 37722 | 2.51(1.86 to 3.39) | |
| **Genetic ancestry** | | | | |
| European | 3511 | 31833 | 2.60(1.86 to 3.65) | 0.64 |
| Mixed | 2085 | 5889 | 2.18(1.13 to 4.22) | |
| **Sex** | | | | |
| Men | 3749 | 19246 | 2.29(1.56 to 3.37) | 0.49 |
| Women | 1847 | 18476 | 2.84(1.75 to 4.61) | |
| **Smoking Status** | | | | |
| Never | 1498 | 15197 | 3.31(2.00 to 5.46) | 0.51 |
| Former | 1598 | 9741 | 2.15(1.23 to 3.77) | |
| Current | 2339 | 5675 | 2.46(1.29 to 4.69) | |
| **Drinking Status** | | | | |
| Never | 1218 | 3398 | 2.90(1.31 to 6.43) | 0.96 |
| Ever | 4098 | 26338 | 2.96(2.06 to 4.27) | |
| **Geographic Region** | | | | |
| Europe | 2337 | 30967 | 2.24(1.56 to 3.22) | 0.94 |
| North America | 1770 | 3694 | 2.02(1.11 to 3.69) | |
| South America | 1123 | 2239 | 2.36(1.04 to 5.33) | |
| **Subsite** | | | | |
| Larynx | 4409 | 37722 | 1.04(0.69 to 1.58) | 0.55 |
| Hypopharynx | 898 | 37722 | 1.48(0.65 to 3.33) | |
| Oropharynx (HPV-) | 1473 | 23132 | 1.32(0.67 to 2.60) | |
| Oropharynx (HPV+) | 1980 | 18166 | 1.68(0.99 to 2.84) | |

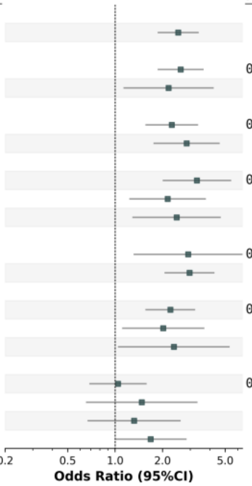

**c** **Cross-ancestry (larynx)**

## rs10774632

| Variable | NO.Case | NO.Control | OR.(95% Conf. Int.) | P-Heterogeneity |
|---|---|---|---|---|
| **Cross-ancestry meta-analysis** | | | | |
| Larynx | 4409 | 37722 | 1.20(1.13 to 1.28) | |
| **Genetic ancestry** | | | | |
| European | 2787 | 31833 | 1.24(1.15 to 1.34) | 0.2 |
| Mixed | 1622 | 5889 | 1.14(1.03 to 1.26) | |
| **Sex** | | | | |
| Men | 3826 | 19246 | 1.17(1.09 to 1.25) | 0.12 |
| Women | 583 | 18476 | 1.33(1.15 to 1.55) | |
| **Smoking Status** | | | | |
| Never | 552 | 15197 | 1.09(0.90 to 1.32) | 0.55 |
| Former | 1726 | 9741 | 1.20(1.07 to 1.33) | |
| Current | 2114 | 5675 | 1.24(1.11 to 1.38) | |
| **Drinking Status** | | | | |
| Never | 774 | 3398 | 1.17(1.00 to 1.37) | 0.86 |
| Ever | 3366 | 26338 | 1.19(1.10 to 1.28) | |
| **Geographic Region** | | | | |
| Europe | 2003 | 30967 | 1.18(1.08 to 1.28) | 0.16 |
| North America | 1354 | 3694 | 1.22(1.09 to 1.37) | |
| South America | 770 | 2324 | 1.01(0.89 to 1.15) | |
| Middle East | 282 | 504 | 1.11(0.86 to 1.44) | |
| **Subsite** | | | | |
| Oral cavity | 5596 | 37722 | 1.04(0.98 to 1.09) | 0.0003*** |
| Hypopharynx | 898 | 37722 | 1.20(1.07 to 1.34) | |
| Oropharynx (HPV-) | 1473 | 23132 | 1.23(1.12 to 1.34) | |
| Oropharynx (HPV+) | 2212 | 22803 | 0.96(0.88 to 1.05) | |

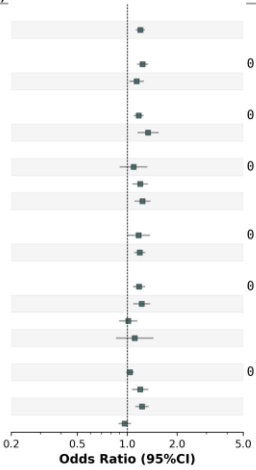


## rs11571833

| Variable | NO.Case | NO.Control | OR.(95% Conf. Int.) | P-Heterogeneity |
|---|---|---|---|---|
| **Cross-ancestry meta-analysis** | | | | |
| Larynx | 4409 | 37722 | 2.09(1.65 to 2.66) | |
| **Genetic ancestry** | | | | |
| European | 2787 | 31833 | 2.12(1.62 to 2.78) | 0.81 |
| Mixed | 1622 | 5889 | 1.98(1.17 to 3.34) | |
| **Sex** | | | | |
| Men | 3826 | 19246 | 2.16(1.66 to 2.81) | 0.69 |
| Women | 583 | 18476 | 1.89(1.02 to 3.51) | |
| **Smoking Status** | | | | |
| Never | 552 | 15197 | 3.02(1.73 to 5.28) | 0.35 |
| Former | 1726 | 9741 | 1.89(1.18 to 3.04) | |
| Current | 2114 | 5675 | 2.91(1.74 to 4.86) | |
| **Drinking Status** | | | | |
| Never | 774 | 3398 | 2.06(1.08 to 3.95) | 0.73 |
| Ever | 3366 | 26338 | 2.34(1.70 to 3.22) | |
| **Geographic Region** | | | | |
| Europe | 2003 | 30967 | 2.00(1.51 to 2.64) | |
| North America | 1354 | 3694 | 2.20(1.42 to 3.41) | 0.87 |
| South America | 770 | 2324 | 1.75(0.87 to 3.49) | |
| Middle East | 282 | 504 | 1.37(0.42 to 4.48) | |
| **Subsite** | | | | |
| Oral cavity | 5596 | 37722 | 1.63(1.32 to 2.02) | 0.009** |
| Hypopharynx | 898 | 37722 | 2.73(1.91 to 3.90) | |
| Oropharynx (HPV-) | 1473 | 23132 | 2.48(1.82 to 3.38) | |
| Oropharynx (HPV+) | 2212 | 22803 | 1.45(1.06 to 1.97) | |

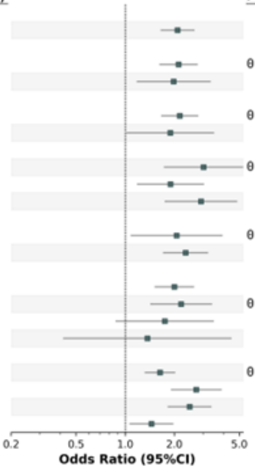


## rs10419397

| Variable | NO.Case | NO.Control | OR.(95% Conf. Int.) | P-Heterogeneity |
|---|---|---|---|---|
| **Cross-ancestry meta-analysis** | | | | |
| Larynx | 4409 | 37722 | 1.17(1.11 to 1.24) | |
| **Genetic ancestry** | | | | |
| European | 2787 | 31833 | 1.18(1.10 to 1.26) | 0.66 |
| Mixed | 1622 | 5889 | 1.15(1.03 to 1.27) | |
| **Sex** | | | | |
| Men | 3826 | 19246 | 1.16(1.09 to 1.24) | 0.57 |
| Women | 583 | 18476 | 1.22(1.06 to 1.40) | |
| **Smoking Status** | | | | |
| Never | 552 | 15197 | 1.19(1.00 to 1.43) | 0.8 |
| Former | 1726 | 9741 | 1.14(1.03 to 1.26) | |
| Current | 2114 | 5675 | 1.19(1.08 to 1.31) | |
| **Drinking Status** | | | | |
| Never | 774 | 3398 | 1.13(0.96 to 1.32) | 0.81 |
| Ever | 3366 | 26338 | 1.15(1.07 to 1.24) | |
| **Geographic Region** | | | | |
| Europe | 2003 | 30967 | 1.19(1.11 to 1.27) | 0.78 |
| North America | 1354 | 3694 | 1.19(1.08 to 1.32) | |
| South America | 770 | 2324 | 1.10(0.96 to 1.26) | |
| Middle East | 282 | 504 | 1.17(0.90 to 1.52) | |
| **Subsite** | | | | |
| Oral cavity | 5596 | 37722 | 1.09(1.04 to 1.14) | 0.59 |
| Hypopharynx | 898 | 37722 | 1.17(1.05 to 1.30) | |
| Oropharynx (HPV-) | 1473 | 23132 | 1.07(0.98 to 1.17) | |
| Oropharynx (HPV+) | 2212 | 22803 | 1.09(1.01 to 1.17) | |

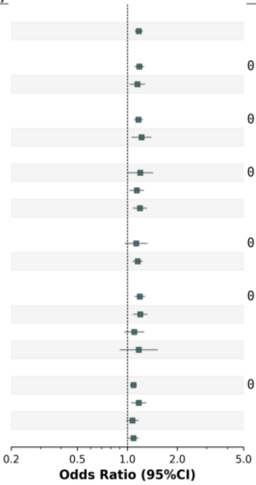

**d** **Cross-ancestry (hypopharynx)**

## rs1154462

| Variable | NO.Case | NO.Control | OR.(95% Conf. Int.) | P-Heterogeneity |
|---|---|---|---|---|
| **Cross-ancestry meta-analysis** | | | | |
| Hypopharynx | 898 | 37722 | 1.34(1.22 to 1.49) | |
| **Genetic ancestry** | | | | |
| European | 566 | 31833 | 1.38(1.22 to 1.56) | 0.52 |
| Mixed | 332 | 5889 | 1.28(1.08 to 1.53) | |
| **Sex** | | | | |
| Men | 759 | 19246 | 1.38(1.24 to 1.55) | 0.2 |
| Women | 139 | 18476 | 1.16(0.90 to 1.49) | |
| **Smoking Status** | | | | |
| Never | 151 | 15197 | 1.40(1.08 to 1.81) | 0.05 |
| Former | 297 | 9741 | 1.10(0.92 to 1.32) | |
| Current | 439 | 5675 | 1.47(1.26 to 1.72) | |
| **Drinking Status** | | | | |
| Never | 99 | 3398 | 1.56(1.13 to 2.16) | 0.28 |
| Ever | 739 | 26338 | 1.30(1.16 to 1.46) | |
| **Geographic Region** | | | | |
| Europe | 426 | 30967 | 1.30(1.13 to 1.49) | 0.13 |
| North America | 238 | 3694 | 1.64(1.34 to 2.01) | |
| South America | 188 | 2324 | 1.16(0.91 to 1.47) | |
| Middle East | 46 | 504 | 1.50(0.95 to 2.37) | |
| **Subsite** | | | | |
| Oral cavity | 5596 | 37722 | 1.07(1.02 to 1.12) | 0.41 |
| Larynx | 4409 | 37722 | 1.05(1.00 to 1.11) | |
| Oropharynx (HPV-) | 1473 | 23132 | 1.13(1.04 to 1.23) | |
| Oropharynx (HPV+) | 2212 | 22803 | 1.03(0.96 to 1.11) | |

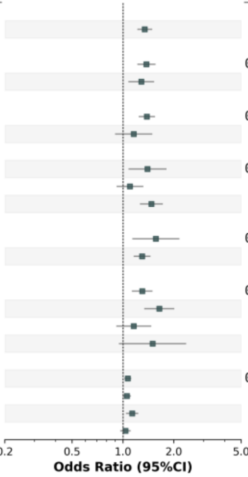


## rs11571815

| Variable | NO.Case | NO.Control | OR.(95% Conf. Int.) | P-Heterogeneity |
|---|---|---|---|---|
| **Cross-ancestry meta-analysis** | | | | |
| Hypopharynx | 898 | 37722 | 2.73(1.91 to 3.90) | |
| **Genetic ancestry** | | | | |
| European | 566 | 31833 | 2.93(1.97 to 4.37) | 0.42 |
| Mixed | 332 | 5889 | 2.03(0.91 to 4.54) | |
| **Sex** | | | | |
| Men | 759 | 19246 | 2.67(1.80 to 3.97) | 0.8 |
| Women | 139 | 18476 | 3.00(1.29 to 6.98) | |
| **Smoking Status** | | | | |
| Never | 151 | 15197 | 1.80(0.53 to 6.10) | 0.58 |
| Former | 297 | 9741 | 3.65(1.99 to 6.71) | |
| Current | 439 | 5675 | 3.54(1.90 to 6.59) | |
| **Drinking Status** | | | | |
| Never | 99 | 3398 | 2.47(0.75 to 8.12) | 0.81 |
| Ever | 739 | 26338 | 2.89(1.92 to 4.35) | |
| **Geographic Region** | | | | |
| Europe | 426 | 30967 | 2.90(1.83 to 4.60) | 0.62 |
| North America | 179 | 2937 | 4.16(2.07 to 8.38) | |
| South America | 184 | 2239 | 2.33(0.78 to 6.92) | |
| Middle East | 46 | 504 | 1.18(0.13 to 10.32) | |
| **Subsite** | | | | |
| Oral cavity | 5596 | 37722 | 1.63(1.32 to 2.02) | 0.04* |
| Larynx | 4409 | 37722 | 2.09(1.64 to 2.66) | |
| Oropharynx (HPV-) | 1473 | 23132 | 2.48(1.82 to 3.37) | |
| Oropharynx (HPV+) | 2212 | 22803 | 1.45(1.06 to 1.97) | |

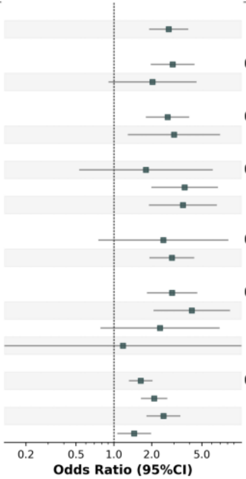


## rs181194133

| Variable | NO.Case | NO.Control | OR.(95% Conf. Int.) | P-Heterogeneity |
|---|---|---|---|---|
| **Cross-ancestry meta-analysis** | | | | |
| Hypopharynx | 898 | 37722 | 3.24(2.12 to 4.96) | |
| **Genetic ancestry** | | | | |
| European | 566 | 31833 | 3.44(2.23 to 5.31) | 0.15 |
| Mixed | 332 | 5889 | 0.66(0.07 to 6.13) | |
| **Sex** | | | | |
| Men | 759 | 19246 | 2.98(1.79 to 4.99) | 0.39 |
| Women | 139 | 18476 | 4.45(2.09 to 9.46) | |
| **Smoking Status** | | | | |
| Never | 151 | 15197 | 3.15(1.23 to 8.07) | 0.96 |
| Former | 206 | 8018 | 3.75(1.73 to 8.13) | |
| Current | 439 | 5675 | 3.58(1.69 to 7.59) | |
| **Drinking Status** | | | | |
| Never | 99 | 3398 | 3.48(0.78 to 15.60) | 0.92 |
| Ever | 463 | 22669 | 3.77(2.30 to 6.17) | |
| **Geographic Region** | | | | |
| Europe | 383 | 28811 | 2.53(1.46 to 4.38) | 0.16 |
| North America | 179 | 2937 | 6.86(3.09 to 15.23) | |
| Middle East | 46 | 504 | 2.85(0.41 to 19.75) | |
| **Subsite** | | | | |
| Larynx | 4409 | 37722 | 1.10(0.79 to 1.52) | 0.97 |
| Oral cavity | 5596 | 37722 | 1.22(0.92 to 1.62) | |
| Oropharynx (HPV-) | 1473 | 23132 | 1.19(0.72 to 1.99) | |
| Oropharynx (HPV+) | 2212 | 22803 | 1.15(0.78 to 1.70) | |

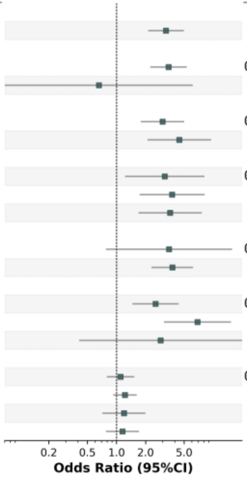

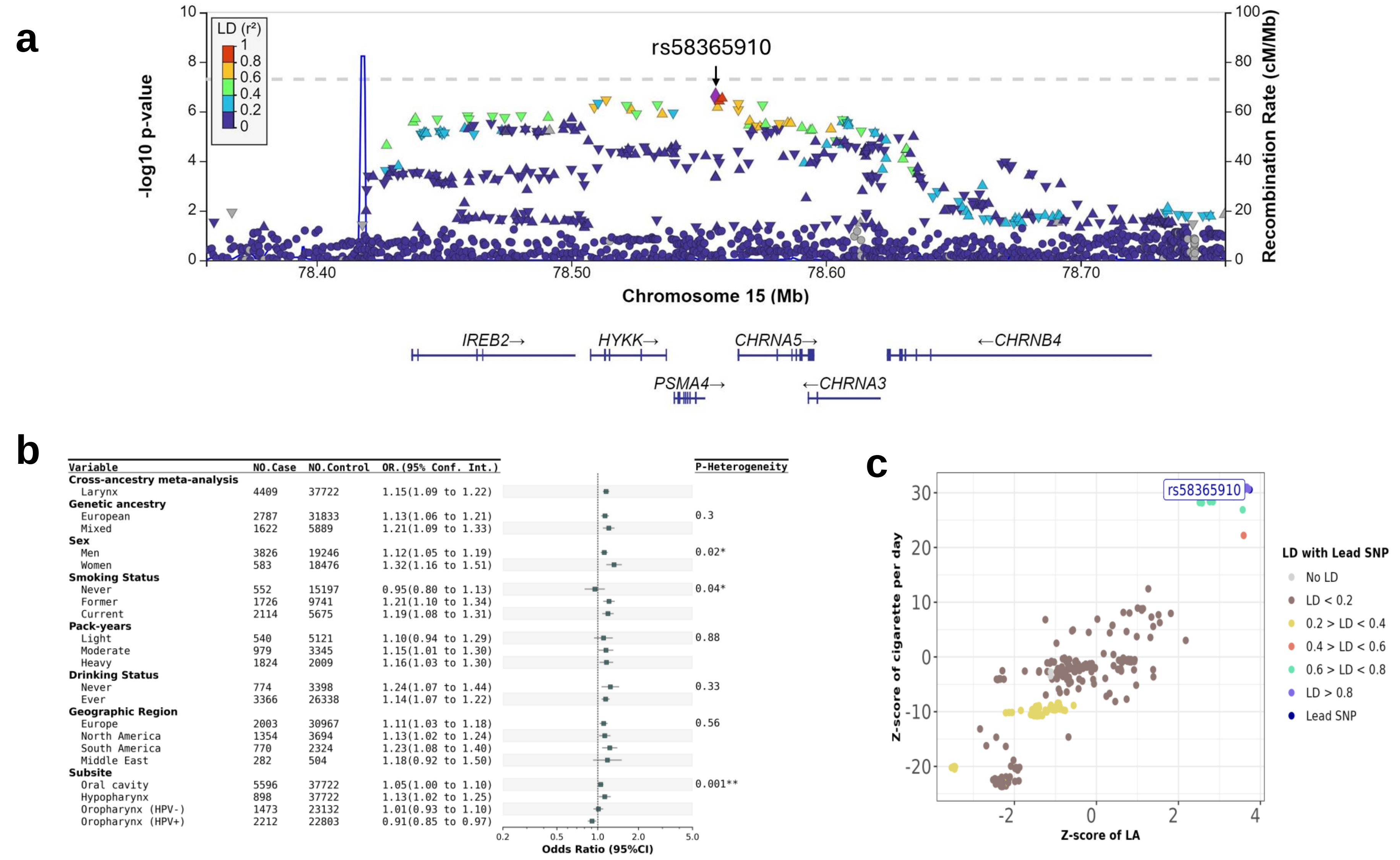


**Supplementary Figure 8. rs58365910 shows a suggestive association with laryngeal cancer at the 15q25 locus.** a) Regional plot of rs58365910, an intergenic variant mapped to *CHRNA5/PSMA4* , *showed a* suggestive association with laryngeal cancer ($p_{meta}$=2.53x$10^{-7}$). b) Forest plot of Odds ratios (ORs) for rs58365910 stratified by sex, smoking status, pack-year, drinking status, and geographic region. c) Forest plot presents the ORs for laryngeal cancer risk associated with different combinations of smoking and drinking behaviors in meta-analysis, European- and Mixed group GWAS analyses. d) rs58365910 was colocalized with cigarette-smoked per day (CPD) (PP4 score=0.89).

**a** **Cross-ancestry (all sites combined)**

Chr6:33046667

| Variable | NO.Case | NO.Control | OR(95% Conf. Int.) | P-Heterogeneity |
|---|---|---|---|---|
| **Cross-ancestry meta-analysis** | | | | |
| All sites combined | 18865 | 36207 | 1.10(1.07 to 1.14) | |
| **Genetic ancestry** | | | | |
| Admixed | 4300 | 4520 | 1.06(0.98 to 1.13) | 0.63 |
| African | 403 | 280 | 1.13(0.87 to 1.47) | |
| European | 13360 | 30277 | 1.12(1.08 to 1.17) | |
| Middle East | 491 | 479 | 1.07(0.87 to 1.32) | |
| South Asian | 311 | 651 | 1.04(0.80 to 1.36) | |
| **Drinking Status** | | | | |
| Ever | 12722 | 30490 | 1.09(1.05 to 1.14) | 0.99 |
| Never | 2855 | 3049 | 1.09(0.99 to 1.20) | |
| **Sex** | | | | |
| Female | 4136 | 16753 | 1.07(0.97 to 1.19) | 0.59 |
| Male | 14729 | 19454 | 1.11(1.06 to 1.15) | |
| **Smoking Status** | | | | |
| Current | 6854 | 5055 | 1.07(0.98 to 1.17) | 0.76 |
| Former | 5749 | 11655 | 1.11(1.05 to 1.18) | |
| Never | 3779 | 17626 | 1.09(1.00 to 1.20) | |
| **Subsite** | | | | |
| Hypopharynx | 798 | 34797 | 1.07(0.95 to 1.21) | 0.01 |
| Larynx | 4357 | 35556 | 1.08(1.02 to 1.15) | |
| Oral cavity | 5536 | 36207 | 1.04(0.98 to 1.09) | |
| Oropharynx (HPV-) | 1452 | 35077 | 1.17(1.06 to 1.28) | |
| Oropharynx (HPV+) | 2182 | 34797 | 1.22(1.12 to 1.33) | |
| **GeographicRegion** | | | | |
| Europe | 7716 | 29343 | 1.04(0.89 to 1.22) | 0.76 |
| NorthAmerica | 7393 | 3817 | 1.11(1.04 to 1.19) | |
| SouthAmerica | 2973 | 2294 | 1.09(1.00 to 1.19) | |

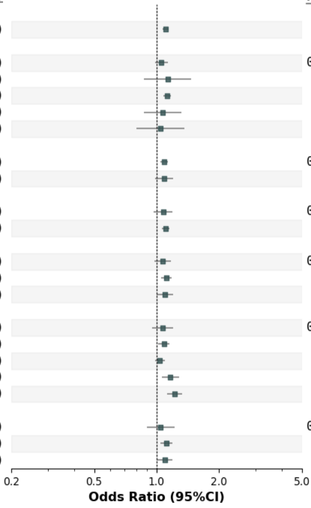


rs28360051

| Variable | NO.Case | NO.Control | OR(95% Conf. Int.) | P-Heterogeneity |
|---|---|---|---|---|
| **Cross-ancestry meta-analysis** | | | | |
| All sites combined | 18865 | 36207 | 1.23(1.14 to 1.33) | |
| **Genetic ancestry** | | | | |
| Admixed | 4300 | 4520 | 1.13(0.93 to 1.37) | 0.56 |
| African | 403 | 280 | 1.00(0.39 to 2.56) | |
| European | 13360 | 30277 | 1.26(1.15 to 1.37) | |
| **Drinking Status** | | | | |
| Ever | 12722 | 30490 | 1.14(0.95 to 1.37) | 0.78 |
| Never | 2855 | 3049 | 1.20(0.90 to 1.58) | |
| **Sex** | | | | |
| Female | 4136 | 16753 | 1.29(1.01 to 1.66) | 0.49 |
| Male | 14729 | 19454 | 1.15(0.92 to 1.44) | |
| **Smoking Status** | | | | |
| Current | 6854 | 5055 | 1.22(1.02 to 1.47) | 0.41 |
| Former | 5749 | 11655 | 1.11(0.86 to 1.43) | |
| Never | 3779 | 17626 | 1.34(1.16 to 1.54) | |
| **Subsite** | | | | |
| Hypopharynx | 798 | 34797 | 1.02(0.47 to 2.25) | 0.01 |
| Larynx | 4357 | 35556 | 1.06(0.91 to 1.22) | |
| Oral cavity | 5536 | 36207 | 1.11(0.98 to 1.26) | |
| Oropharynx (HPV-) | 1452 | 35077 | 1.04(0.68 to 1.59) | |
| Oropharynx (HPV+) | 2182 | 34797 | 1.54(1.31 to 1.80) | |
| **GeographicRegion** | | | | |
| Europe | 7716 | 29343 | 1.29(1.16 to 1.42) | 0.25 |
| NorthAmerica | 7393 | 3817 | 1.54(0.93 to 2.53) | |
| SouthAmerica | 2973 | 2294 | 0.79(0.42 to 1.49) | |

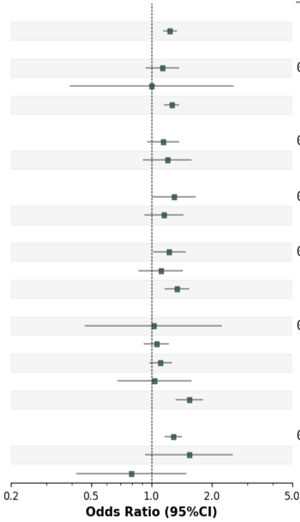


**Supplementary Figure 9. Forest plots of novel variants from HLA fine-mapping analyses.**
For each independent novel top hit identified in the cross-ancestry HLA fine-mapping and ancestry-specific, analyses were stratified by sex, smoking status, drinking status, geographic region, and cancer subsites. Forest plots of variants identified by cancer subsite: a) top hit variants identified in all sites combined, b) variants identified in the HPV- oropharynx, c) variants identified in HPV+ oropharynx, d) variants identified in all sites combined specific to admixed population, e) variants identified in oral cavity specific to European population, and f) variants identified in HPV+ oropharynx specific to European population.

**b** **Cross-ancestry (HPV- oropharynx)**

rs1131212

| Variable | NO.Case | NO.Control | OR(95% Conf. Int.) | P-Heterogeneity |
|---|---|---|---|---|
| **Cross-ancestry meta-analysis** | | | | |
| Oropharynx (HPV-) | 1452 | 35077 | 1.33(1.19 to 1.48) | |
| **Genetic ancestry** | | | | |
| Admixed | 453 | 4520 | 1.33(1.06 to 1.66) | 0.62 |
| African | 65 | 280 | 1.03(0.61 to 1.75) | |
| European | 934 | 30277 | 1.35(1.18 to 1.54) | |
| **Drinking Status** | | | | |
| Ever | 1156 | 30134 | 1.31(1.08 to 1.58) | |
| **Sex** | | | | |
| Female | 298 | 16273 | 1.61(1.09 to 2.39) | 0.29 |
| Male | 1140 | 18708 | 1.29(1.14 to 1.46) | |
| **Smoking Status** | | | | |
| Current | 785 | 4850 | 1.30(1.10 to 1.54) | 0.63 |
| Former | 411 | 11432 | 1.39(1.10 to 1.76) | |
| **Subsite** | | | | |
| Hypopharynx | 798 | 34797 | 1.18(1.03 to 1.36) | 0.00 |
| Larynx | 4357 | 35556 | 1.08(1.01 to 1.16) | |
| Oral cavity | 5536 | 36207 | 1.15(1.08 to 1.22) | |
| Oropharynx (HPV-) | 1452 | 35077 | 1.33(1.19 to 1.48) | |
| Oropharynx (HPV+) | 2182 | 34797 | 0.94(0.87 to 1.03) | |
| **GeographicRegion** | | | | |
| Europe | 679 | 28929 | 1.46(1.24 to 1.71) | 0.69 |
| NorthAmerica | 407 | 3817 | 1.31(0.81 to 2.10) | |
| SouthAmerica | 352 | 2234 | 7.53(0.10 to 544.21) | |

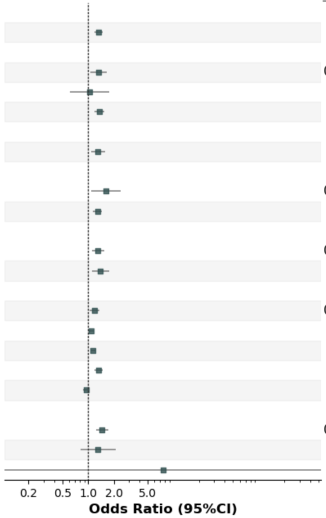

### c Cross-ancestry (HPV+ oropharynx)

#### DRB1 37Asn/Ser

| Variable | NO.Case | NO.Control | OR(95% Conf. Int.) | P-Heterogeneity |
|---|---|---|---|---|
| **Cross-ancestry meta-analysis** | | | | |
| Oropharynx (HPV+) | 2182 | 34797 | 0.68(0.63 to 0.73) | |
| **Genetic ancestry** | | | | |
| Admixed | 222 | 4520 | 0.73(0.59 to 0.91) | 0.43 |
| European | 1960 | 30277 | 0.67(0.62 to 0.73) | |
| **Drinking Status** | | | | |
| Ever | 1517 | 29911 | 0.69(0.63 to 0.76) | |
| **Sex** | | | | |
| Male | 1815 | 18524 | 0.70(0.64 to 0.76) | |
| **Smoking Status** | | | | |
| Former | 987 | 11432 | 0.70(0.54 to 0.90) | 0.75 |
| Never | 869 | 16855 | 0.73(0.64 to 0.82) | |
| **Subsite** | | | | |
| Hypopharynx | 798 | 34797 | 0.90(0.80 to 1.00) | 0.00 |
| Larynx | 4357 | 35556 | 1.04(0.92 to 1.16) | |
| Oral cavity | 5536 | 36207 | 0.97(0.92 to 1.02) | |
| Oropharynx (HPV-) | 1452 | 35077 | 0.85(0.75 to 0.97) | |
| Oropharynx (HPV+) | 2182 | 34797 | 0.68(0.63 to 0.73) | |
| **GeographicRegion** | | | | |
| Europe | 1086 | 28929 | 0.70(0.48 to 1.03) | 0.96 |
| NorthAmerica | 989 | 3633 | 0.71(0.62 to 0.80) | |

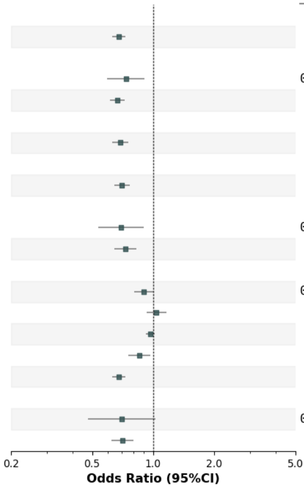


### rs4143334

| Variable | NO.Case | NO.Control | OR(95% Conf. Int.) | P-Heterogeneity |
|---|---|---|---|---|
| **Cross-ancestry meta-analysis** | | | | |
| Oropharynx (HPV+) | 2182 | 34797 | 1.89(1.51 to 2.35) | |
| **Genetic ancestry** | | | | |
| Admixed | 222 | 4520 | 1.55(0.81 to 2.96) | 0.53 |
| European | 1960 | 30277 | 1.93(1.53 to 2.45) | |
| **Drinking Status** | | | | |
| Ever | 1517 | 29911 | 1.96(1.50 to 2.56) | |
| **Sex** | | | | |
| Male | 1815 | 18524 | 1.84(1.44 to 2.36) | |
| **Smoking Status** | | | | |
| Former | 987 | 11432 | 1.84(0.73 to 4.58) | 0.77 |
| Never | 869 | 16855 | 2.15(1.23 to 3.75) | |
| **Subsite** | | | | |
| Hypopharynx | 798 | 34797 | 1.14(0.79 to 1.64) | 0.00 |
| Larynx | 4357 | 35556 | 0.96(0.80 to 1.15) | |
| Oral cavity | 5536 | 36207 | 0.95(0.80 to 1.13) | |
| Oropharynx (HPV-) | 1452 | 35077 | 0.79(0.56 to 1.10) | |
| Oropharynx (HPV+) | 2182 | 34797 | 1.89(1.51 to 2.35) | |
| **GeographicRegion** | | | | |
| Europe | 1086 | 28929 | 1.91(1.40 to 2.60) | 0.85 |
| NorthAmerica | 989 | 3633 | 1.83(1.27 to 2.63) | |

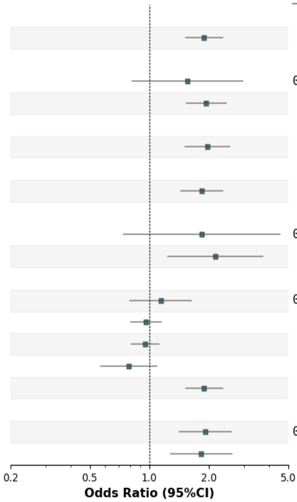


#### DRB1 233Thr

| Variable | NO.Case | NO.Control | OR(95% Conf. Int.) | P-Heterogeneity |
|---|---|---|---|---|
| **Cross-ancestry meta-analysis** | | | | |
| Oropharynx (HPV+) | 2182 | 34797 | 1.27(1.17 to 1.38) | |
| **Genetic ancestry** | | | | |
| Admixed | 222 | 4520 | 1.16(0.94 to 1.45) | 0.39 |
| European | 1960 | 30277 | 1.29(1.18 to 1.41) | |
| **Drinking Status** | | | | |
| Ever | 1517 | 29911 | 1.20(0.99 to 1.47) | |
| **Sex** | | | | |
| Male | 1815 | 18524 | 1.27(1.16 to 1.39) | |
| **Smoking Status** | | | | |
| Former | 987 | 11432 | 1.26(0.96 to 1.65) | 0.95 |
| Never | 869 | 16855 | 1.25(1.09 to 1.42) | |
| **Subsite** | | | | |
| Hypopharynx | 798 | 34797 | 0.95(0.85 to 1.07) | 0.00 |
| Larynx | 4357 | 35556 | 0.98(0.92 to 1.03) | |
| Oral cavity | 5536 | 36207 | 1.00(0.92 to 1.09) | |
| Oropharynx (HPV-) | 1452 | 35077 | 0.93(0.80 to 1.07) | |
| Oropharynx (HPV+) | 2182 | 34797 | 1.27(1.17 to 1.38) | |
| **GeographicRegion** | | | | |
| Europe | 1086 | 28929 | 1.35(1.04 to 1.76) | 0.32 |
| NorthAmerica | 989 | 3633 | 1.17(1.03 to 1.33) | |

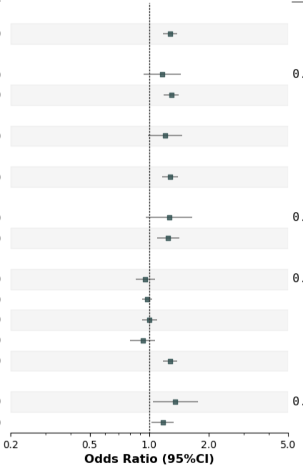


#### B 67Cys/Ser/Tyr

| Variable | NO.Case | NO.Control | OR(95% Conf. Int.) | P-Heterogeneity |
|---|---|---|---|---|
| **Cross-ancestry meta-analysis** | | | | |
| Oropharynx (HPV+) | 2182 | 34797 | 0.81(0.74 to 0.88) | |
| **Genetic ancestry** | | | | |
| Admixed | 222 | 4520 | 0.83(0.66 to 1.04) | 0.85 |
| European | 1960 | 30277 | 0.81(0.73 to 0.88) | |
| **Drinking Status** | | | | |
| Ever | 1517 | 29911 | 0.78(0.71 to 0.87) | |
| **Sex** | | | | |
| Male | 1815 | 18524 | 0.81(0.74 to 0.89) | |
| **Smoking Status** | | | | |
| Former | 987 | 11432 | 0.76(0.67 to 0.87) | 0.44 |
| Never | 869 | 16855 | 0.82(0.72 to 0.94) | |
| **Subsite** | | | | |
| Hypopharynx | 798 | 34797 | 0.90(0.80 to 1.02) | 0.00 |
| Larynx | 4357 | 35556 | 0.98(0.92 to 1.04) | |
| Oral cavity | 5536 | 36207 | 0.87(0.82 to 0.92) | |
| Oropharynx (HPV-) | 1452 | 35077 | 0.95(0.82 to 1.09) | |
| Oropharynx (HPV+) | 2182 | 34797 | 0.81(0.74 to 0.88) | |
| **GeographicRegion** | | | | |
| Europe | 1086 | 28929 | 0.79(0.70 to 0.89) | 0.76 |
| NorthAmerica | 989 | 3633 | 0.81(0.68 to 0.97) | |

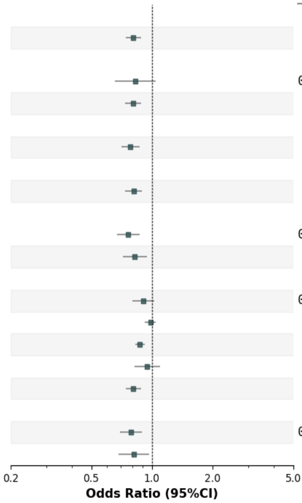

**d** **Admixed (all sites combined)**

rs1536036

| Variable | NO.Case | NO.Control | OR(95% Conf. Int.) | P-Heterogeneity |
|---|---|---|---|---|
| **Cross-ancestry meta-analysis** | | | | |
| All sites combined | 18865 | 36207 | 0.99(0.88 to 1.12) | |
| **Genetic ancestry** | | | | |
| Admixed | 4300 | 4520 | 0.85(0.80 to 0.91) | 0.00 |
| African | 403 | 280 | 1.42(1.12 to 1.79) | |
| European | 13360 | 30277 | 0.99(0.95 to 1.02) | |
| Middle East | 491 | 479 | 0.89(0.73 to 1.09) | |
| South Asian | 311 | 651 | 1.09(0.86 to 1.39) | |
| **Drinking Status** | | | | |
| Ever | 12722 | 30490 | 0.98(0.86 to 1.12) | 0.85 |
| Never | 2953 | 3106 | 0.96(0.82 to 1.13) | |
| **Sex** | | | | |
| Female | 4136 | 16753 | 0.97(0.87 to 1.08) | 0.73 |
| Male | 14729 | 19454 | 1.00(0.86 to 1.15) | |
| **Smoking Status** | | | | |
| Former | 5749 | 11655 | 0.97(0.77 to 1.24) | 0.92 |
| Never | 3810 | 17741 | 0.97(0.91 to 1.02) | |
| **Subsite** | | | | |
| Hypopharynx | 872 | 35556 | 0.93(0.77 to 1.12) | 0.89 |
| Larynx | 4374 | 36207 | 0.95(0.81 to 1.11) | |
| Oral cavity | 5536 | 36207 | 0.99(0.89 to 1.11) | |
| Oropharynx (HPV-) | 1452 | 35077 | 1.01(0.78 to 1.31) | |
| Oropharynx (HPV+) | 2182 | 34797 | 0.92(0.82 to 1.03) | |

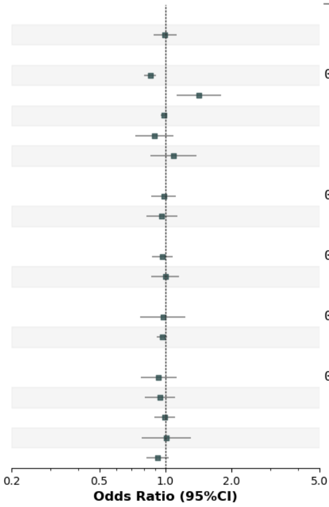

**e** **European (Oral cavity)**

### DRB1 74Ala/Leu/Del

| Variable | NO.Case | NO.Control | OR(95% Conf. Int.) | P-Heterogeneity |
|---|---|---|---|---|
| **Cross-ancestry meta-analysis** | | | | |
| OralCavity | 5536 | 36207 | 0.86(0.75 to 1.00) | |
| **Genetic ancestry** | | | | |
| Admixed | 1574 | 4520 | 1.02(0.92 to 1.12) | 0.00 |
| African | 122 | 280 | 0.72(0.50 to 1.04) | |
| European | 3463 | 30277 | 0.82(0.77 to 0.87) | |
| Middle East | 145 | 479 | 0.90(0.65 to 1.24) | |
| South Asian | 232 | 651 | 0.77(0.56 to 1.04) | |
| **Drinking Status** | | | | |
| Ever | 4040 | 30490 | 0.88(0.75 to 1.05) | 0.65 |
| Never | 1216 | 3106 | 0.84(0.71 to 0.99) | |
| **Sex** | | | | |
| Female | 1805 | 16753 | 0.89(0.71 to 1.12) | 0.78 |
| Male | 3731 | 19454 | 0.86(0.74 to 1.00) | |
| **Smoking Status** | | | | |
| Former | 1556 | 11589 | 0.85(0.70 to 1.03) | 0.86 |
| Never | 1483 | 17626 | 0.88(0.77 to 1.01) | |
| **Subsite** | | | | |
| Hypopharynx | 872 | 35556 | 0.84(0.76 to 0.94) | 0.51 |
| Larynx | 4374 | 36207 | 0.89(0.76 to 1.04) | |
| Oral cavity | 5536 | 36207 | 0.86(0.75 to 1.00) | |
| Oropharynx (HPV-) | 1452 | 35077 | 0.95(0.81 to 1.10) | |
| Oropharynx (HPV+) | 2182 | 34797 | 0.82(0.76 to 0.88) | |
| **GeographicRegion** | | | | |
| Europe | 2269 | 28929 | 0.91(0.69 to 1.20) | 0.79 |
| NorthAmerica | 1719 | 3817 | 0.82(0.66 to 1.02) | |
| SouthAmerica | 1169 | 2294 | 0.93(0.64 to 1.35) | |

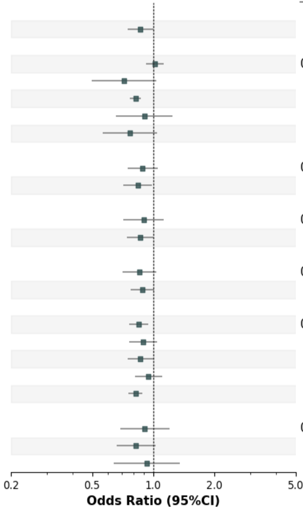


## rs9267280

| Variable | NO.Case | NO.Control | OR(95% Conf. Int.) | P-Heterogeneity |
|---|---|---|---|---|
| **Cross-ancestry meta-analysis** | | | | |
| OralCavity | 5536 | 36207 | 1.17(0.94 to 1.45) | |
| **Genetic ancestry** | | | | |
| Admixed | 1574 | 4520 | 0.95(0.82 to 1.11) | 0.01 |
| African | 122 | 280 | 1.44(0.84 to 2.46) | |
| European | 3463 | 30277 | 1.32(1.19 to 1.47) | |
| Middle East | 145 | 479 | 0.98(0.60 to 1.62) | |
| South Asian | 232 | 651 | 1.45(0.82 to 2.57) | |
| **Drinking Status** | | | | |
| Ever | 4040 | 30490 | 1.14(0.87 to 1.50) | 0.78 |
| Never | 1216 | 3106 | 1.20(0.98 to 1.47) | |
| **Sex** | | | | |
| Female | 1805 | 16753 | 1.19(0.89 to 1.59) | 0.90 |
| Male | 3731 | 19454 | 1.16(0.95 to 1.42) | |
| **Smoking Status** | | | | |
| Former | 1556 | 11589 | 1.13(0.78 to 1.64) | 0.83 |
| Never | 1483 | 17626 | 1.23(1.05 to 1.43) | |
| **Subsite** | | | | |
| Hypopharynx | 872 | 35556 | 1.06(0.87 to 1.30) | 0.66 |
| Larynx | 4374 | 36207 | 0.98(0.84 to 1.15) | |
| Oral cavity | 5536 | 36207 | 1.17(0.94 to 1.45) | |
| Oropharynx (HPV-) | 1452 | 35077 | 1.12(0.93 to 1.35) | |
| Oropharynx (HPV+) | 2182 | 34797 | 1.01(0.89 to 1.16) | |
| **GeographicRegion** | | | | |
| Europe | 2269 | 28929 | 1.10(0.74 to 1.63) | 0.72 |
| NorthAmerica | 1719 | 3817 | 1.23(0.68 to 2.21) | |
| SouthAmerica | 1169 | 2294 | 0.99(0.83 to 1.18) | |

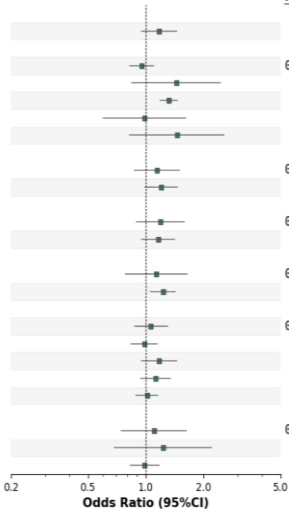

**f** **European (HPV+ oropharynx)**

HLA-B*51:01

| Variable | NO.Case | NO.Control | OR(95% Conf. Int.) | P-Heterogeneity |
|---|---|---|---|---|
| **Cross-ancestry meta-analysis** | | | | |
| Oropharynx (HPV+) | 2182 | 34797 | 1.37(0.69 to 2.72) | |
| **Genetic ancestry** | | | | |
| Admixed | 222 | 4520 | 0.94(0.61 to 1.45) | 0.00 |
| European | 1960 | 30277 | 1.90(1.55 to 2.31) | |
| **Drinking Status** | | | | |
| Ever | 1517 | 29911 | 1.38(0.61 to 3.10) | 0.78 |
| Never | 437 | 2296 | 1.57(0.99 to 2.49) | |
| **Sex** | | | | |
| Female | 367 | 16273 | 1.54(0.40 to 5.86) | 0.87 |
| Male | 1815 | 18524 | 1.36(0.80 to 2.34) | |
| **Smoking Status** | | | | |
| Former | 987 | 11432 | 1.30(0.58 to 2.95) | 0.68 |
| Never | 869 | 16855 | 1.87(1.39 to 2.51) | |
| **Subsite** | | | | |
| Hypopharynx | 872 | 35556 | 1.08(0.85 to 1.37) | 0.77 |
| Larynx | 4374 | 36207 | 1.10(0.97 to 1.24) | |
| Oral cavity | 5536 | 36207 | 1.20(1.08 to 1.34) | |
| Oropharynx (HPV-) | 1452 | 35077 | 1.15(0.88 to 1.49) | |
| Oropharynx (HPV+) | 2182 | 34797 | 1.37(0.69 to 2.72) | |
| **GeographicRegion** | | | | |
| Europe | 1086 | 28929 | 1.88(1.23 to 2.89) | 0.67 |
| NorthAmerica | 989 | 3633 | 1.69(1.27 to 2.25) | |

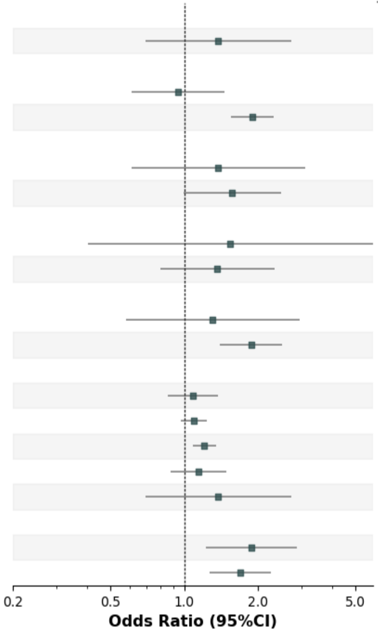

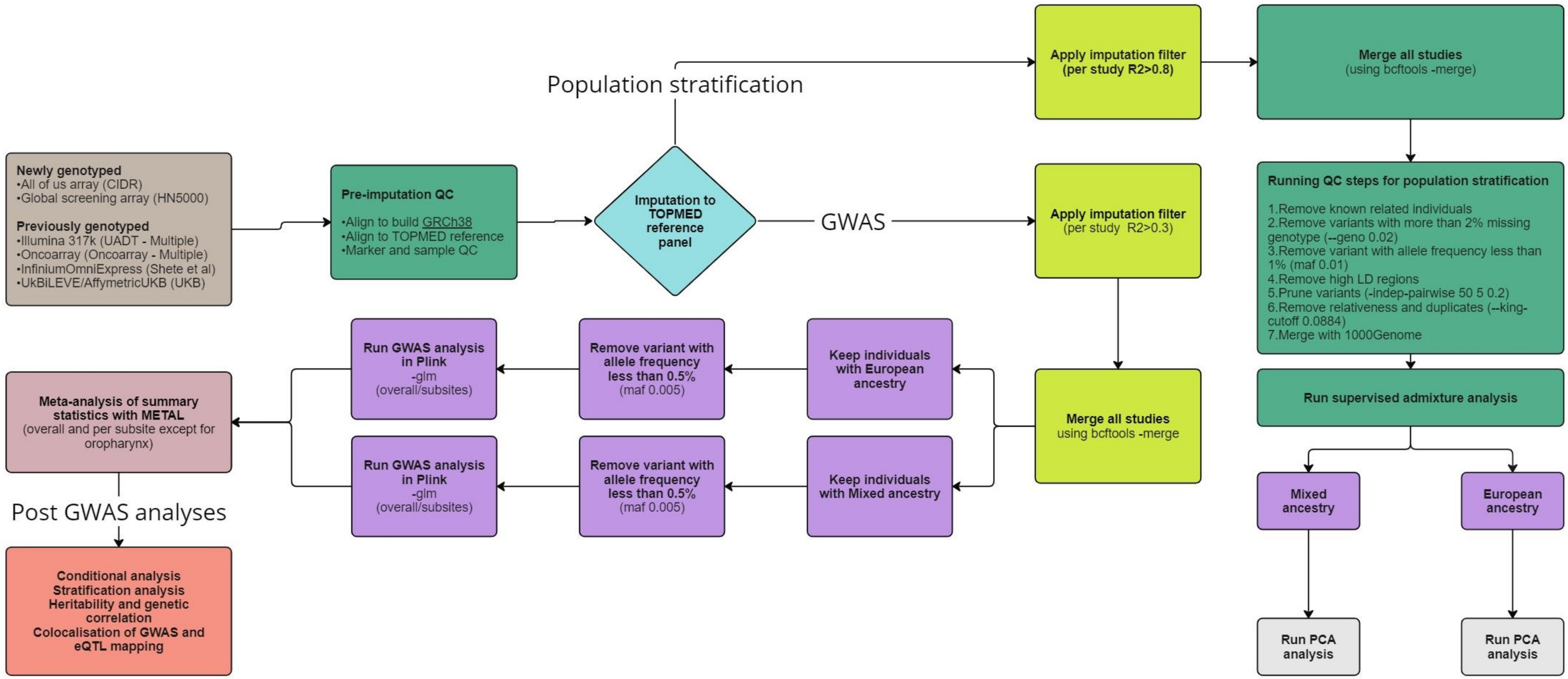


**Supplementary Figure 10. Flow diagram showing steps of genetic data curation and quality control (QC), imputation and analysis for GWAS.**

CIDR = Centre for Inherited Disease Research, UADT = Upper aerodigestive tract GWAS paper (McKay et al. 2011), UKB = UK Biobank, MAF = Minor allele frequency, LD = Linkage disequilibrium, PCA = Principal component analysis, eQTL= Expression quantitative trait loci (Created by Miro).

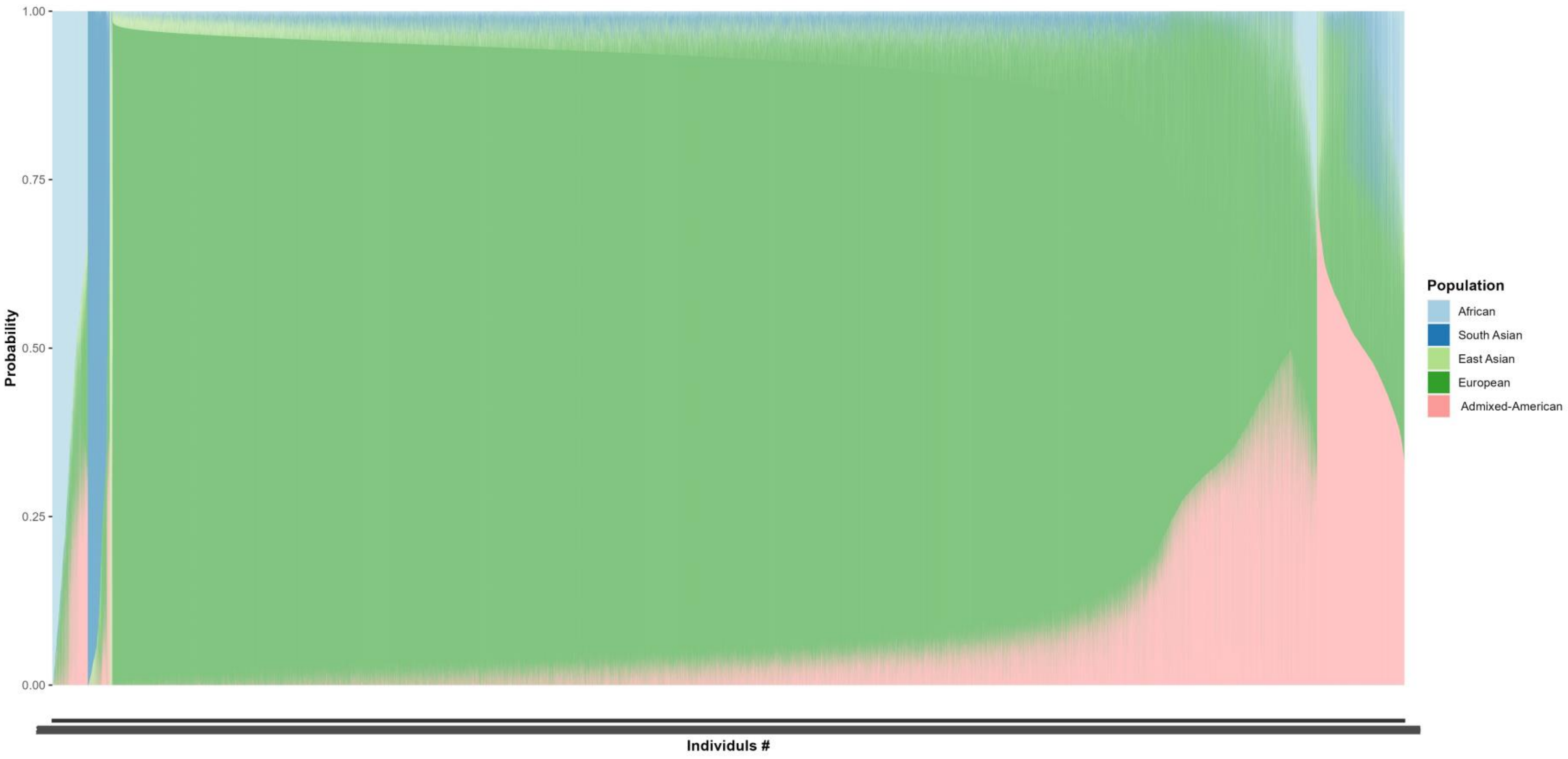


**Supplementary Figure 11. Population stratification using supervised Admixture analysis.**
Supervised ADMIXTURE analysis was conducted on a dataset comprising 61,129 individuals, with the 1000 Genome super populations serving as the reference which includes 2504 individuals. Each point on the x-axis corresponds to an individual, while the y-axis indicates their ancestry proportion.

**a**

| GWAS Group | Ancestry group | Case | | | | | Control |
|---|---|---|---|---|---|---|---|
| | | Oral cavity | Larynx | Hypopharynx | Oropharynx | Other | |
| **European** | European | 3511 | 2787 | 566 | 4134 | 2119 | 32914 |
| **Mixed** | African | 125 | 145 | 30 | 104 | 2 | 273 |
| | South Asian | 184 | 21 | 3 | 35 | 0 | 488 |
| | East Asian | 28 | 21 | 2 | 14 | 0 | 24 |
| | Admixed American | 6 | 8 | 0 | 7 | 0 | 30 |
| | Admixed | 1742 | 1427 | 297 | 1117 | 111 | 5128 |

**b**

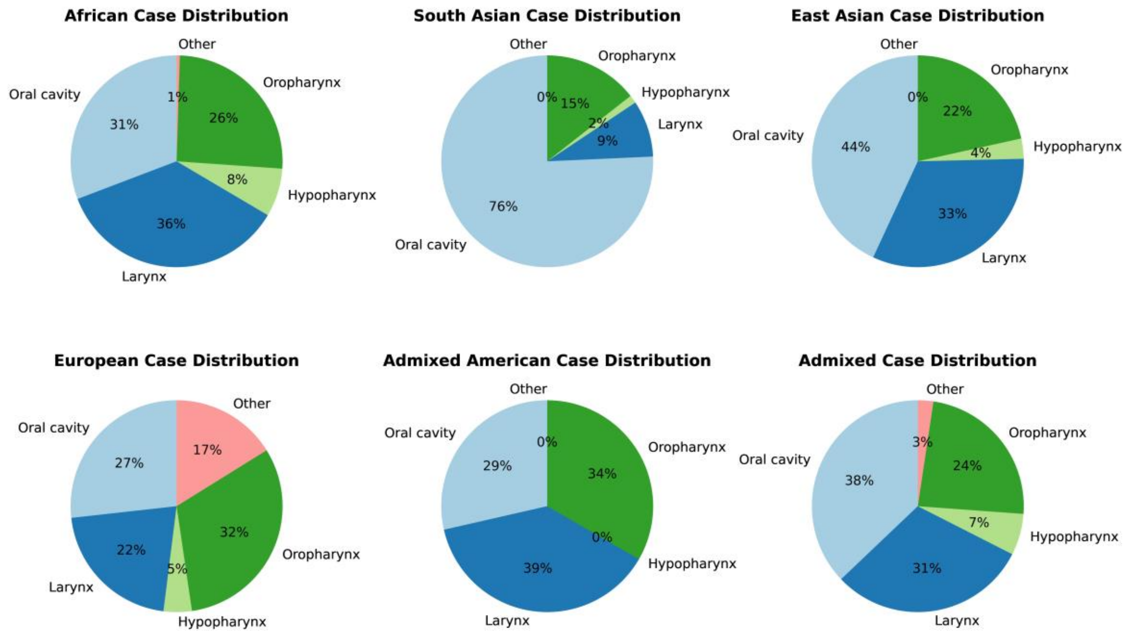


**Supplementary Figure 12. Ancestral composition and case distribution across each ancestral group.**

a) The classification of individuals into various ancestral groups was based on a 70% threshold, distinguishing those with dominant ancestry (≥70%) from those with admixed ancestry (<70%). Due to the small sample size of non-European dominant ancestries (N = 1,561), these individuals were combined with the admixed cases and collectively referred to as the “Mixed” ancestry group. b) Distribution of cancer cases among different ancestral groups. The “Other” case group includes cases with unknown primary site, overlapping sites, Not Otherwise Specified (NOS), or unavailable data.

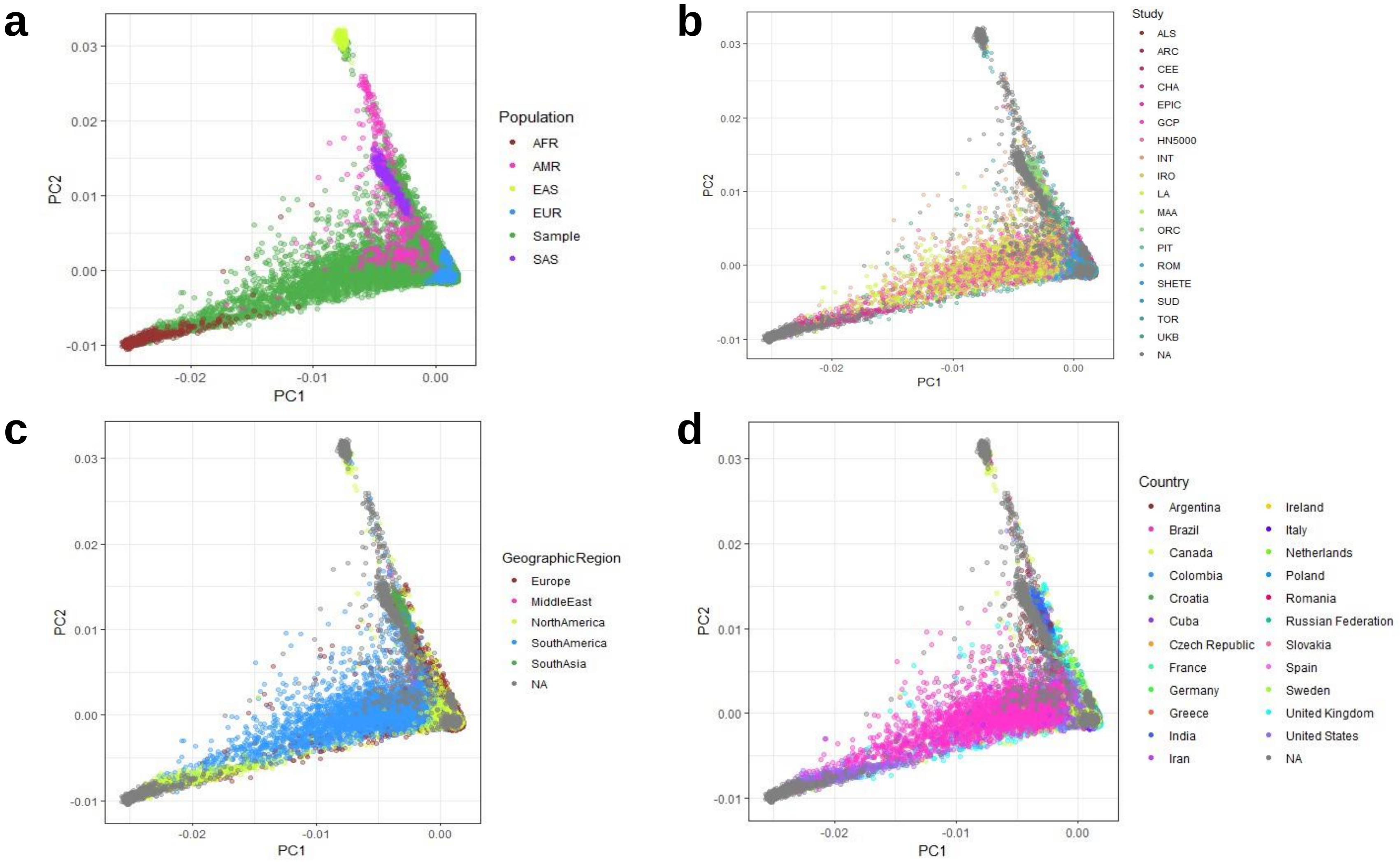


**Supplementary Figure 13. Principle component analysis.**

Principal Component Analysis (PCA) was conducted on a dataset of 61,129 individuals, including 2,504 individuals from the 1000 Genomes Project, representing five super populations as a reference. The analysis is visualized using Principal Components 1 (PC1) and 2 (PC2). a) The PCA plot shows the study population (green) compared to the five super populations from the 1000 Genomes Project (AFR: African; AMR: Admixed American; EAS: East Asian; EUR: European; SAS: South Asian). b) The study population is grouped by the geographic region of recruitment. c) The population is categorized by the study they were involved in. d) The population is displayed based on country. The 1000 Genomes super populations are shown in grey in panels b, c, and d.

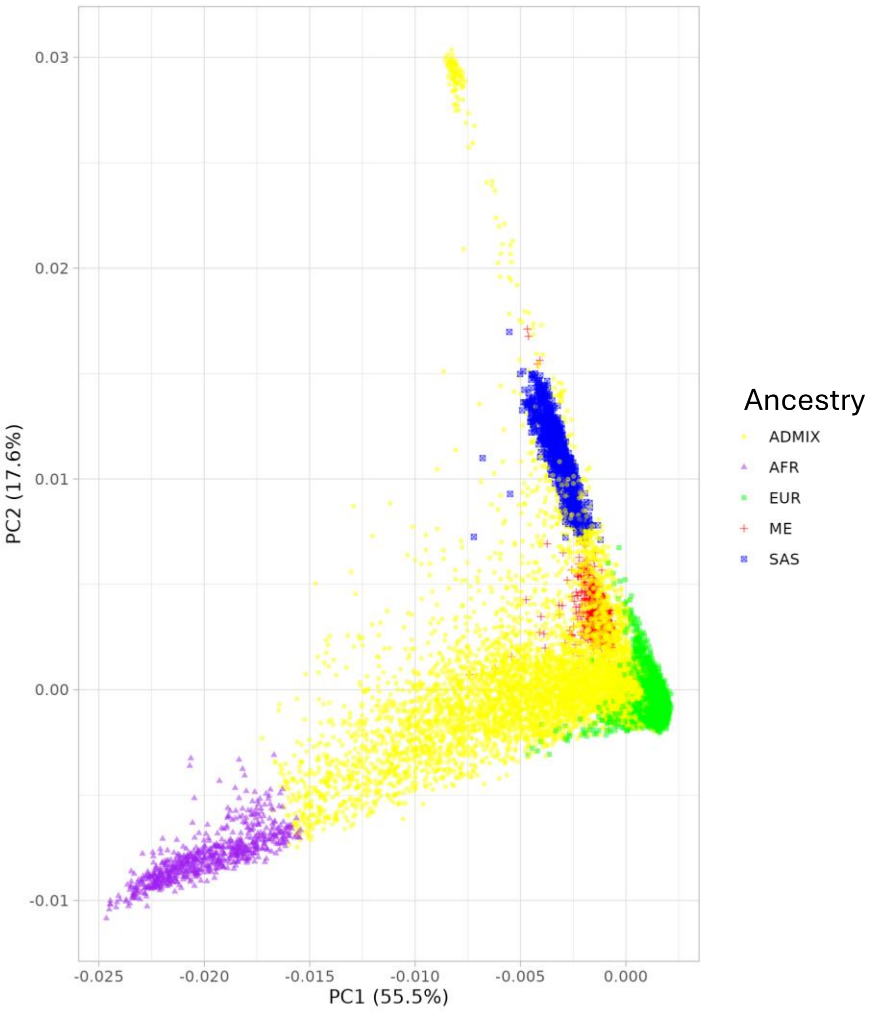


**Supplementary Figure 14. PCA plot of population stratification using supervised Admixture analysis for HLA analysis.** Principal Component Analysis (PCA) was conducted to represent five population ancestries. The analysis is visualized using Principal Components 1 (PC1) and 2 (PC2). AFR: African is coloured in purple; ADMIX in yellow; EUR: European in green; SAS: South Asian in blue and ME: Middle East in red.

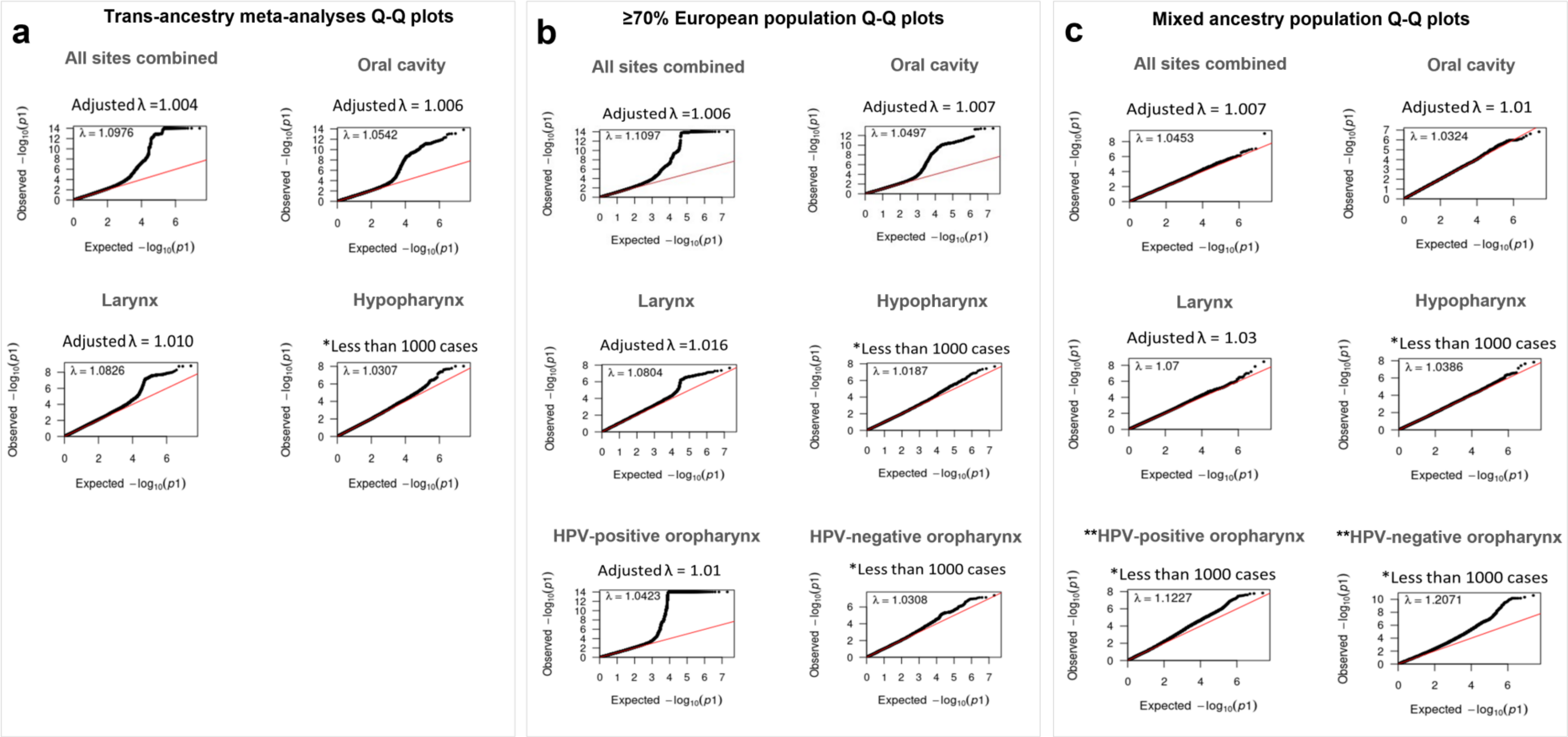


**Supplementary Figure 15. Cross ancestry and population-specific Q-Q (quantile-quantile) plots.** a) Q-Q plots from subsite analyses in cross-ancestry meta-analysis GWAS. b) Q-Q plots from GWAS with a predominant European sample representation (≥70%). c) Q-Q plots from analyses conducted on mixed ancestry populations. The red line represents the expected null distribution. "λ" = genomic inflation factor; "Adjusted λ" = genomic inflation factor for an equivalent study of 1,000 cases and 1,000 controls;

*For instances with fewer than 1,000 cases, only λ was reported.

**Since the Q-Q plot for HPV-positive and HPV-negative oropharyngeal cases in the mixed ancestry population showed inflation, the results were excluded from further analyses in this study.

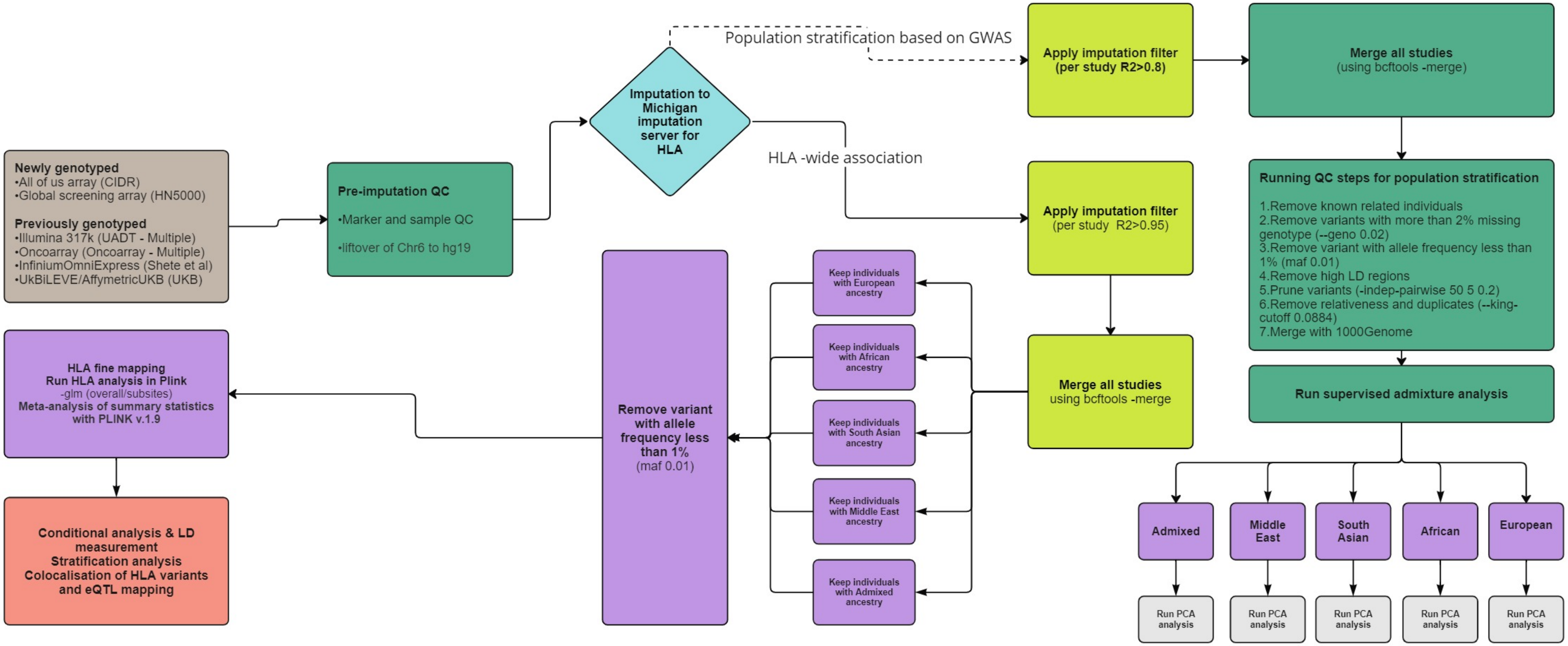


**Supplementary Figure 16.** Flow diagram showing steps of genetic data curation and quality control (QC), imputation and analysis for HLA fine mapping.
CIDR = Centre for Inherited Disease Research, UADT = Upper airway digestive tract GWAS paper (McKay et al. 2011), UKB = UK Biobank, MAF = Minor allele frequency, LD = Linkage disequilibrium, PCA = Principal component
