## Supplementary Notes for "Cross-ancestral GWAS identifies 29 novel variants across Head and Neck Cancer subsites"

### Supplementary Note 1: HN5000 ALSPAC combined imputation notes

This note describes the 'double imputation' method used to imputed the HN5000 cases with previously genotyped ALSPAC controls. Given the two studies have been genotyped on different arrays, simply combining the genotyped variants and imputing would mean the majority of variants from both studies are dropped as there is limited overlap between arrays.

The strategy here uses 2 rounds of imputation, the first allows high quality imputed variants to be used alongside variants genotyped on only one of the arrays for the second round of imputation.

ALSPAC re-genotyped – 411 ALSPAC participants who had previously been genotyped were re-genotyped alongside the HN5000 cases (DNA from immortalised cell lines). This allows some sensitivity analyses to be conducted with cases and controls genotyped together on the same array.

Schematic to show double imputation method:

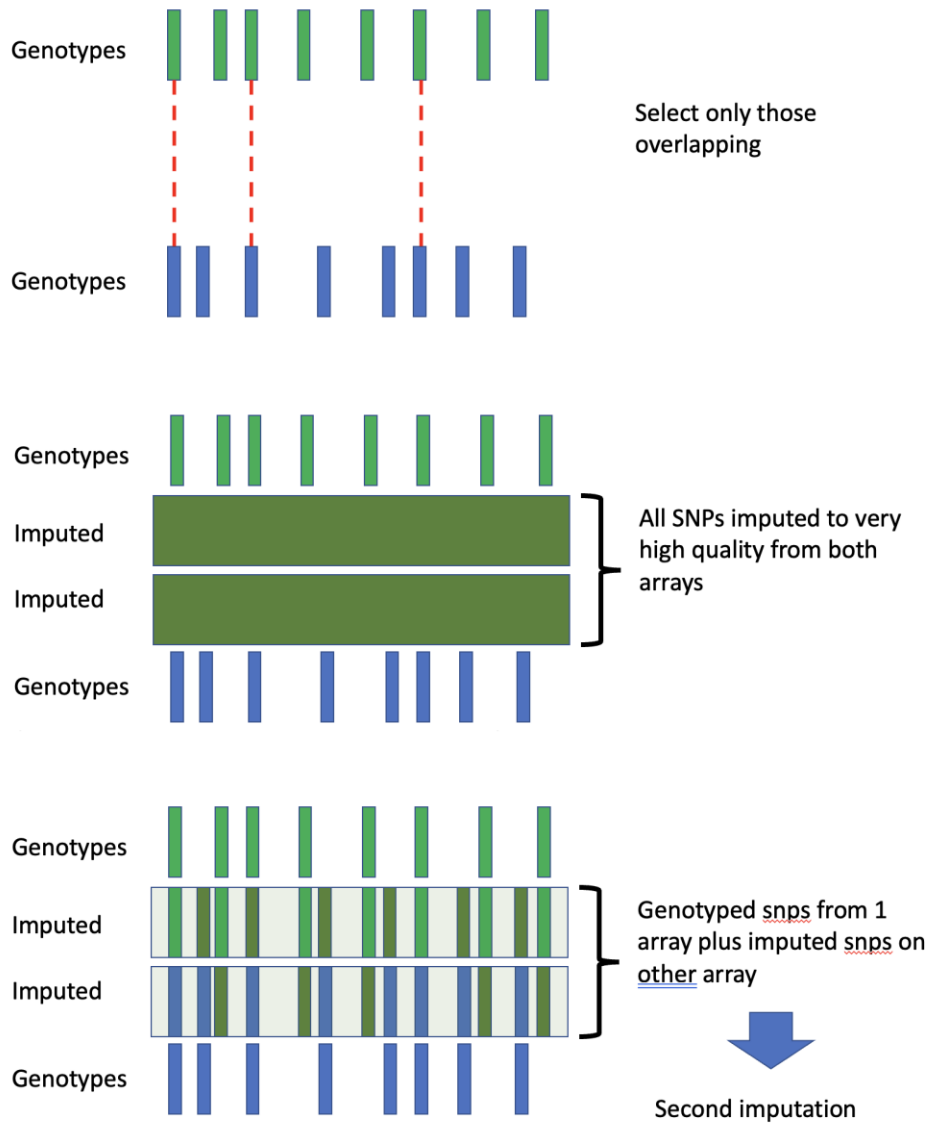

#### 1) Genotype calling from Intensity (IDat) files

- QC steps in GenomeStudio
- The GenomeStudio QC was run with HN5000 and ALSPAC (re-genotyped) samples combined.
- Sample QC
  - Import intensity files into GS
  - Cluster file from Ilumina: https://webdata.illumina.com/downloads/productfiles/global-screening-array/v2-0/gsa-24-v2-0-A1-cluster-file.zip
  - GSA manifest file from Illumina (build GRCh38): https://webdata.illumina.com/downloads/productfiles/global-screening-array/v2-0/infinium-global-screening-array-24-v2-0-a2-manifest-file-bpm.zip
  - Run sample calculations (to generate sample call rate)
  - Remove samples with <97% call rate (UK Biobank recommendation)
    - HN5000/ALSPAC – 71 samples removed
  - Recalculate SNP statistics
- SNP QC
  - Run SNP QC and zero SNPs not meeting thresholds (from Illumina technical note)
  - Sort SNP Table by Cluster Sep. zero SNPs with ≤0.3 (illumina recommendation)
    - HN5000/ALSPAC – 3865 removed
  - Sort SNP Table by call frequency (Call_Freq). Zero SNPs <0.97 (illumina rec)
    - HN5000/ALSPAC – 14504 removed
  - Sort SNP Table by AB R Mean, the mean normalized intensity (R) of the heterozygote cluster. This metric helps identify SNPs with low intensity data and has values increasing from 0. Zero SNPs ≤0.2
    - HN5000/ALSPAC – 38 removed
  - Sort SNP Table by AB T Mean, the mean of the normalized theta values of the heterozygote cluster. Zero SNPs ≤0.2
    - HN5000/ALSPAC –540 removed
- Update the sample statistics
- Sex mismatch
  - Add reported sex to Gender column
  - Estimate Gender – select all samples, right click select Estimate gender
  - Assess for mismatch and investigate possibility of mislabelled samples
    - HN5000/ALSPAC – 21 reported Female but GS reports Male
    - HN5000/ALSPAC – 18 reported Male but GS reports Female

#### 2) Round 1 Imputation

Each study was imputed to TOPMED imputation panel following completing QC steps

#### 3) Combine HN5000 and ALSPAC ready for round 2 imputation.

Aim: To create a set of variants from both studies which are either genotyped or imputed to a very high quality but also are informative for imputation. To ensure the variants are informative, the variants selected must be included on at least one of the arrays.

NB. In previous attempts, only selecting variants which are of high quality imputation does not ensure imputation informative variants are included in the combination - this resulted in poor quality round 2 imputation.

Following round 1 imputation:

- HN5000
  - 480,496 genotyped variants (a)
  - 307,174,027 imputed variants
- ALSPAC
  - 451,189 genotyped variants (a)
  - 306,899,501 imputed variants

Steps to select variants:

- Identify variants genotyped on both arrays
  - 94,289
- Identify variants genotyped in one study and imputed in the other
  - Imputed in HN5000 and genotyped in ALSPAC = 356,900
  - Imputed in ALSPAC and genotyped in HN5000 = 386,175
- Of these, identify those with high quality imputation (Rsq>0.90)
  - Imputed in HN5000 and genotyped in ALSPAC = 330,272 (b)
  - Imputed in ALSPAC and genotyped in HN5000 = 300,315 (b)
- Genotyped or high-quality imputation and on the other array (a + b)
  - HN5000 = 810,768
  - ALSPAC = 751,504
- Overlapping in both studies
  - 724,876
- Select these variants from both studies
- Combine the two studies
  - Bcftools merge –force-samples
- Convert imputed variants to 'Best Guess' genotypes ready for imputation
  - bcftools annotate -x FOMAT

Check imputation quality in the 2 studies of the overlapping variants

- High quality and similar

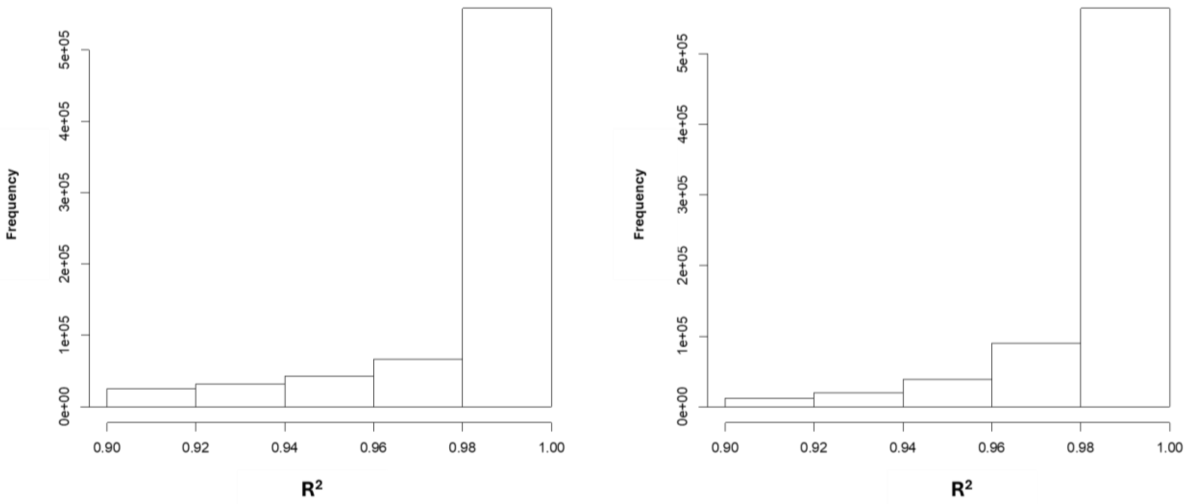


Supplementary Note Figure 1: Distribution of variants in imputation quality ($R^2$) bins in ALSPAC and HN5000.

**4) Round 2 imputation**

The combined studied are then imputed together to TOPMED

- Allele frequency r2 between uploaded samples and reference panel = 0.942
